# Multi-population Genome-Wide Association Study Identifies Multiple Novel Loci associated with Asymptomatic Intracranial Large Artery Stenosis

**DOI:** 10.1101/2025.05.06.25327093

**Authors:** Minghua Liu, Farid Khasiyev, Antonio Spagnolo-Allende, Danurys L Sanchez, Howard Andrews, Qiong Yang, Alexa Beiser, Ye Qiao, Jose Rafael Romero, Tatjana Rundek, Adam M Brickman, Jennifer J Manly, Mitchell SV Elkind, Sudha Seshadri, Christopher Chen, Oscar H Del Brutto, Saima Hilal, Bruce A Wasserman, Giuseppe Tosto, Myriam Fornage, Jose Gutierrez

## Abstract

**Background:** Intracranial large artery stenosis (ILAS) is one of the most common causes of stroke worldwide and is associated with the risk for future vascular events. Asymptomatic ILAS is a frequent finding on neuroimaging and shares many risk factors with atherosclerotic vascular disease. Whether asymptomatic ILAS is driven by genetic variants is not well-understood.

**Methods and Results:** This study included 4960 participants from seven geographically diverse population-based cohorts (34% Whites, 16% African Americans, 22% Hispanics, 24% Asians, 5% native Ecuadorians). We defined asymptomatic ILAS as luminal stenosis > 50% in any large brain artery using time-of-flight magnetic resonance angiography (MRA). A genome-wide association study revealed one variant in *RP11-552D8.1* (rs75615271; OR, 1.22 [1.11-1.33]; *P*=4.85×10^−8^) associated with global ILAS at genome-wide significance (*P*<5×10^−8^). Gene-based association analysis identified a gene-set enriched in chr1q32 region, including *NEK2*, *LPGAT1*, *INTS7*, *DTL*, and *TMEM206*, in global ILAS (*P*=1.34 ×10^−7^) and anterior ILAS (*P*=1.77 ×10^−8^).

**Conclusion:** This study reveals one variant rs75615271 associated with asymptomatic ILAS in a multi-population. Further functional studies may help elucidate the role that this variant plays in the pathophysiology of asymptomatic ILAS.

## Background

Intracranial large artery stenosis (ILAS), most often caused by intracranial atherosclerotic disease, is one of the common causes of stroke and is associated with the risk for future vascular events ^1–3^. ILAS is considered to be a major cause of stroke, accounting for 30-50% of cases of ischemic stroke in the Asian, non-Hispanic Black, and Hispanic populations, but only about 10% in non-Hispanic whites of European descent^4^. The prevalence of ILAS varies greatly according to racial/ethnic origin, e.g., ILAS is much less frequent in Western countries than in Asia^5^. Various explanations for racial/ethnic differences in the prevalence of ILAS have been advanced, including genetic susceptibility of intracranial vessels to atherosclerosis, as well as differences in lifestyle and risk factors, such as hypertension, diabetes, dyslipidemia, and smoking^6^. Previously, a case-control study revealed that variant *RNF213* c.14429G > A (p.Arg4810Lys, rs112735431) has a strong association with ILAS^7^. A recent study revealed that *RNF213* p.Arg4810Lys also increases the risk of ischemic stroke due to intracranial artery atherosclerosis^8^. Moreover, *RNF213* p.Arg4810Lys was associated with coronary artery disease and pulmonary hypertension^9–11^. The association between *RNF213* and stenosis of the intracranial, coronary, and other systemic arteries indicates that genetic traits may partially account for intracranial stenosis, either by predisposing vascular risk factors or by a direct contribution to an established atherosclerotic mechanism. Other candidate genes implicated in previous genetics studies with ILAS include *ADIPOQ*, *PDE4D*, *LPL*, and *CYP11B2*^12–15^. Asymptomatic ILAS is a frequent finding on neuroimaging. The prevalence of asymptomatic ILAS increases with age in patients with transient ischemic attack and minor stroke^16^. A study in a stroke-free population indicated that asymptomatic ILAS is a risk factor for cerebral and systemic vascular events with risk increasing as stenosis severity worsens ^17^. Asymptomatic ILAS shares many risk factors with atherosclerotic vascular disease. However, whether asymptomatic ILAS is driven by common and rare genetics is not well-understood. In this study, we performed a multi-population discovery genome-wide association study (GWAS) analysis in diverse population cohorts within and outside the United States. We detailed the variants, genes, and biologic pathways relevant to the genetic architecture of asymptomatic ILAS and described the similarities and differences between asymptomatic ILAS and other clinical cardiovascular diseases (stroke, coronary artery disease, atrial fibrillation, etc.) using Mendelian randomization, gene association analysis, and expression quantitative trait loci colocalization, and by exploring shared overlap with other large population intracranial stenosis GWAS research.

## Method

### Sampled populations

#### Atherosclerosis Risk in Communities (ARIC) study

The ARIC study is a population-based prospective cohort investigating vascular risks. It includes 15,792 persons aged 45-64 years at baseline (1987-1989), randomly selected from four US communities^18^. Participants completed seven clinic examinations conducted from 1987 to 2019. The institutional review boards at the collaborating medical institutions (The Johns Hopkins University, University of North Carolina at Chapel Hill, Wake Forest University, University of Mississippi Medical Center, and University of Minnesota) approved the study. All participants provided written informed consent.

#### The Northern Manhattan Study (NOMAS)

NOMAS is a prospective cohort initially focused on determining the incidence of stroke and vascular events in a diverse urban population^19^. Participants were recruited through random digit dialing from 1993 to 2001. Written informed consent was provided by all study participants. The study was approved by institutional review boards at Columbia University Medical Center and the University of Miami.

#### Washington Heights–Inwood Columbia Aging Project (WHICAP study)

WHICAP is a prospective, population-based study focused on aging and dementia. Established through multiple recruitment waves, participants were first recruited in 1992 from a random sample of Medicare-eligible adults residing in the neighborhoods of Washington Heights and Inwood in northern Manhattan. Participants undergo evaluations every 18-24 months, which include a comprehensive neuropsychological battery, medical and neurologic examination, and health-related survey^20^. Dementia and its subtypes are determined in a consensus conference involving neurologists and neuropsychologists. The study was approved by the institutional review board at Columbia University Medical Center. All participants provided written informed consent.

#### Epidemiology of Dementia In Singapore (EDIS study)

The EDIS study is a cross-sectional design drawing participants who were long-term local citizens from the Singapore Epidemiology of Eye Disease (SEED) study, consisting of the Singapore Chinese Eye Study, Singapore Malay Eye Study-2, and the Singapore Indian Eye Study-2, aged 60 years and above ^21–23^. Specific enrolment targets were set for the Chinese, Malay, and Indian communities to ensure that the final cohort was proportionate to Singapore’s ethnic composition. The Singapore Eye Research Institute Review Board approved the study. Bilingual coordinators obtained written informed consent from participants in their preferred language prior to enrollment.

#### Memory Clinic in Singapore (MCS study)

The MCS study involved patients attending the memory clinics at National University Hospital and St. Luke’s Hospital from 2009 to 2015. Patients were referred by primary, secondary and tertiary care facilities due to consistent memory complaints and were assessed by a team of clinicians, psychologists, and nurses in the Memory Aging and Cognition Center, National University of Singapore. The institutional review board at National University Hospital approved the study. All participants provided written informed consent.

#### Framingham Heart Study (FHS)

FHS is a prospective, community-based cohort. Participants who survived to the 7th examination were invited to undergo a brain MRI between 1999 to 2005, with a final sample of 2144 stroke-free individuals. For these analyses, we used an FHS subsample with available MRA as part of the stroke case study. The study was approved by the institutional review board at Boston Medical Center. All participants provided written informed consent.

#### The Atahualpa Project (TAP)

TAP is a population-based study designed to evaluate prevalence, incidence, and correlates of major neurological and cardiovascular disorders in community residents aged ≥ 40 years in rural Ecuador. The neuroimaging sub-study enrolled all Atahualpa residents aged ≥ 60 years who had no contraindications for magnetic resonance imaging and provided the informed consent ^24^. The study was approved by the Institutional Review Board of Hospital-Clínica Kennedy, Guayaquil.

We followed the STrengthening the Reporting of OBservational studies in Epidemiology (STROBE) reporting guidelines for cohort studies ^25^.

### Measurement of Asymptomatic ILAS

The software LKEB Automated Vessel Analysis (LAVA) (Leiden University Medical Center, The Netherlands, build date October 19th, 2018) was utilized to view all MRA images that collected from each site; then analyze the images centrally and determine the presence and severity of intracranial arterial stenosis. Briefly, this software uses a flexible 3D tubular Non-Uniform Rational B-Splines model to automatically identify the margins of the arterial lumen based on voxel intensity with excellent reliability ^12^. A prespecified and harmonized imaging analysis protocol ^26^ was employed to measure cross-sectional arterial diameters for up to 13 intracranial arteries per participant, including left and right anterior cerebral arteries (ACAs), middle cerebral arteries (MCAs), internal carotid arteries (ICAs), posterior cerebral arteries (PCAs), posterior communicating arteries (PCOMs) and intracranial portion of vertebral arteries (VAs); and the basilar artery (BA). The severity of intracranial stenosis was evaluated and quantified by two trained neurologists independently according to the narrowest lumen area compared with the immediately preceding normal segment, or the next normal appearing lumen if the stenosis was at the arterial origin. Stenosis was classified as clinically relevant when it was estimated to be equal to or larger than 50% of the normal lumen ^27^. Asymptomatic ILAS was defined as presence of one or more stenotic vessels on MRA, where the stenosis is equal to or larger than 50% of the normal lumen. Anterior ILAS referred to asymptomatic ILAS in ICAs, MCAs, ACAs, and PCOMs; posterior ILAS referred to asymptomatic ILAS in PCAs, VAs and BA; global ILAS referred to asymptomatic ILAS detected in any of the intracranial arteries.

### Genome-Wide Association Study

Description of genotyping, quality control and imputation in each study is provided in supplemental Table 1. In brief, sample and variant quality control criteria included the following: 1) <10% missingness of genotype calls, 2) Hardy-Weinberg Equilibrium p-value <1×10^-6^, 3) imputation quality < 0.3, 4) minor allele frequency (MAF) <0.0001, 5) call rate < 97.5% for MAF > 1% and call rate < 99% for MAF < 1%.

GWAS was performed separately for each race/ethnicity ^28^ group in each cohort. We used multiple logistic regression in PLINK with adjustment for age, sex, and three principal components, and filtered for variants with minor allele frequency >1%. EasyQC was employed to conduct quality control for GWAS results. Multi-population results were determined using a fixed-effect inverse-variance-based method implemented in METAL ^29^. Variants with minor allele frequencies (MAF) < 1% and those not present in at least two studies were excluded after the meta-analyses. Cross-study heterogeneity was assessed using Cochran’s Q-test, and variants with heterogeneity p-value <0.05 were excluded. Population-specific meta-analyses were performed to identify population-specific variants. An association with a *P* < 5×10^-8^ was considered genome-wide significant.

GWAS lead SNPs (*P* < 1×10^-5^) and genomic regions were annotated on the web-based platform FUMA (Functional Mapping and Annotation of Genome-Wide Association Studies)^30^. Briefly, pairwise linkage disequilibrium statistics were calculated from a 1000 Genomes Project Phase 3 reference panel (mixed for multi-population meta-analysis, White, African American, American admixed for Hispanic individual, Asian, and Amerindian). Independent GWAS loci were identified using R^2^ < 0.6 (the default parameter in FUMA) and independent lead single nucleotide polymorphisms (SNPs) were further selected from the set of independent GWAS variants using R^2^ < 0.1. Candidate SNPs are in LD with any of the independent significant SNPs at R^2^ > 0.6. A genomic region was then determined using the calculated linkage-disequilibrium structure (multiple independent SNPs were merged if < 250 kb from each linkage-disequilibrium block).

### Gene-based association and gene enrichment analysis

All GWAS variants were annotated by location (intergenic, intron, exon) and nearest gene using the single nucleotide polymorphism database (dbSNP) of nucleotide sequence information. Gene-set analysis was performed for multi-population results using Multi-Marker Analysis of GenoMic Annotation (MAGMA)^31^ implemented in the FUMA^30^. Gene sets were obtained from Msigdb v7.0^32^. Gene associations were considered significant if they met a p-value < 2.6×10^-6^ (0.05/19,021 protein coding genes). Candidate SNPs were mapped to the nearest gene within 50kb or an expression quantitative trait locus (eQTL) genes in Genotype-Tissue Expression (GTEx) project data version 8 (v8)^33^.

Mutation intolerance was calculated by probability of being loss-of-function intolerant (pLI) score from ExAC database^34^ and non-coding residual variation intolerance score (ncRVIS)^35^. The higher the pLI score is, the more intolerant to loss-of-function mutations the gene is. The higher the ncRVIS is, the more intolerant to noncoding variants the gene is. To explore the interaction between the target region and multiple genes, chromatin interaction mapping was performed. The Hi-C data was used to identify with significant chromatin interactions at FDR= 1×10^-6^ ^36^. To test the relationship between highly expressed genes in a specific tissue and genetic association, gene-property analysis is performed using average expression of genes per tissue type as a gene covariate. Gene expression values are log2 transformed average Reads Per Kilobase of transcript per Million mapped reads (RPKM) per tissue type based on GTEx RNA-seq data. Tissue expression analysis is performed for 30 general tissue types and 53 specific tissue types in GTEx v8 database, separately. MAGMA was performed using the result of gene analysis (gene-based P-value) and tested for one side (greater) with conditioning on average expression across all tissue types.

### Mendelian Randomization Analysis

Mendelian randomization was performed to select biomarkers previously identified as risk factors or relevant to pathobiology for asymptomatic ILAS with ischemic stroke^37^, small vessel stroke^37^, atrial fibrillation (AF)^38^, and coronary artery disease^39^. To minimize bias from correlated instruments, variants with asymptomatic ILAS association p-value <1.0x10^-5^ were LD-clumped at r^2^ < 0.01 ^40^ against the 1000 Genome LD reference calculated for White, African American, Asian and Hispanic, and Ecuadorian populations. Variants with MAFs < 0.01 in the reference population were excluded from MR analysis. Causal association was primarily evaluated using the inverse-variance weighted (IVW) method, additionally performed sensitivity analysis using simple median-based method, weighted median-based method, and MR-Egger method. All MR analyses were performed using the “TwoSampleMR” R package ^40^.

## Results

### Multi-population GWAS Identifies a Novel Locus Associated with Asymptomatic ILAS

We conducted a multi-population GWAS for asymptomatic ILAS levels in 4960 participants, including 1677 Whites, 799 African Americans, 1064 Hispanics, 1191 Asians, and 229 native Ecuadorian. Mean age of the participants across studies ranged from 67 to 76 years, with proportions of women ranging from 40% to 64%. Detailed demographic information is presented in Table 1.

**Table 1.** Demographic information of studies.

| **Cohort** | **ARIC** | **NOMAS** | **EDIS** | **MCS** | **FHS** | **Atahualpa** | **WHICAP** |
| --- | --- | --- | --- | --- | --- | --- | --- |
| N = 4960 | 1565 | 1109 | 841 | 350 | 206 | 229 | 660 |
| Age, mean (sd) | 75.8<br>(5.3) | 70.1<br>(8.4) | 70.2<br>(6.6) | 71.0<br>(8.2) | 72.7<br>(11.7) | 66.7 (6.4) | 76.4<br>(6.2) |
| Female (%) | 58.8 | 60.5 | 51.7 | 55.7 | 54.3 | 40.1 | 64 |
| Ancestry (%) |  |  |  |  |  |  |  |
| European | 74.3 | 13.8 | - | - | 100 | - | 23.4 |
| African | 25.7 | 16.6 | - | - | - | - | 32.2 |
| Hispanic | - | 69.6 | - | - | - | - | 44.2 |
| Asian | - | - | 100 | 100 | - | - |  |
| Native Ecuadorian | - | - | - | - | - | 100 |  |
| Hypertension (%) | 74.9 | 68.1 | 80.8 | 67.8 | 68.4 | 47.6 | 69 |
| Diabetes (%) | 33.9 | 19 | 37.3 | 31.8 | 23.9 | 32.3 | 30.1 |
| Dyslipidemia (%) | 56 | 45 | 76 | 71.4 | 51.6 | 17 | 51.5 |
| Current Smoking (%) | 6 | 52.7 | 27.9 | 7.1 | 9.3 | 1.7 | 5.3 |
| Asymptomatic ILAS (%) | 10.4 | 7.4 | 7.7 | 20.7 | 21 | 6.1 | 9.8 |
| Anterior asymptomatic ILAS (%) | 4.5 | 4.9 | 3.6 | 17.1 | 16.6 | 5.2 | 7.2 |
| Posterior asymptomatic ILAS (%) | 7.6 | 2.8 | 5.1 | 7.5 | 9 | 1.3 | 4 |

We identified one variant rs75615271 (*RP11-552D8.1*) associated with global ILAS at genome-wide significance (*P*<5×10^−8^; Figure 1A). One copy of the A allele (AF=0.11) for rs75615271 (OR, 1.22 [1.11-1.33]; *P*=4.85×10^−8^) was associated with increased risk of global ILAS (Table 2). We did not find the variants associated with anterior or posterior ILAS at genome-wide significance. However, we identified rs75615271 as the lead variant that was associated with anterior ILAS (OR, 1.16 [1.08-1.25]; *P*=2.19×10^−7^). The genomic control lambda value for was 1.03 for global ILAS GWAS, 1.02 for anterior ILAS GWAS and 1.08 for posterior ILAS GWAS (Figure 2). We did not observe population-specific variants related to asymptomatic ILAS based on the population-specific meta-analyses at genome-wide significance (Supplemental Figure 1-6, Supplemental Table 2).

**Figure 1.**
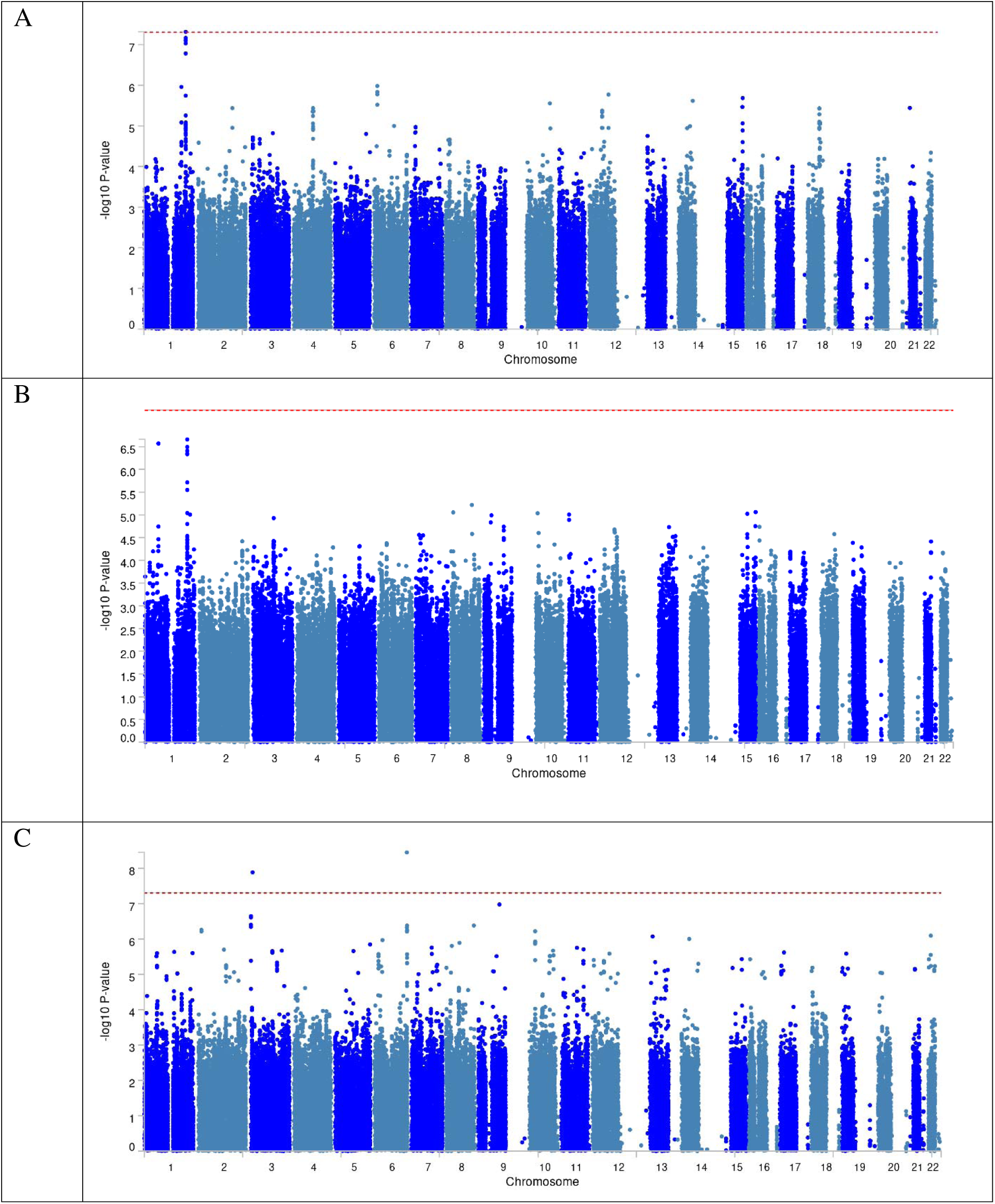
Manhattan Plot of GWAS summary statistics. (A) Global ILAS. (B) Anterior ILAS. (C) Posterior ILAS.

**Figure 2.**
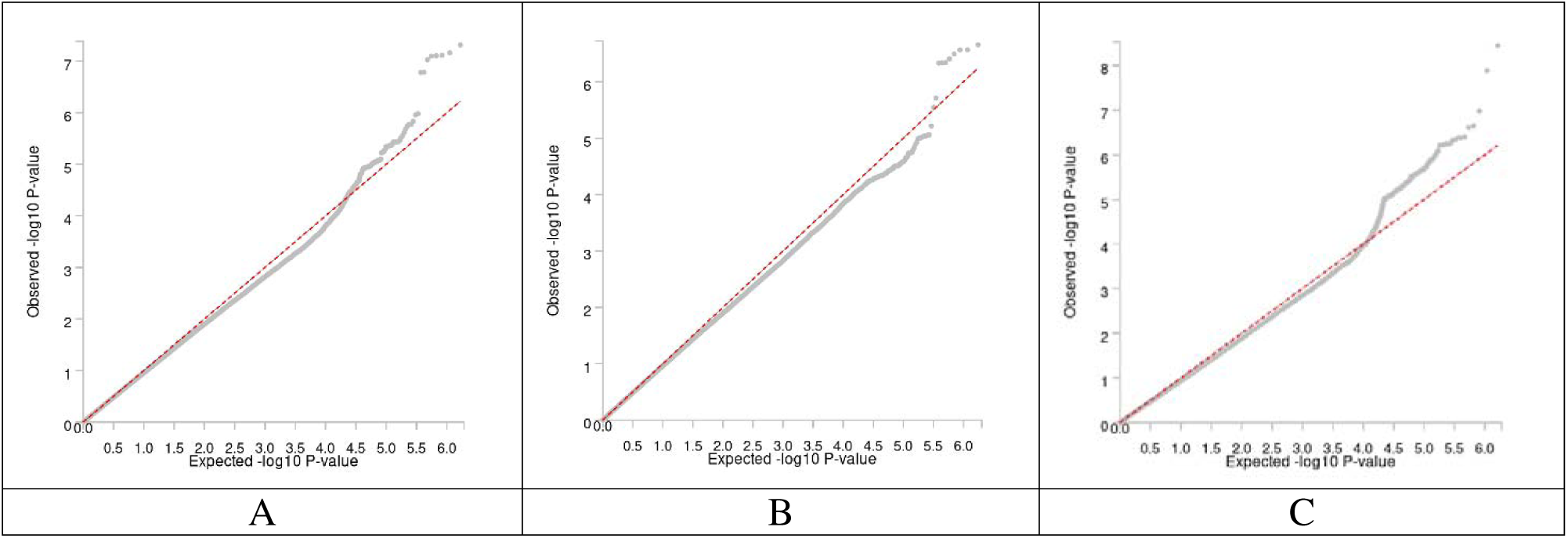
QQ plot. (A) Global ILAS. (B) Anterior ILAS. (C) Posterior ILAS.

**Figure 3.**
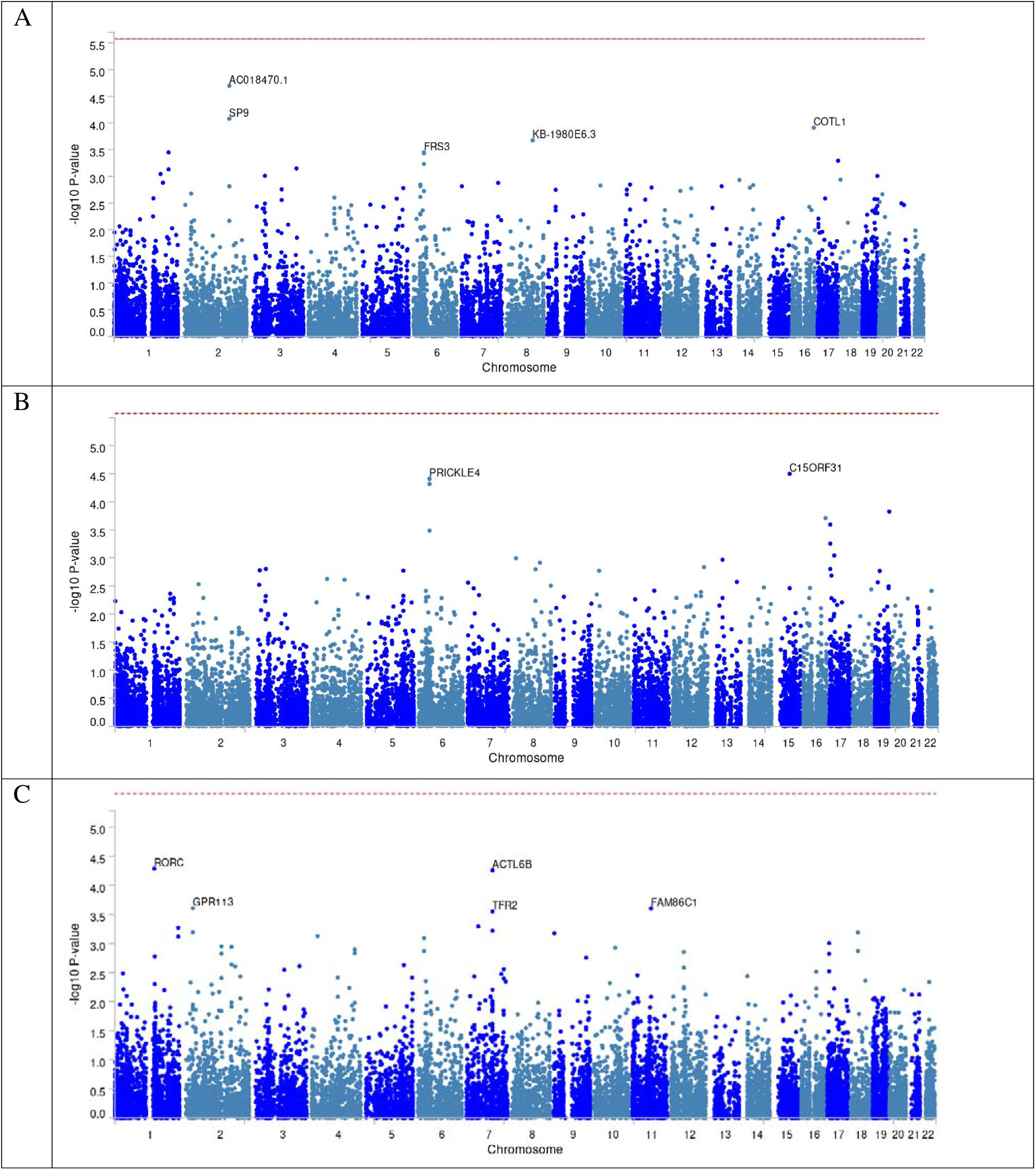
Mahattan Plot of gene-based test. (A) Global ILAS. (B) Anterior ILAS. (C) Posterior ILAS.

**Figure 4.**
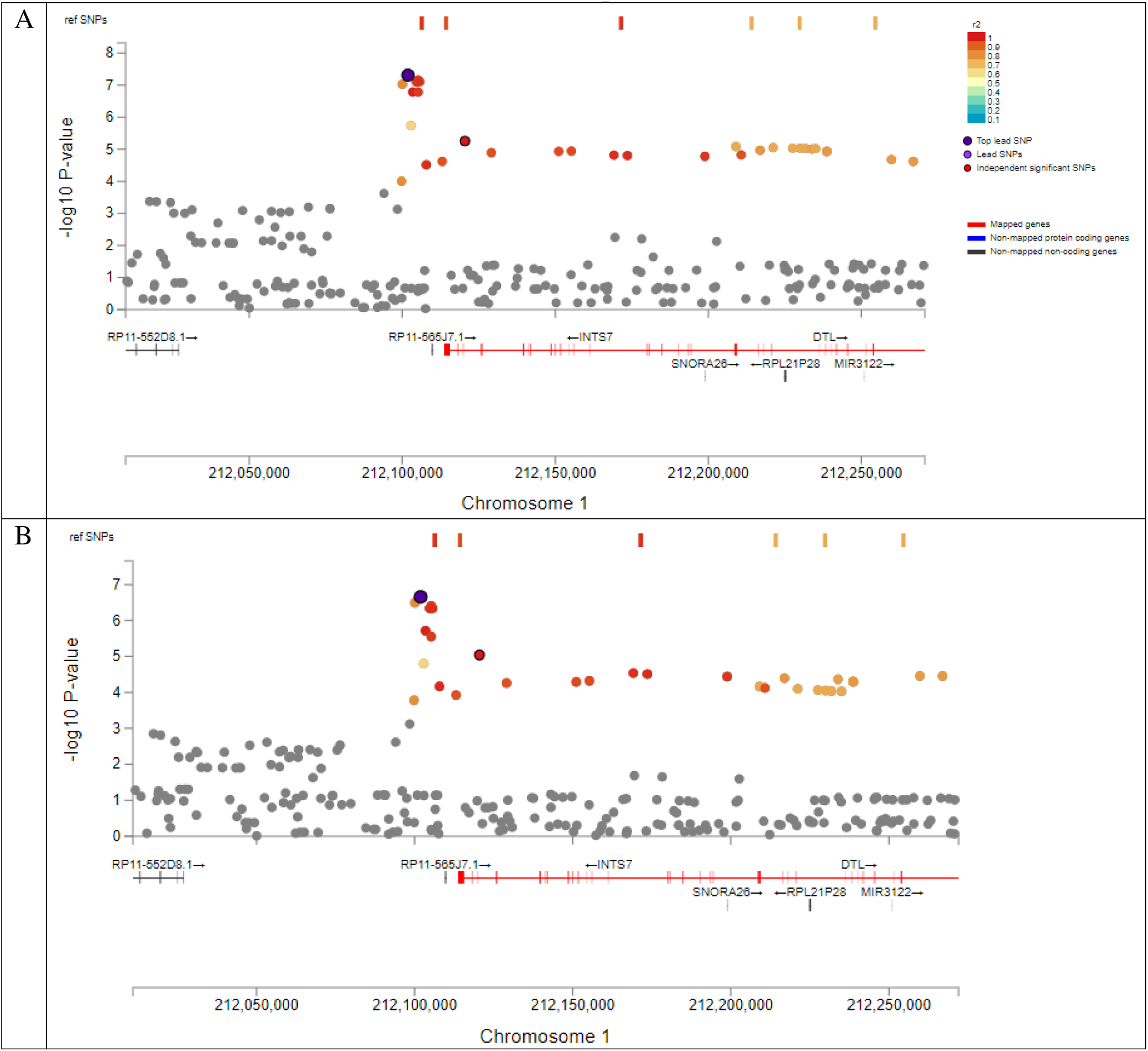
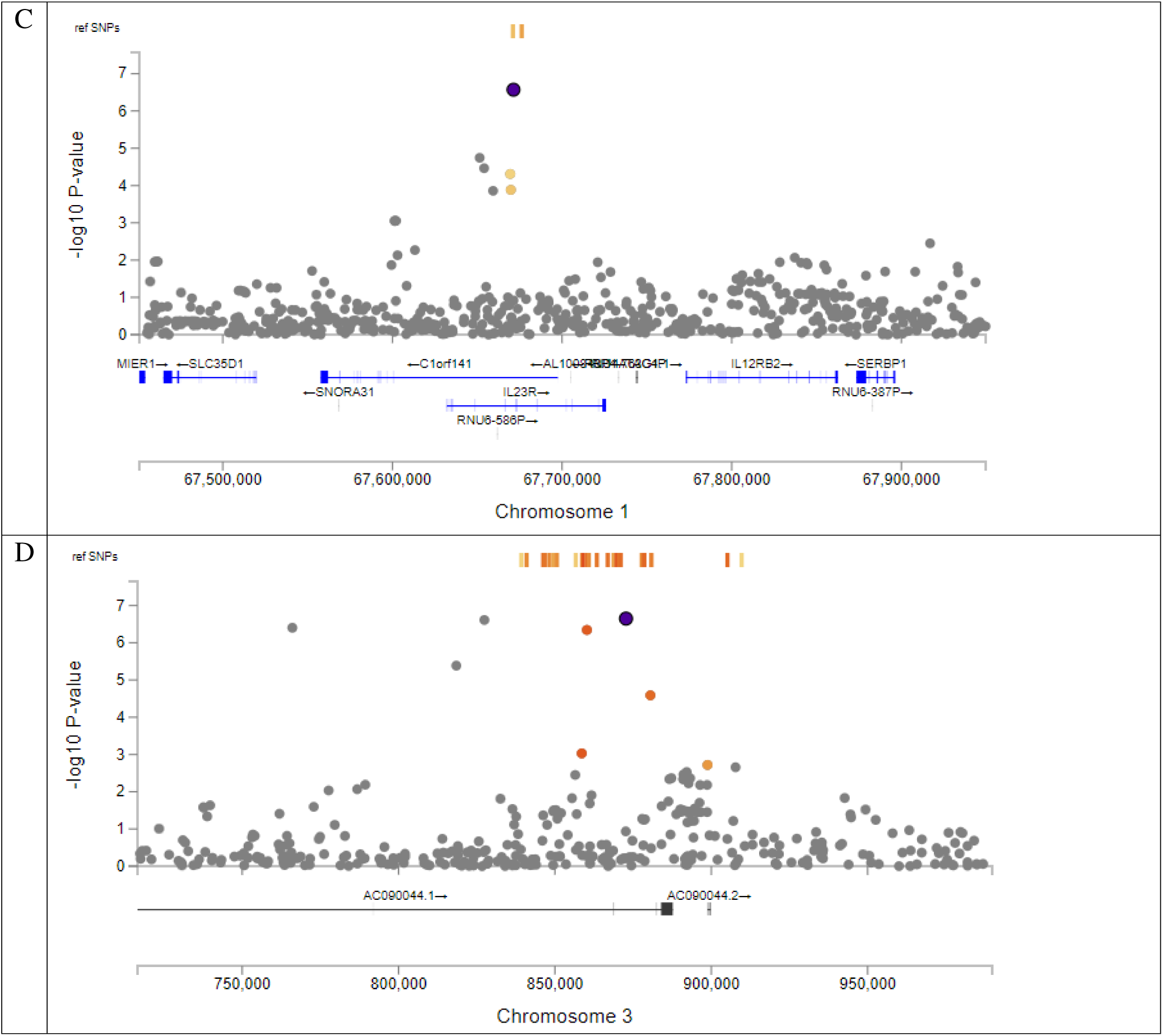
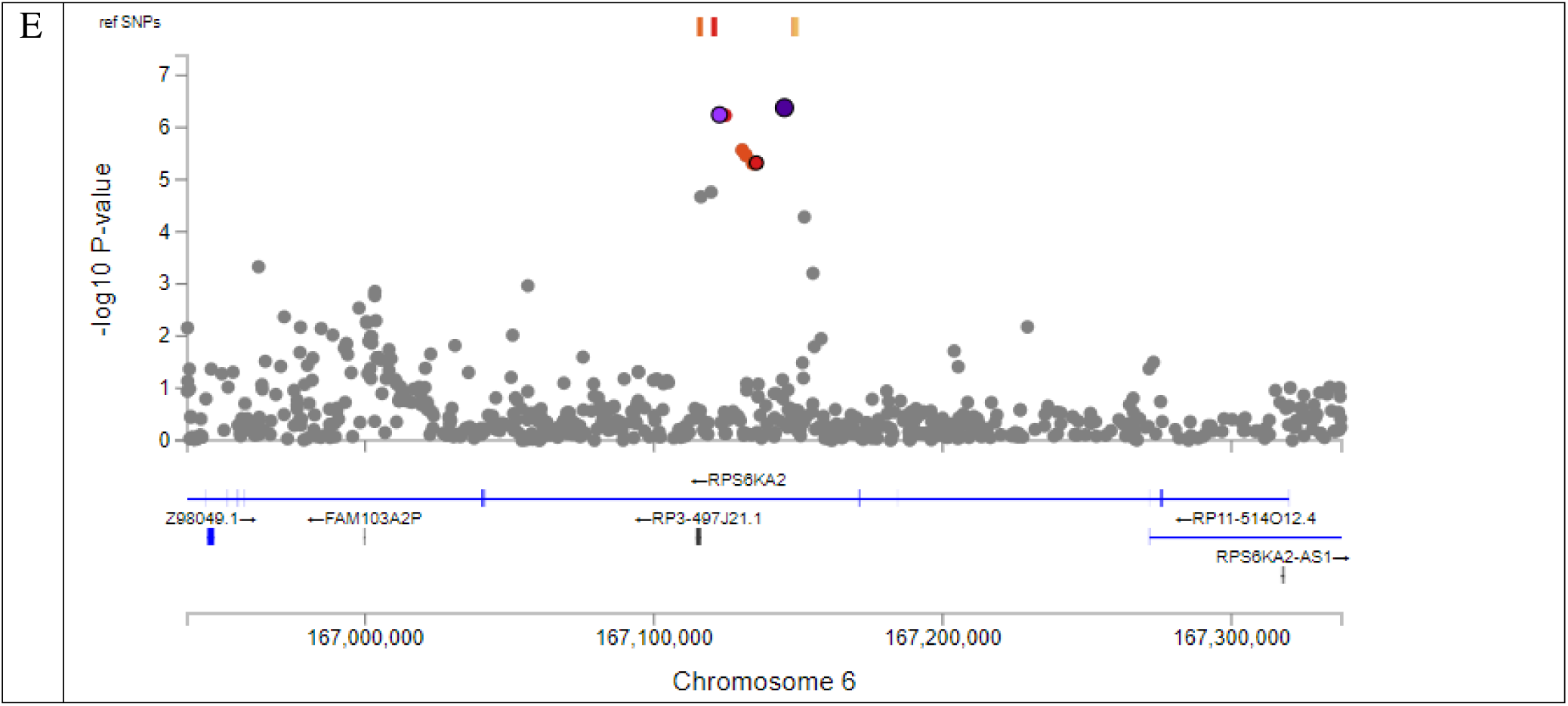
Regional plot. (A) rs75615271 (1:212101892:A:G) for global ILAS. (B) rs75615271 (1:212101892) and (C) rs79148417 (1:67671368) for anterior ILAS. (D) rs62238282 (3:872733) and (E) rs78189747 (6:167145219) for posterior ILAS.

**Figure 5.**
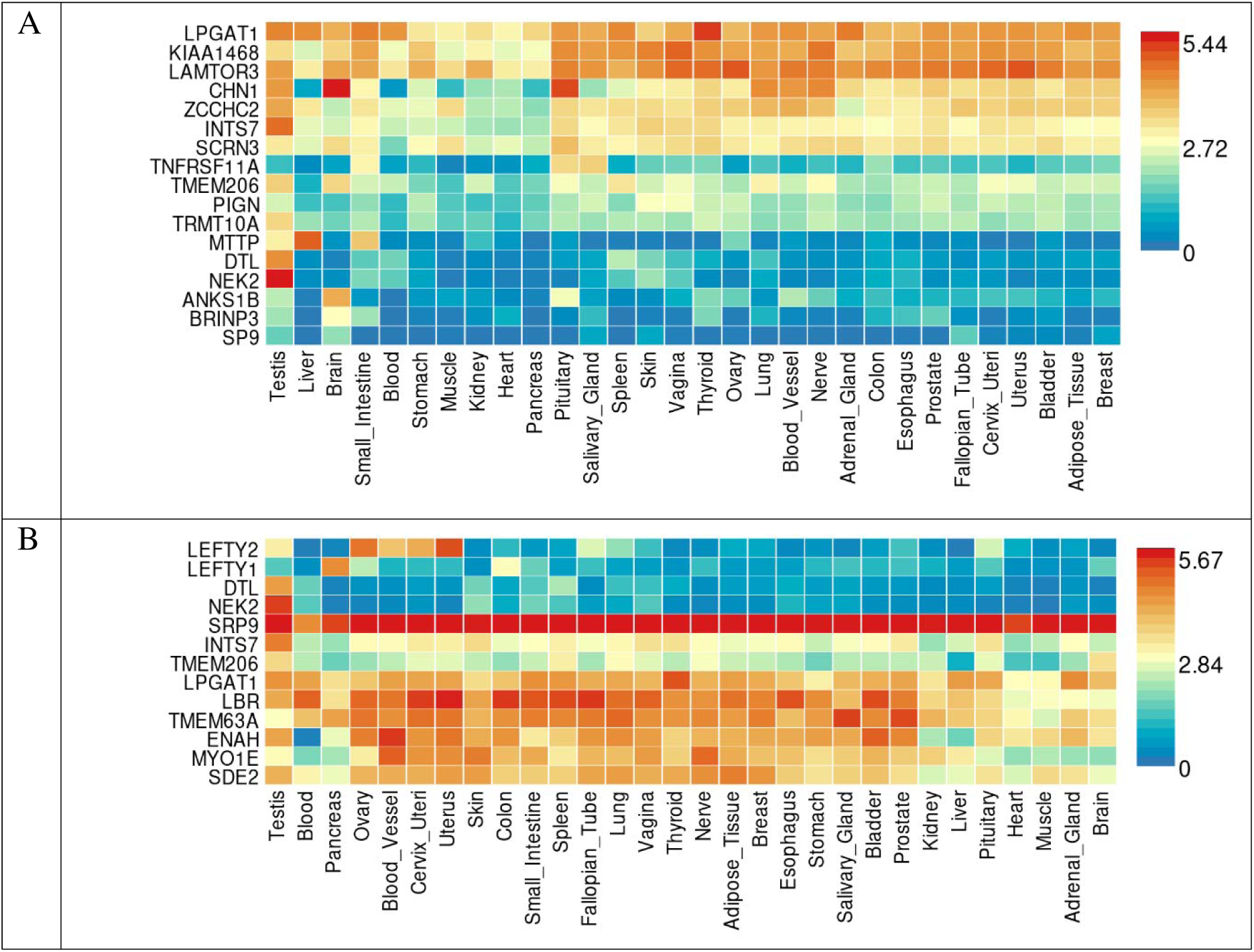
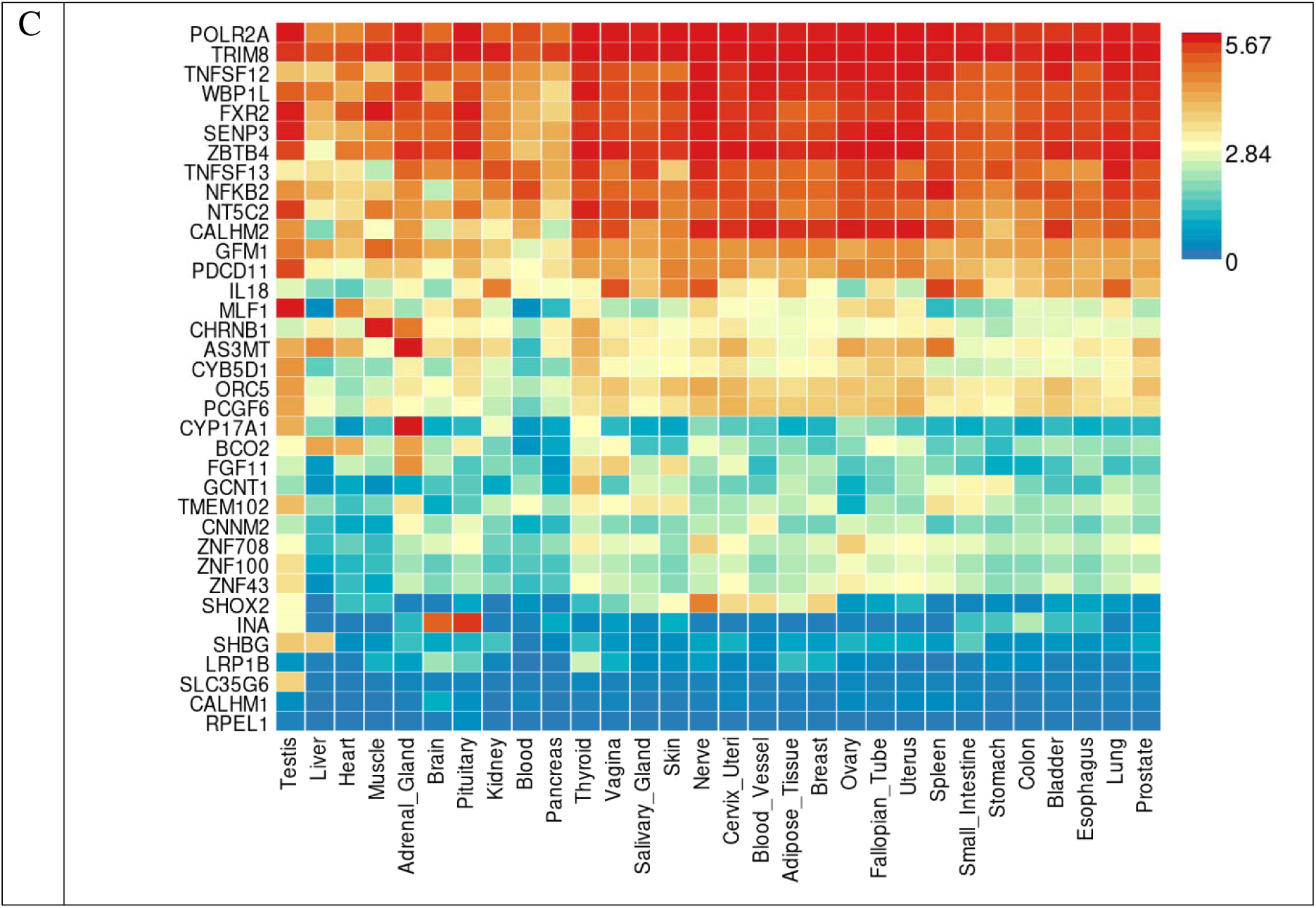
Gene expression heatmap in GTEx v8 dataset 30 general tissue types. (A) Global ILAS. (B) Anterior ILAS. (C) Posterior ILAS.

**Figure 6.**
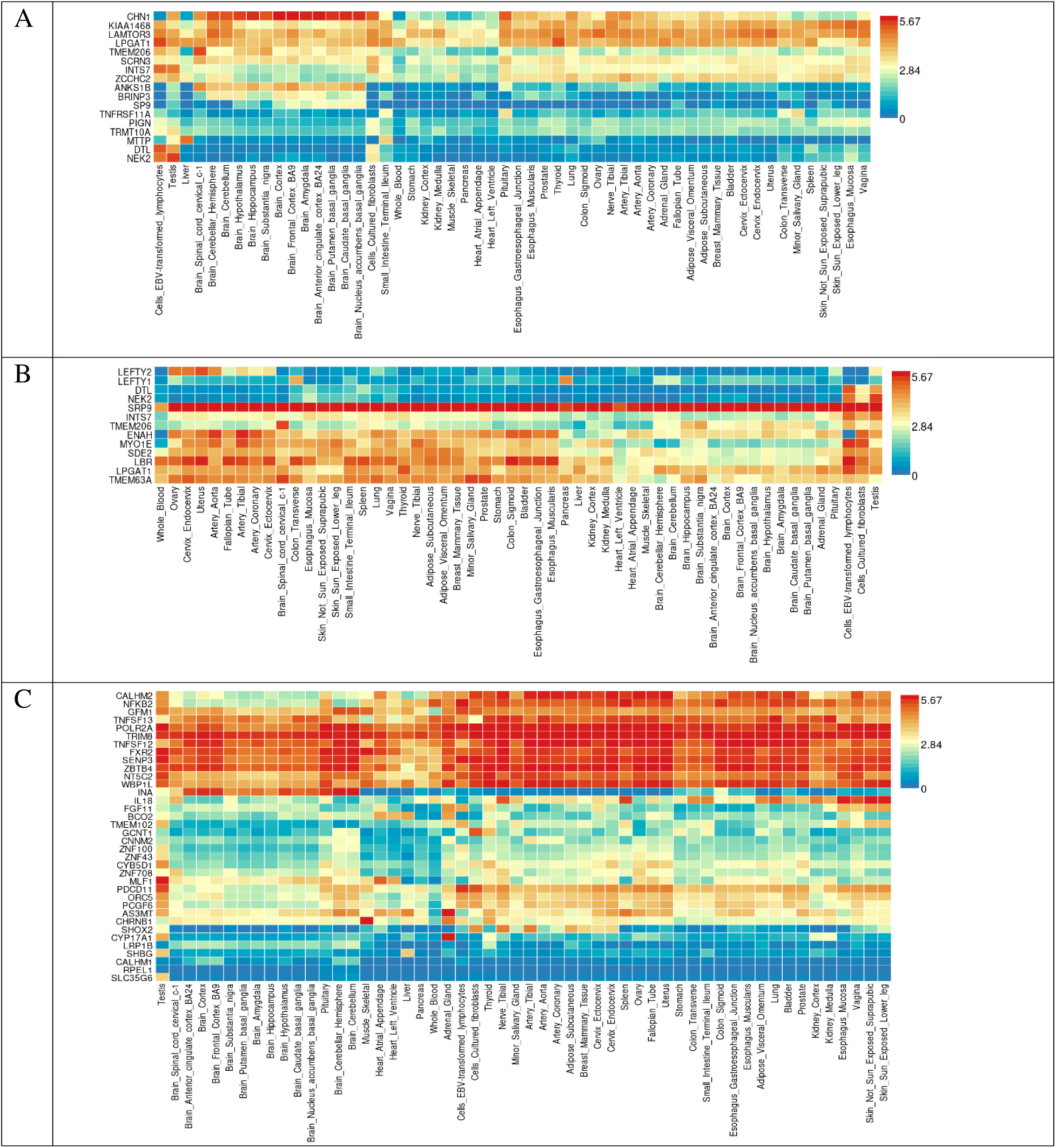
Gene expression heatmap in the GTEx v8 dataset 54 tissue types. (A) Global ILAS. (B) Anterior ILAS. (C) Posterior ILAS.

**Table 2.**
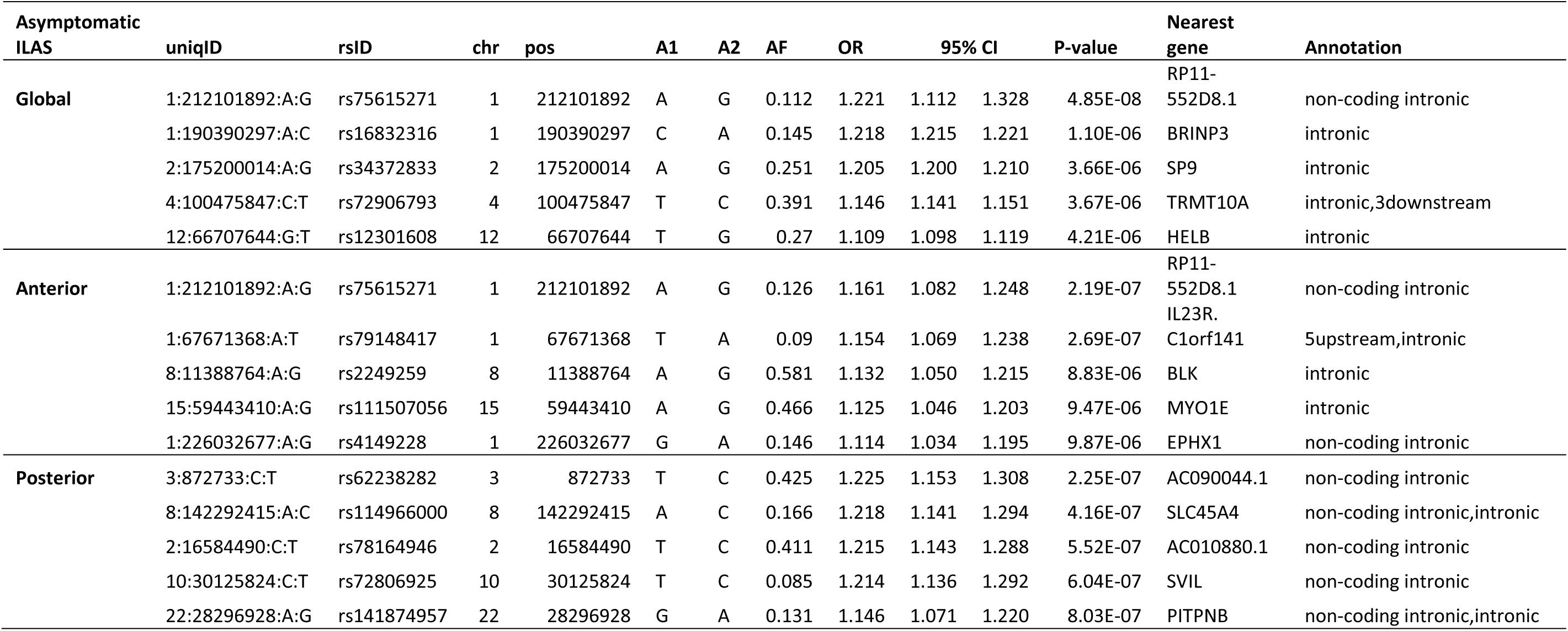
Variants (P <5×10−8) associated with ILAS.

### Gene-based Association Analysis and Gene-set Enrichment

We performed Gene-based association test and did not observe gene-based genome-wide significant associations with asymptomatic ILAS (Figure 3). The top gene was *AC018470.1* (*P*=2.02×10^−5^) for global ILAS, *C15ORF31* (*P*=3.19×10^−5^) for anterior ILAS, and *RORC* (*P*=5.20×10^−5^) for posterior ILAS, respectively. All candidate SNPs, which are in LD of any independent lead SNPs, were mapped to genes. Figure 4 shows the regional plot of top SNPs in global, anterior and posterior ILAS. Based on pLI score and the non-coding residual variation intolerance score, the most intolerant genes were *PIGN* for global ILAS, *SDE2* for anterior ILAS and *BCO2* for posterior ILAS (Table 3). The MAGMA gene-based association analysis identified one gene-set associated with anterior ILAS (Bonferroni adjusted *P*=0.035) and posterior ILAS (Bonferroni adjusted *P*=0.033), respectively (Table 4). These gene-sets were KEGG non-homologous end-joining pathway (ko03450) associated with anterior ILAS and microtubule bundle formation (GO:0005879) associated with posterior ILAS (Table 5). Genes mapped to candidate SNPs were further investigated for gene-set enrichment and functional consequences against reference panels (Table 6). We found a gene set of global ILAS and anterior ILAS that was enriched in chr1q32 region, including *NEK2*, *LPGAT1*, *INTS7*, *DTL*, and *TMEM206*. Additionally, a gene set was enriched in chr10q24 in posterior ILAS (*P*=2.48x10^-20^). Furthermore, we identified one Hispanic-specific gene(*P2RX5*) related to anterior ILAS, and one Asian specific gene (*TMPRSS7*) related to posterior ILAS (Supplemental Figure 7-9). The MAGMA gene-based association analysis identified two gene-sets associated with Hispanic specific anterior ILAS (positive regulation of immature T cell proliferation, adjusted *P*=0.002; T cell activation, adjusted *P*=0.032); and one gene-set (Reactome SARS-CoV-1 infection pathway, adjusted *P*=0.008) associated with White-specific posterior ILAS (Supplemental Table 3 and 4).

**Figure 7.**
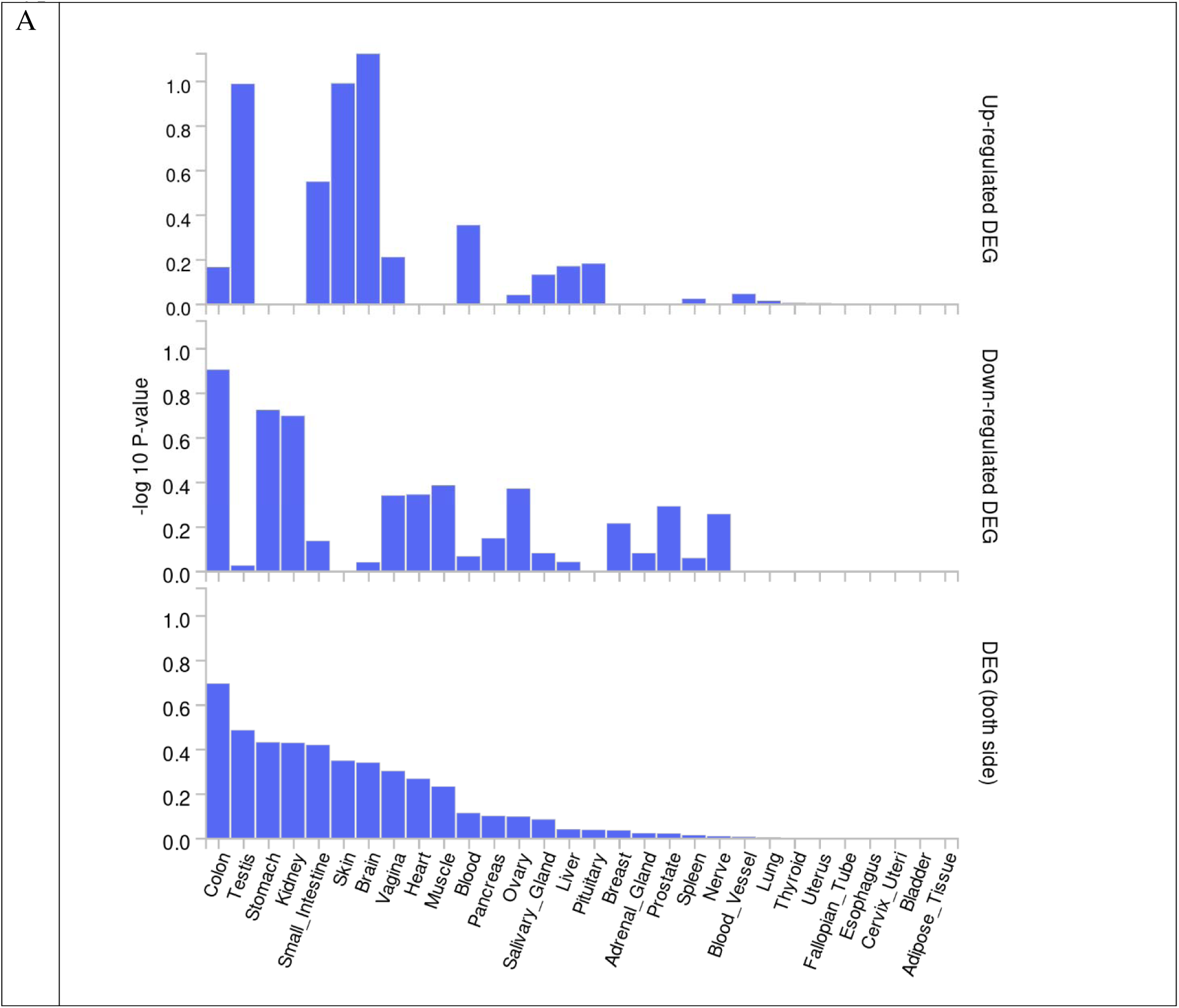
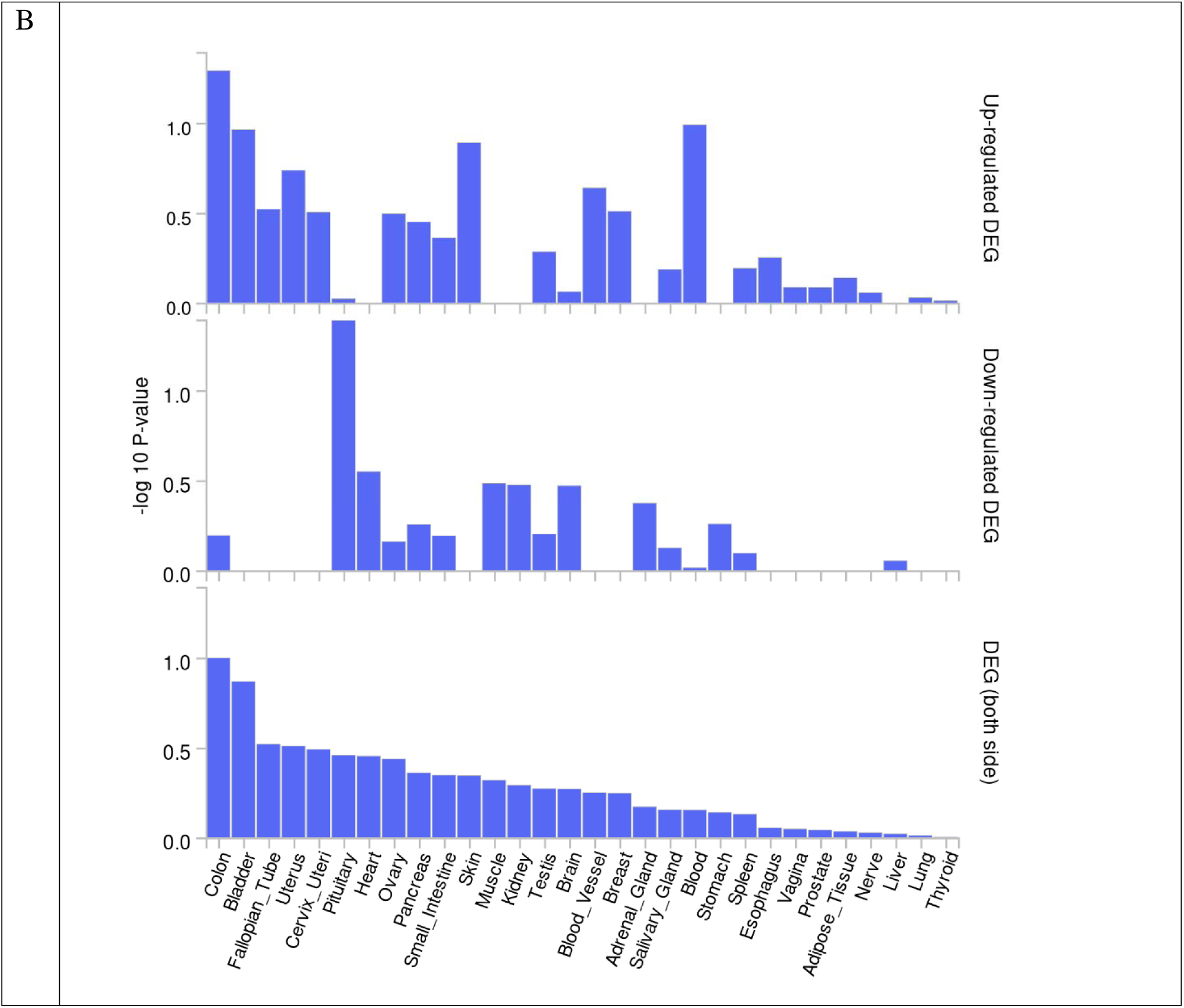
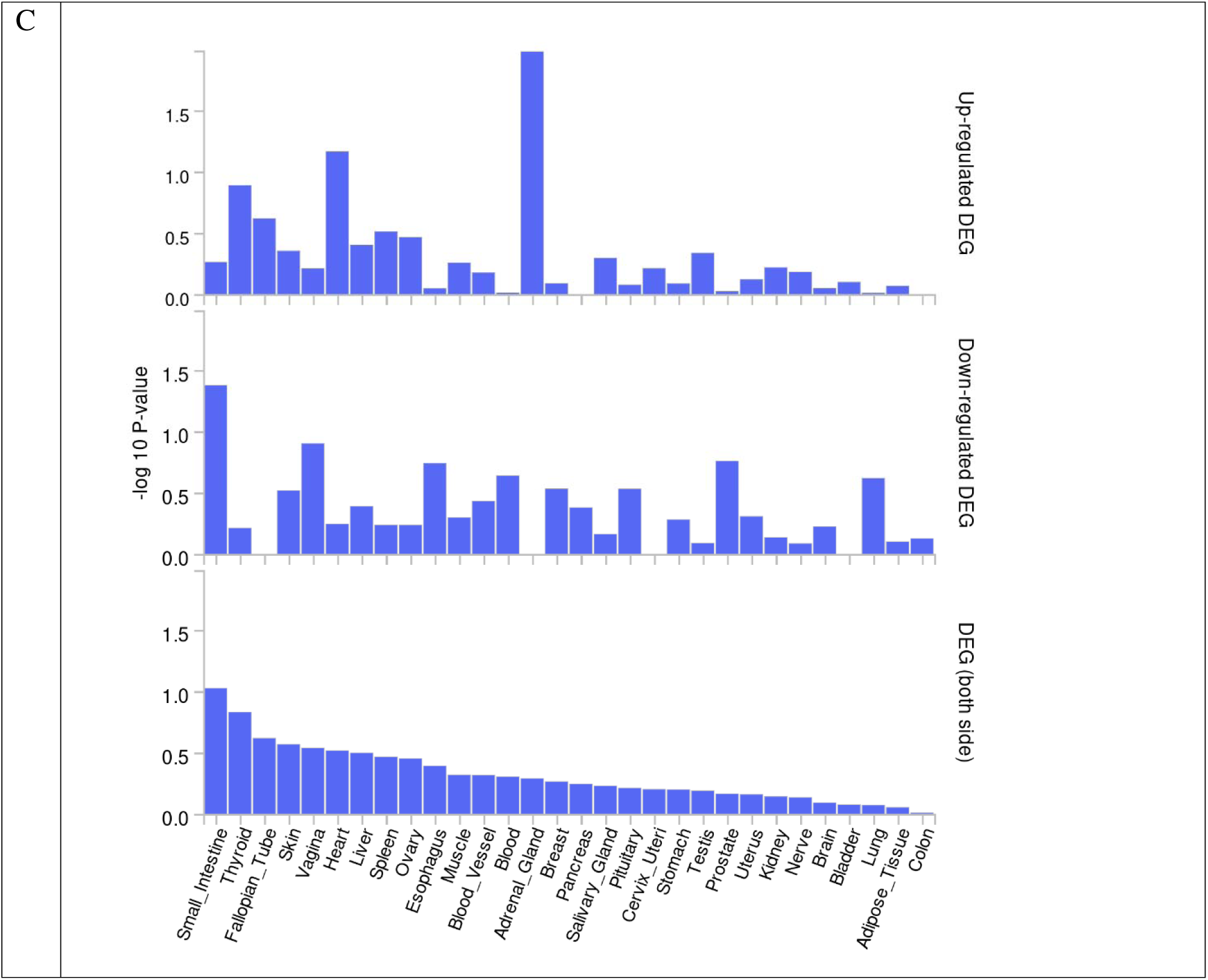
Enrichment test of differentially expressed genes in GTEx v8 dataset 30 general tissue types. (A) Global ILAS. (B) Anterior ILAS. (C) Posterior ILAS.

**Figure 8.**
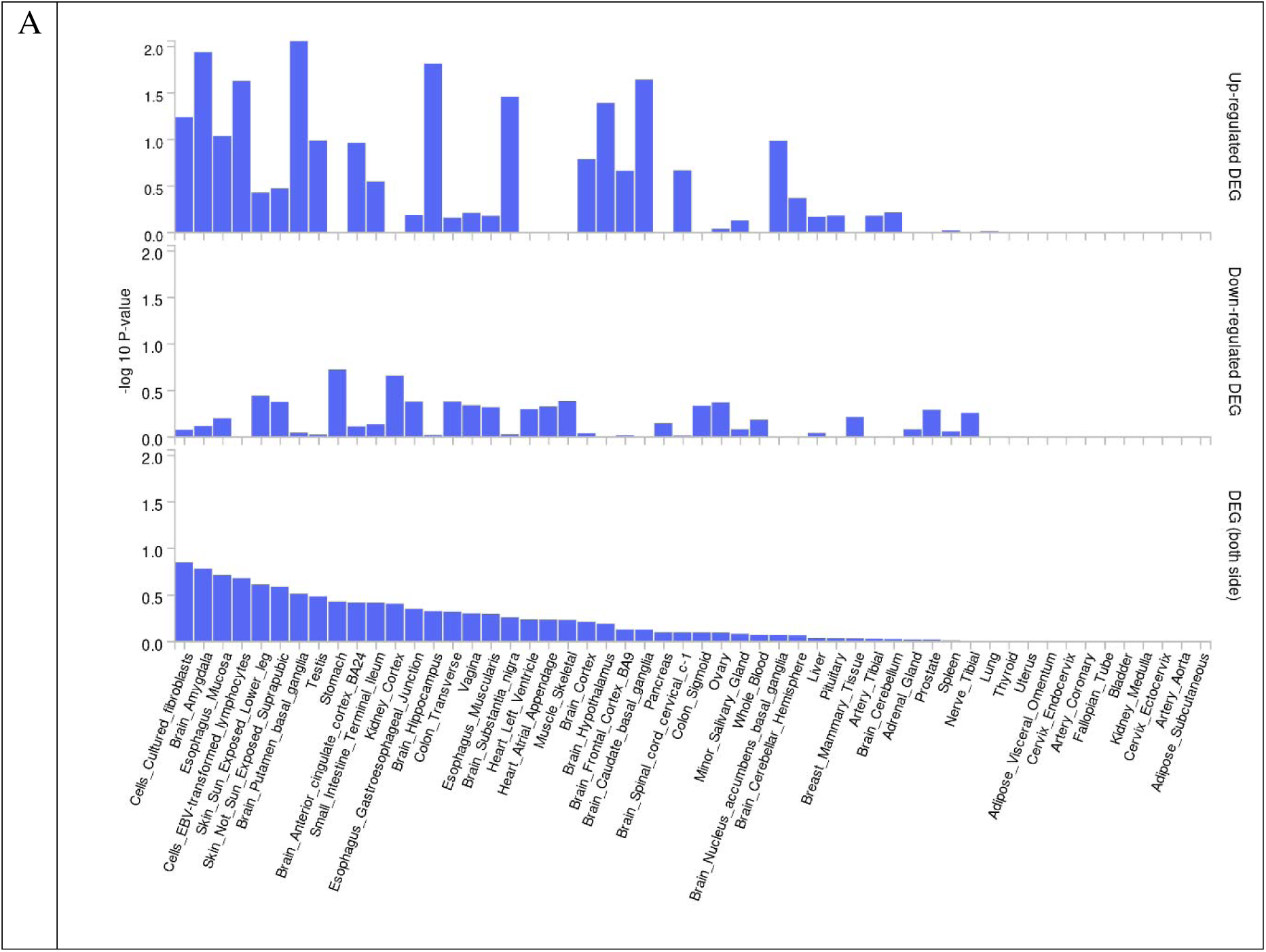
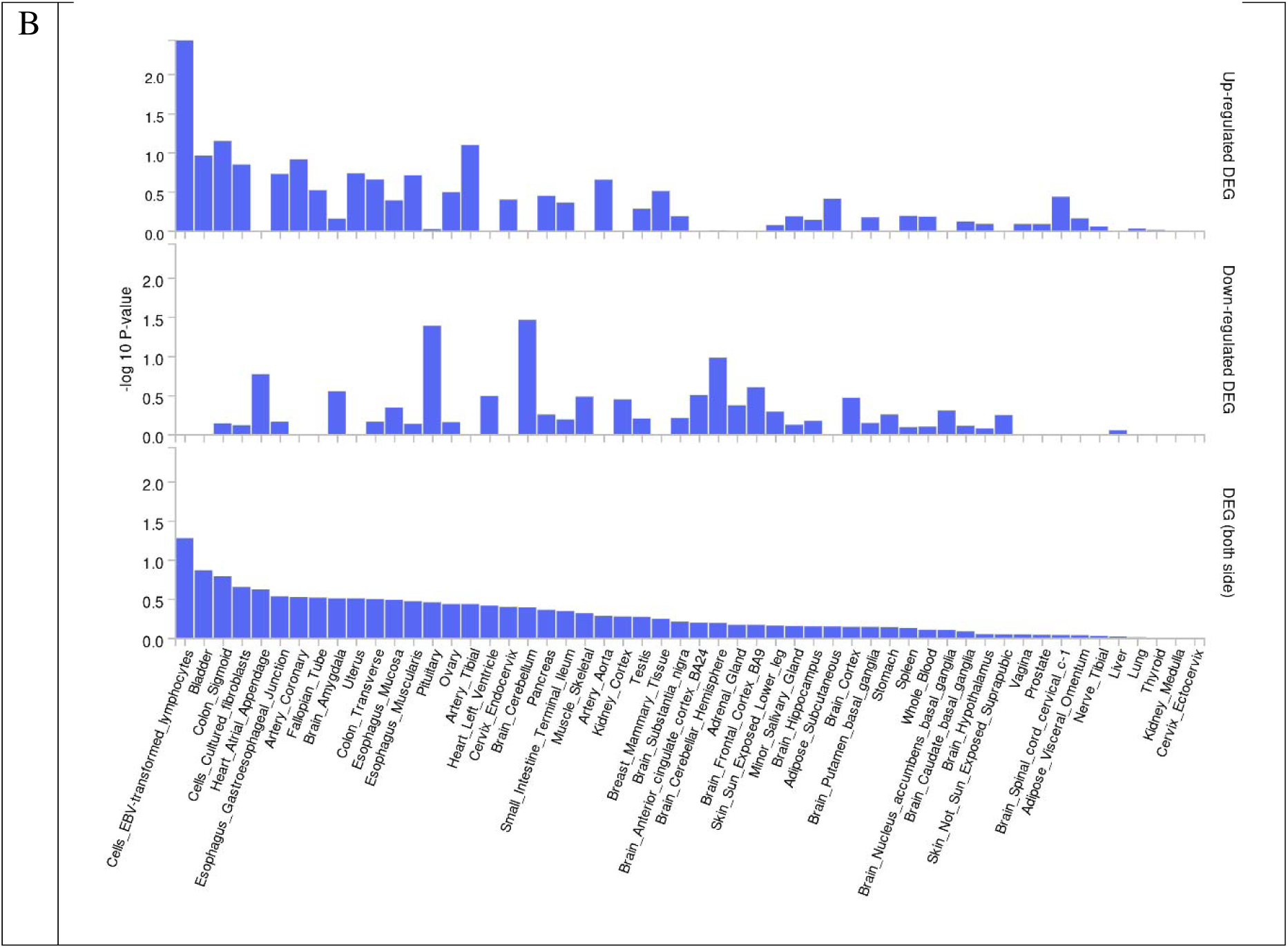
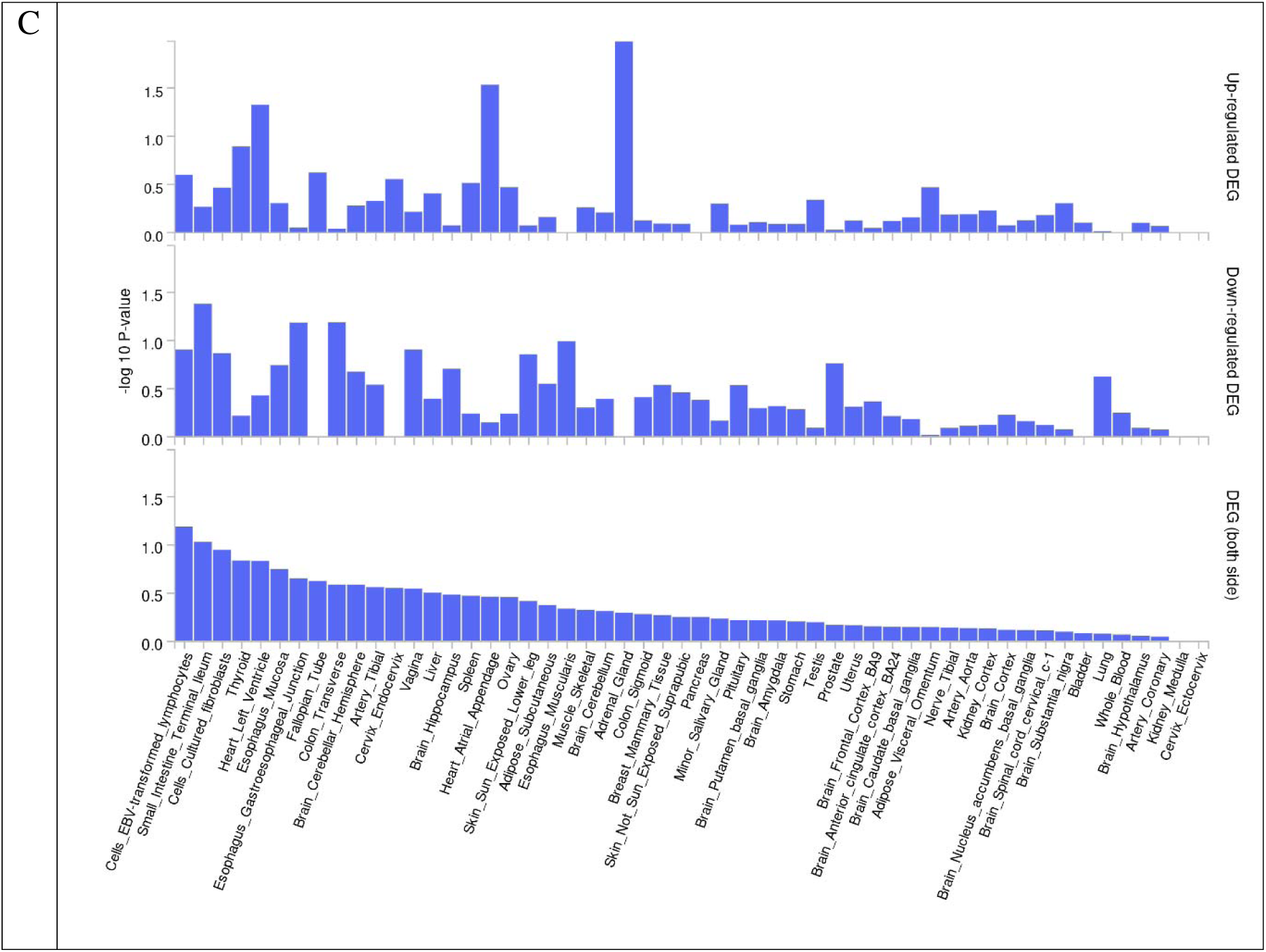
Enrichment test of differentially expressed genes in GTEx v8 dataset 54 tissue types. (A) Global ILAS. (B) Anterior ILAS. (C) Posterior ILAS.

**Figure 9.**
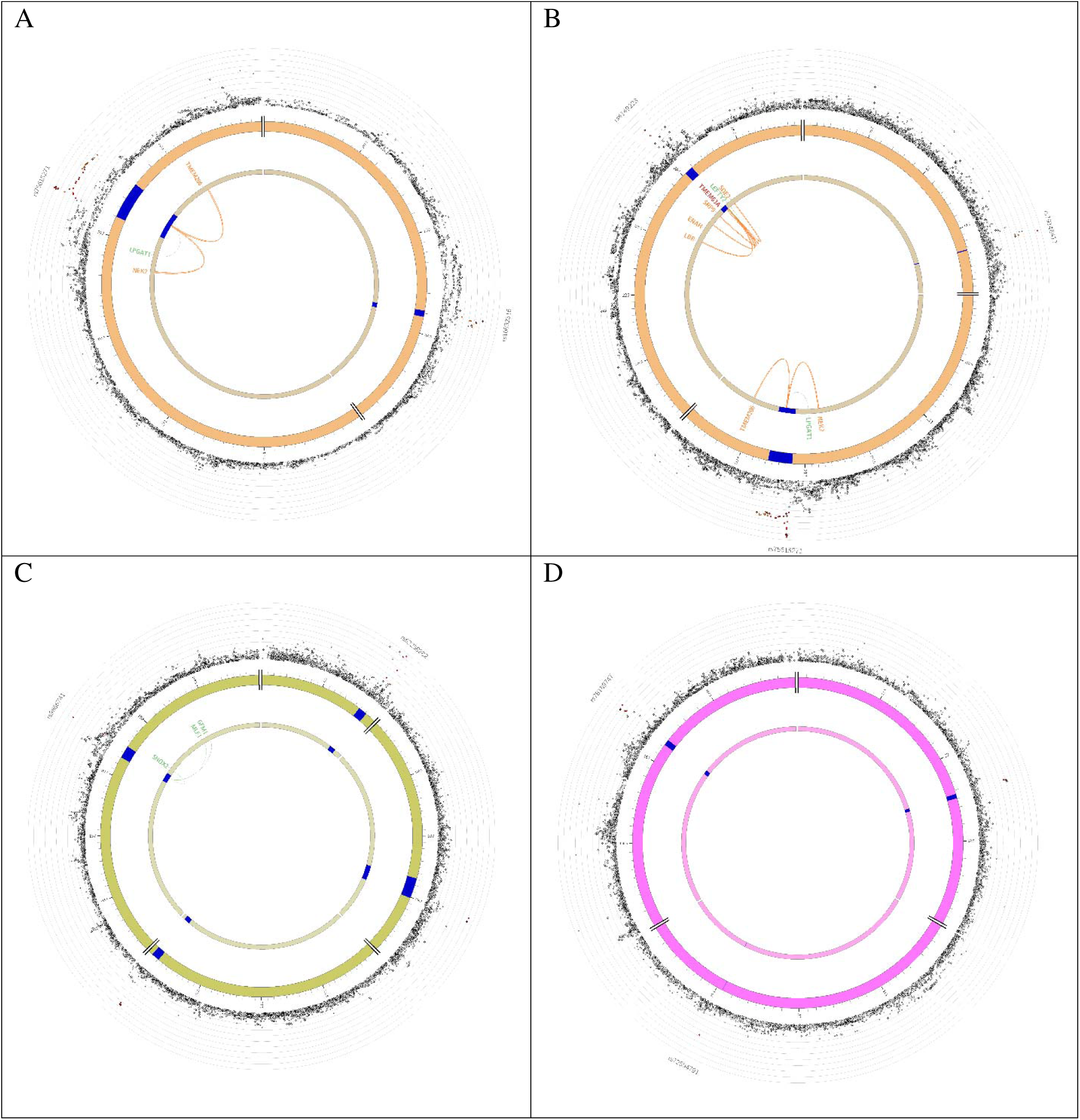
Circos plot for the genomewide level locus. Chromatin interaction between genomic risk locus (blue arc) and genes were showed by orange line. The green color line linked the top SNP and the eQTL genes. (A) Chromosome 1 top SNPs for Global ILAS. (B) Chromosome 1 rs75615271 top SNP for Anterior ILAS. (C) Chromosome 3 top SNP for Posterior ILAS. (D) Chromosome 6 top SNP for Posterior ILAS.

**Table 3.**
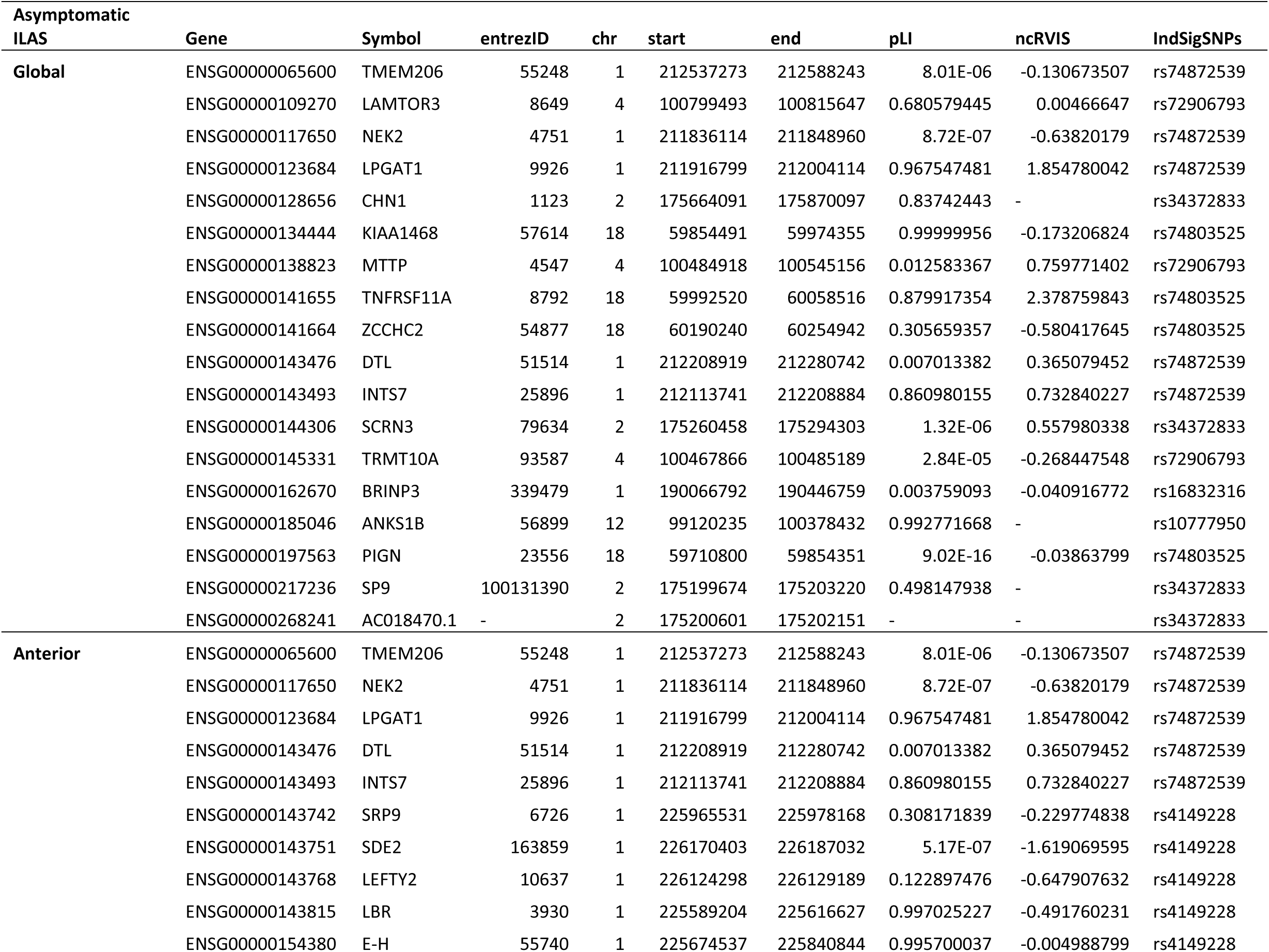
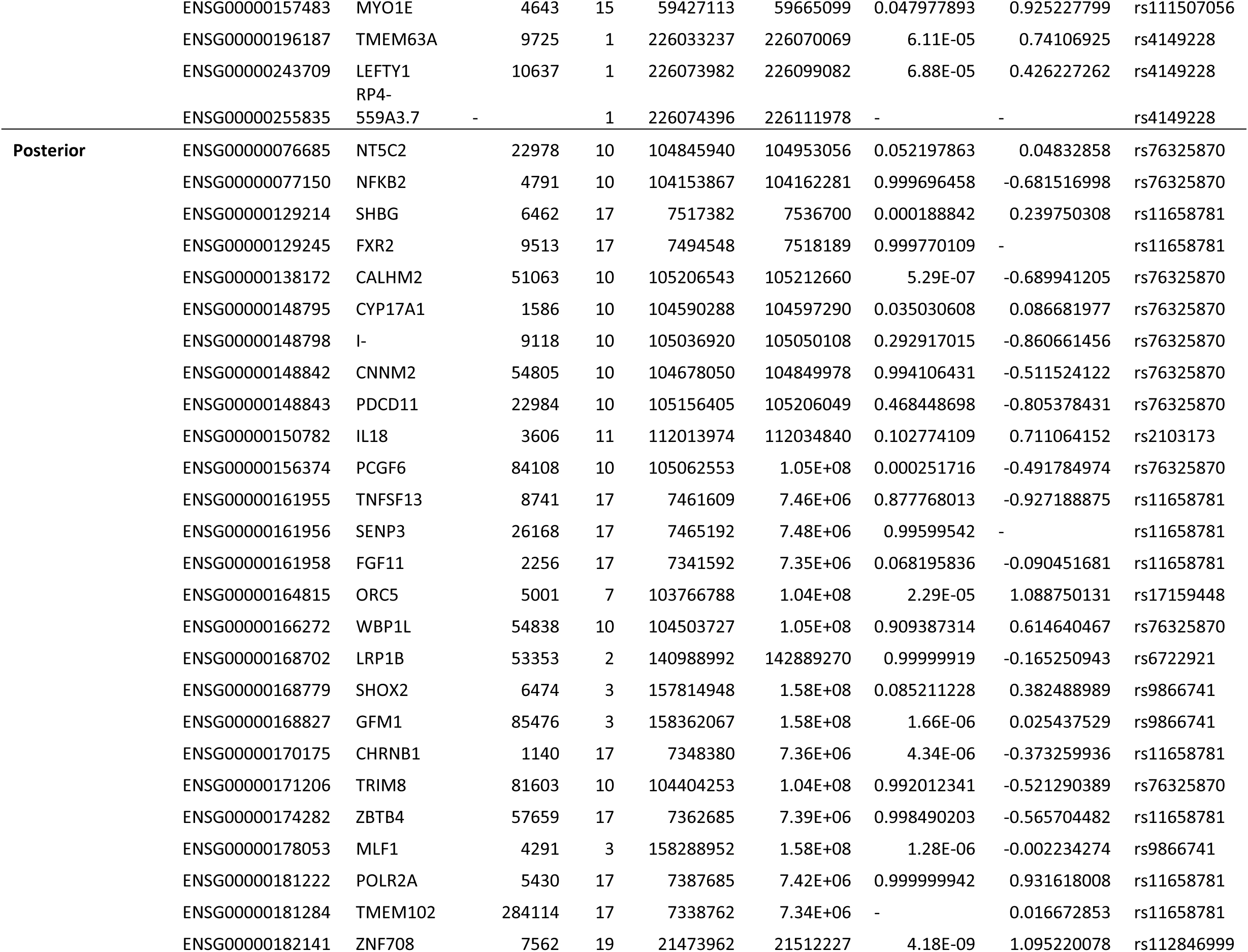
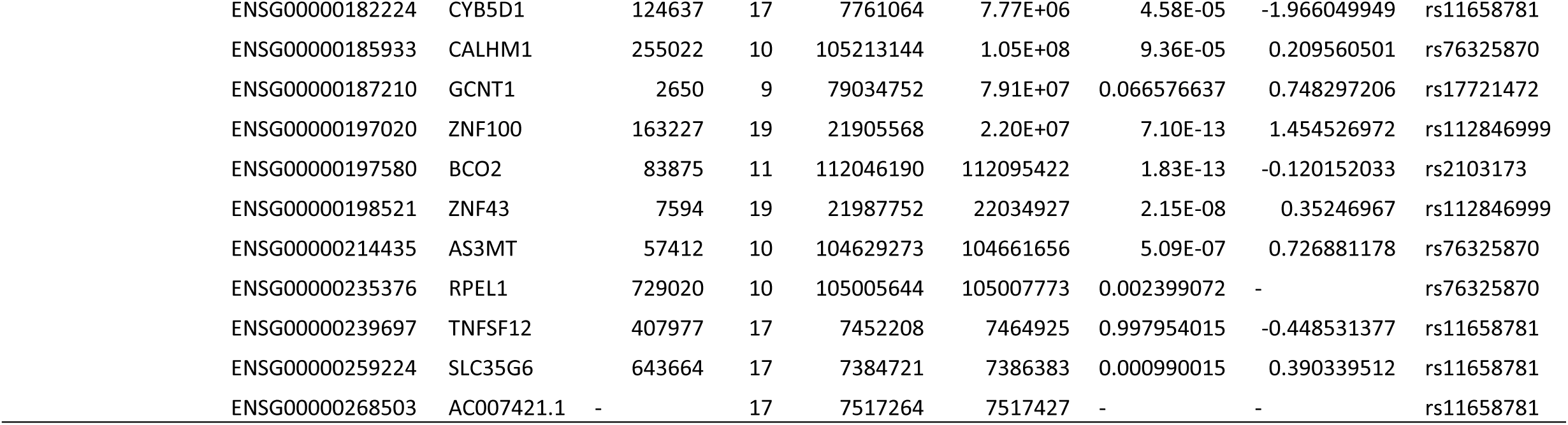
Mapped genes.

**Table 4.**
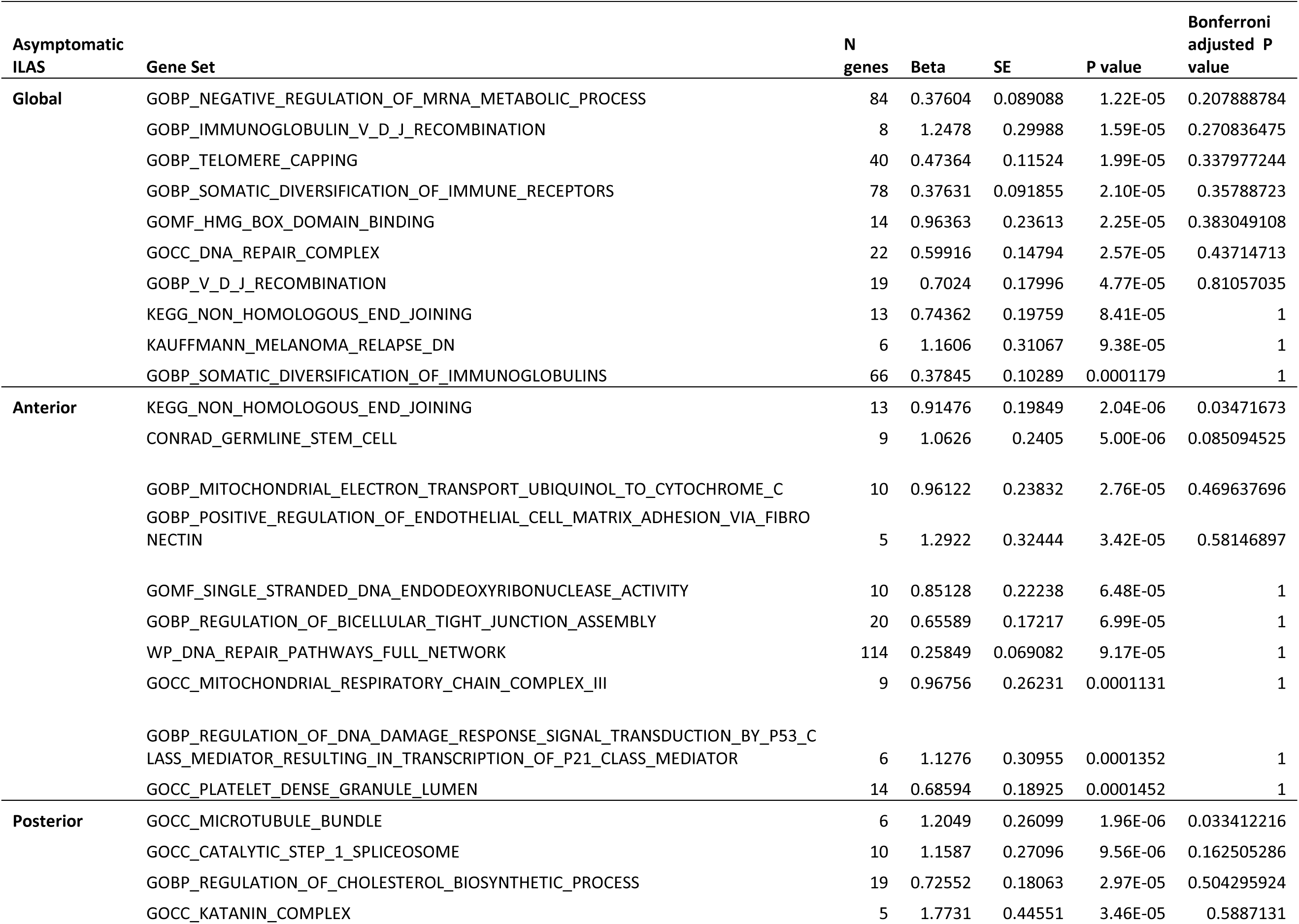
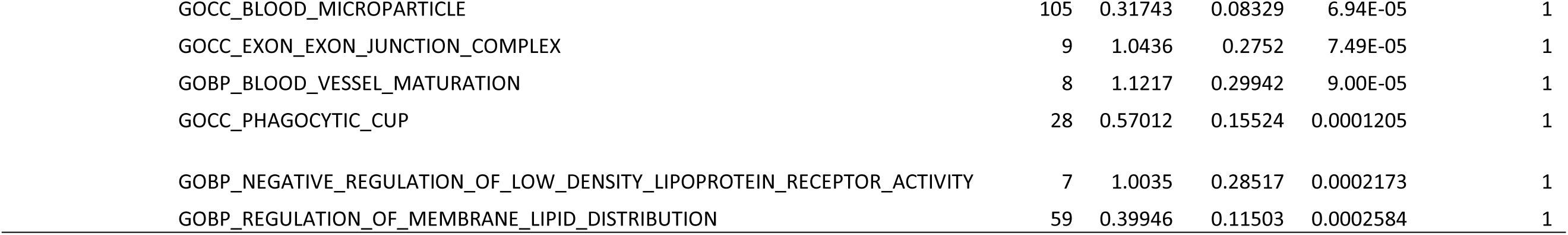
MAGMA pathway analysis.

| Asymptomatic<br>ILAS | Gene Set | N<br>genes | Beta | SE | P value | Bonferroni<br>adjusted P<br>value |
| --- | --- | --- | --- | --- | --- | --- |
| Global | GOBP_NEGATIVE_REGULATION_OF_MRNA_METABOLIC_PROCESS | 84 | 0.37604 | 0.089088 | 1.22E-05 | 0.207888784 |
|  | GOBP_IMMUNOGLOBULIN_V_D_J_RECOMBINATION | 8 | 1.2478 | 0.29988 | 1.59E-05 | 0.270836475 |
|  | GOBP_TELOMERE_CAPPING | 40 | 0.47364 | 0.11524 | 1.99E-05 | 0.337977244 |
|  | GOBP_SOMATIC_DIVERSIFICATION_OF_IMMUNE_RECEPTORS | 78 | 0.37631 | 0.091855 | 2.10E-05 | 0.35788723 |
|  | GOMF_HMG_BOX_DOMAIN_BINDING | 14 | 0.96363 | 0.23613 | 2.25E-05 | 0.383049108 |
|  | GOCC_DNA_REPAIR_COMPLEX | 22 | 0.59916 | 0.14794 | 2.57E-05 | 0.43714713 |
|  | GOBP_V_D_J_RECOMBINATION | 19 | 0.7024 | 0.17996 | 4.77E-05 | 0.81057035 |
|  | KEGG_NON_HOMOLOGOUS_END_JOINING | 13 | 0.74362 | 0.19759 | 8.41E-05 | 1 |
|  | KAUFFMANN_MELANOMA_RELAPSE_DN | 6 | 1.1606 | 0.31067 | 9.38E-05 | 1 |
|  | GOBP_SOMATIC_DIVERSIFICATION_OF_IMMUNOGLOBULINS | 66 | 0.37845 | 0.10289 | 0.0001179 | 1 |
| Anterior | KEGG_NON_HOMOLOGOUS_END_JOINING | 13 | 0.91476 | 0.19849 | 2.04E-06 | 0.03471673 |
|  | CONRAD_GERMLINE_STEM_CELL | 9 | 1.0626 | 0.2405 | 5.00E-06 | 0.085094525 |
|  | GOBP_MITOCHONDRIAL_ELECTRON_TRANSPORT_UBIQUINOL_TO_CYTOCHROME_C | 10 | 0.96122 | 0.23832 | 2.76E-05 | 0.469637696 |
|  | GOBP_POSITIVE_REGULATION_OF_ENDOTHELIAL_CELL_MATRIX_ADHESION_VIA_FIBRO<br>NECTIN | 5 | 1.2922 | 0.32444 | 3.42E-05 | 0.58146897 |
|  | GOMF_SINGLE_STRANDED_DNA_ENDODEOXYRIBONUCLEASE_ACTIVITY | 10 | 0.85128 | 0.22238 | 6.48E-05 | 1 |
|  | GOBP_REGULATION_OF_BICELLULAR_TIGHT_JUNCTION_ASSEMBLY | 20 | 0.65589 | 0.17217 | 6.99E-05 | 1 |
|  | WP_DNA_REPAIR_PATHWAYS_FULL_NETWORK | 114 | 0.25849 | 0.069082 | 9.17E-05 | 1 |
|  | GOCC_MITOCHONDRIAL_RESPIRATORY_CHAIN_COMPLEX_III | 9 | 0.96756 | 0.26231 | 0.0001131 | 1 |
|  | GOBP_REGULATION_OF_DNA_DAMAGE_RESPONSE_SIGNAL_TRANSDUCTION_BY_P53_C<br>LASS_MEDIATOR_RESULTING_IN_TRANSCRIPTION_OF_P21_CLASS_MEDIATOR | 6 | 1.1276 | 0.30955 | 0.0001352 | 1 |
|  | GOCC_PLATELET_DENSE_GRANULE_LUMEN | 14 | 0.68594 | 0.18925 | 0.0001452 | 1 |
| Posterior | GOCC_MICROTUBULE_BUNDLE | 6 | 1.2049 | 0.26099 | 1.96E-06 | 0.033412216 |
|  | GOCC_CATALYTIC_STEP_1_SPLICEOSOME | 10 | 1.1587 | 0.27096 | 9.56E-06 | 0.162505286 |
|  | GOBP_REGULATION_OF_CHOLESTEROL_BIOSYNTHETIC_PROCESS | 19 | 0.72552 | 0.18063 | 2.97E-05 | 0.504295924 |
|  | GOCC_KATANIN_COMPLEX | 5 | 1.7731 | 0.44551 | 3.46E-05 | 0.5887131 |

|  |  |  |  |  |  |
| --- | --- | --- | --- | --- | --- |
| GOCC_BLOOD_MICROPARTICLE | 105 | 0.31743 | 0.08329 | 6.94E-05 | 1 |
| GOCC_EXON_EXON_JUNCTION_COMPLEX | 9 | 1.0436 | 0.2752 | 7.49E-05 | 1 |
| GOBP_BLOOD_VESSEL_MATURATION | 8 | 1.1217 | 0.29942 | 9.00E-05 | 1 |
| GOCC_PHAGOCYTIC_CUP | 28 | 0.57012 | 0.15524 | 0.0001205 | 1 |
| GOBP_NEGATIVE_REGULATION_OF_LOW_DENSITY_LIPOPROTEIN_RECEPTOR_ACTIVITY | 7 | 1.0035 | 0.28517 | 0.0002173 | 1 |
| GOBP_REGULATION_OF_MEMBRANE_LIPID_DISTRIBUTION | 59 | 0.39946 | 0.11503 | 0.0002584 | 1 |

**Table 5.** Geneset GO analysis.

| GENE | Symbol | CHR | START | STOP | N_SNPS | P |
| --- | --- | --- | --- | --- | --- | --- |
| **KEGG non-homologous end-joining pathway associated with anterior ILAS** |  |  |  |  |  |  |
| ENSG00000079246 | XRCC5 | 2 | 216970187 | 217073026 | 137 | 0.53086 |
| ENSG00000187736 | NHEJ1 | 2 | 219938039 | 220027587 | 96 | 0.12963 |
| ENSG00000152422 | XRCC4 | 5 | 82371317 | 82651606 | 191 | 0.011586 |
| ENSG00000113522 | RAD50 | 5 | 131889711 | 131982313 | 41 | 0.11128 |
| ENSG00000122678 | POLM | 7 | 44109846 | 44124139 | 11 | 0.039869 |
| ENSG00000253729 | PRKDC | 8 | 48683669 | 48874743 | 34 | 0.66164 |
| ENSG00000152457 | DCLRE1C | 10 | 14937358 | 14998431 | 38 | 0.001697 |
| ENSG00000107447 | DNTT | 10 | 98062085 | 98100321 | 34 | 0.49185 |
| ENSG00000166169 | POLL | 10 | 103336639 | 103350027 | 7 | 0.1911 |
| ENSG00000168496 | FEN1 | 11 | 61558109 | 61566716 | 1 | 0.4407 |
| ENSG00000020922 | MRE11A | 11 | 94150895 | 94229074 | 39 | 0.14837 |
| ENSG00000174405 | LIG4 | 13 | 108857787 | 108872716 | 15 | 0.18778 |
| ENSG00000196419 | XRCC6 | 22 | 42015123 | 42062044 | 5 | 0.90636 |
| **GOCC_MICROTUBULE_BUNDLE associated posterior ILAS** |  |  |  |  |  |  |
| ENSG00000171368 | TPPP | 5 | 658883 | 695510 | 36 | 0.13995 |
| ENSG00000137343 | ATAT1 | 6 | 30592619 | 30616600 | 12 | 0.20469 |
| ENSG00000072518 | MARK2 | 11 | 63604400 | 63680491 | 28 | 0.34922 |
| ENSG00000137497 | NUMA1 | 11 | 71711910 | 71793739 | 11 | 0.041269 |
| ENSG00000159713 | TPPP3 | 16 | 67421712 | 67429438 | 1 | 0.7629 |
| ENSG00000168502 | SOGA2 | 18 | 8703659 | 8834776 | 117 | 0.21855 |

**Table 6.**
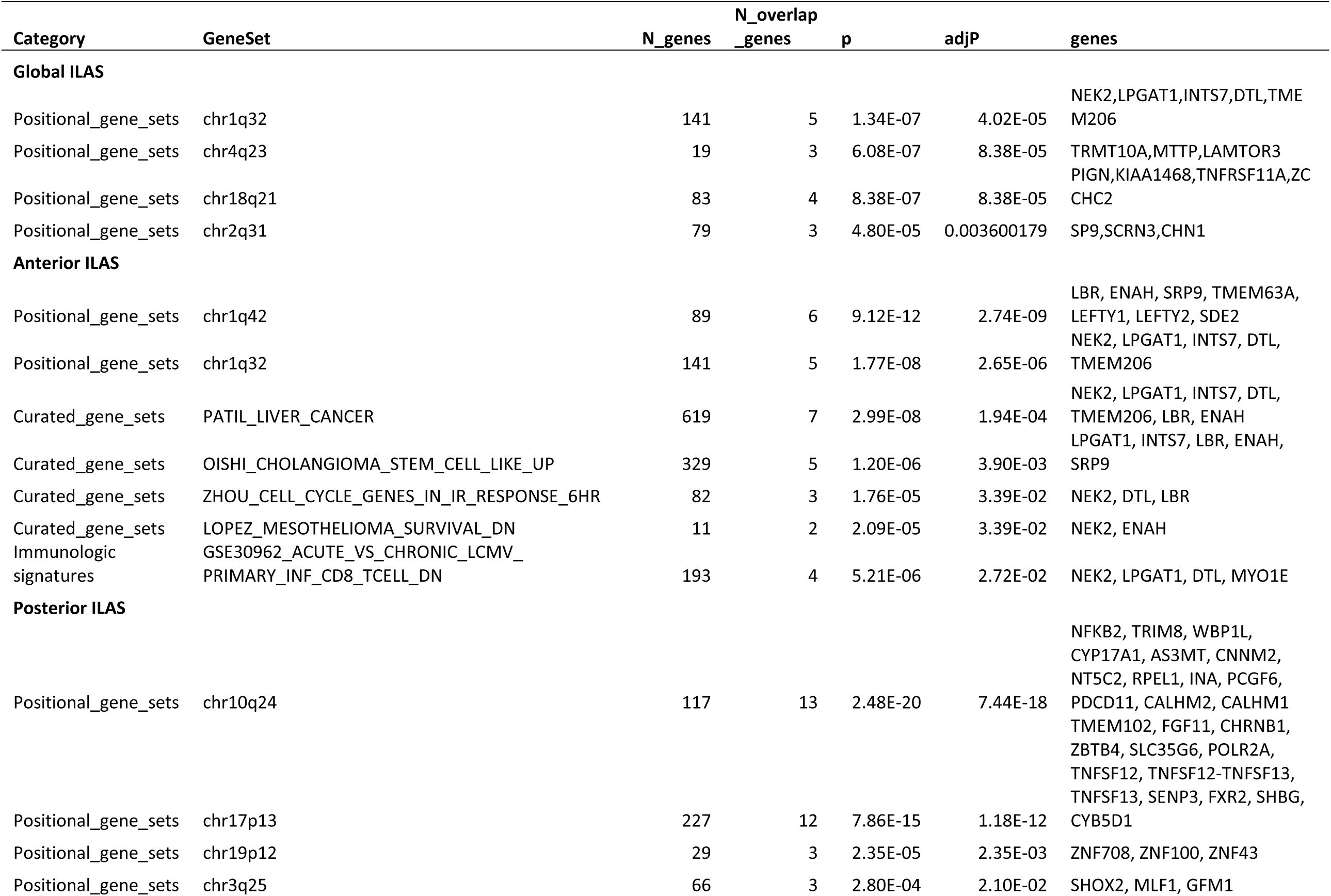
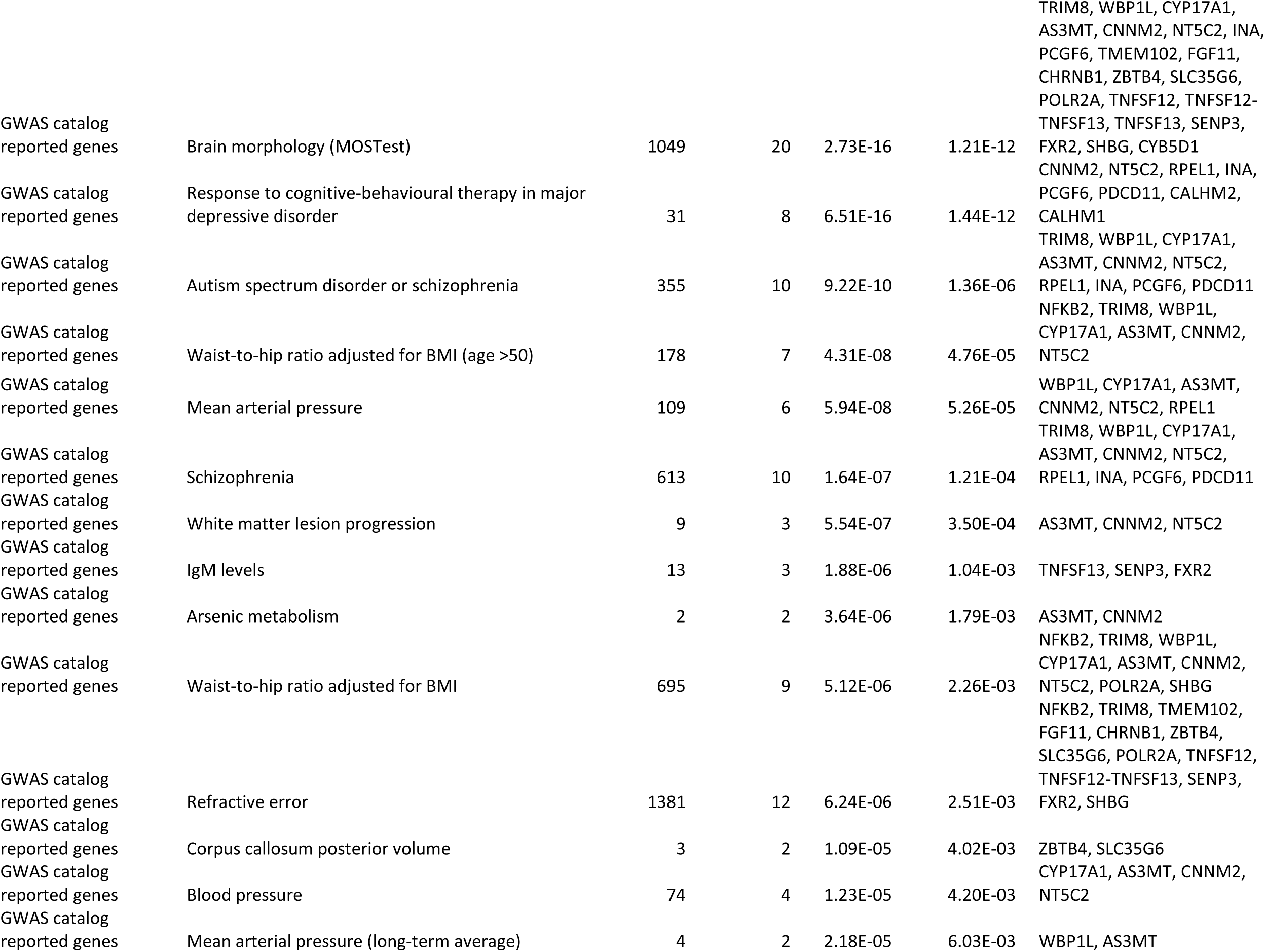
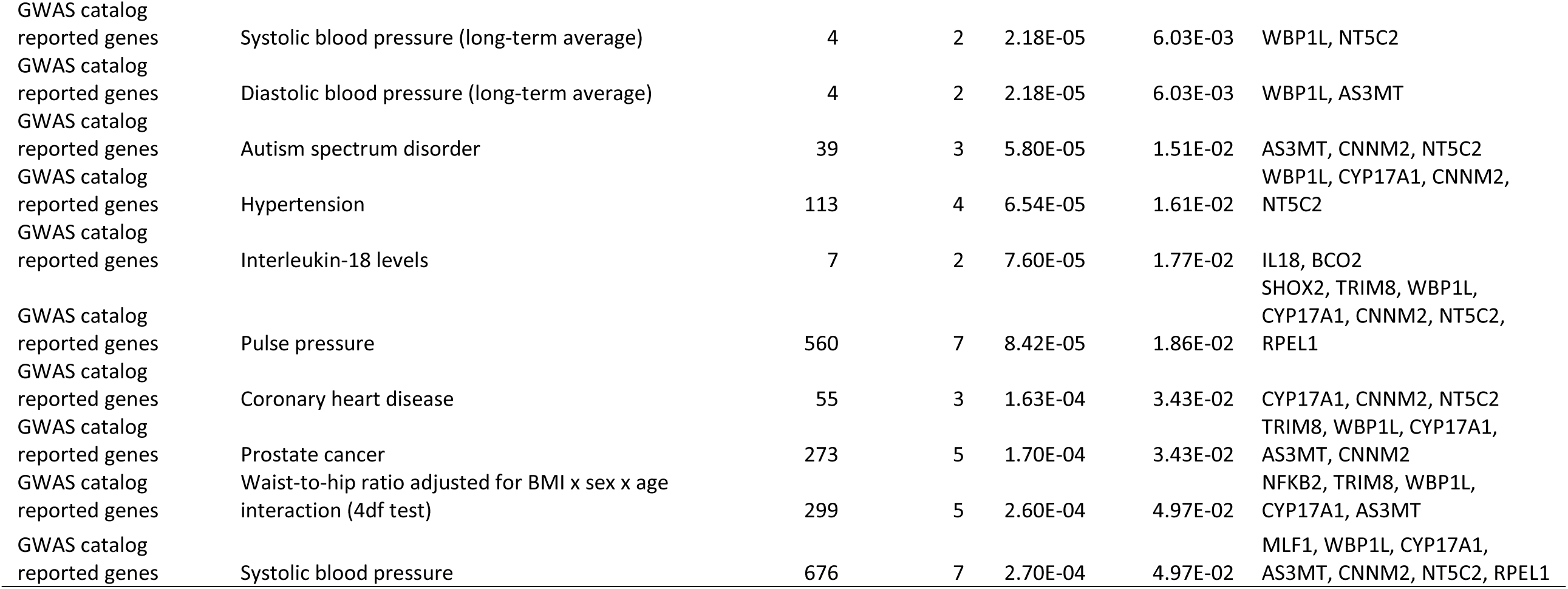
Gene enrichment analysis.

### Tissue-Specific Colocalization Analyses

We performed colocalization analysis for the locus identified in the GWAS analysis and gene-based analysis with gene expression using Genotype-Tissue Expression v8 eQTL data (Table 7). Tissue specific gene expression of 30 general tissue types and 53 specific tissue types of Genotype-Tissue Expression eQTL v8 were presented by heatmap (Figure 5 and 6). In the analysis of 30 general tissues, we identified that *CHN1* associated with global ILAS had higher mRNA expression in the brain tissue (Figure 5). We did not observe the differentially expressed genes based on the enrichment test (Figure 7 and 8).

**Table 7.**
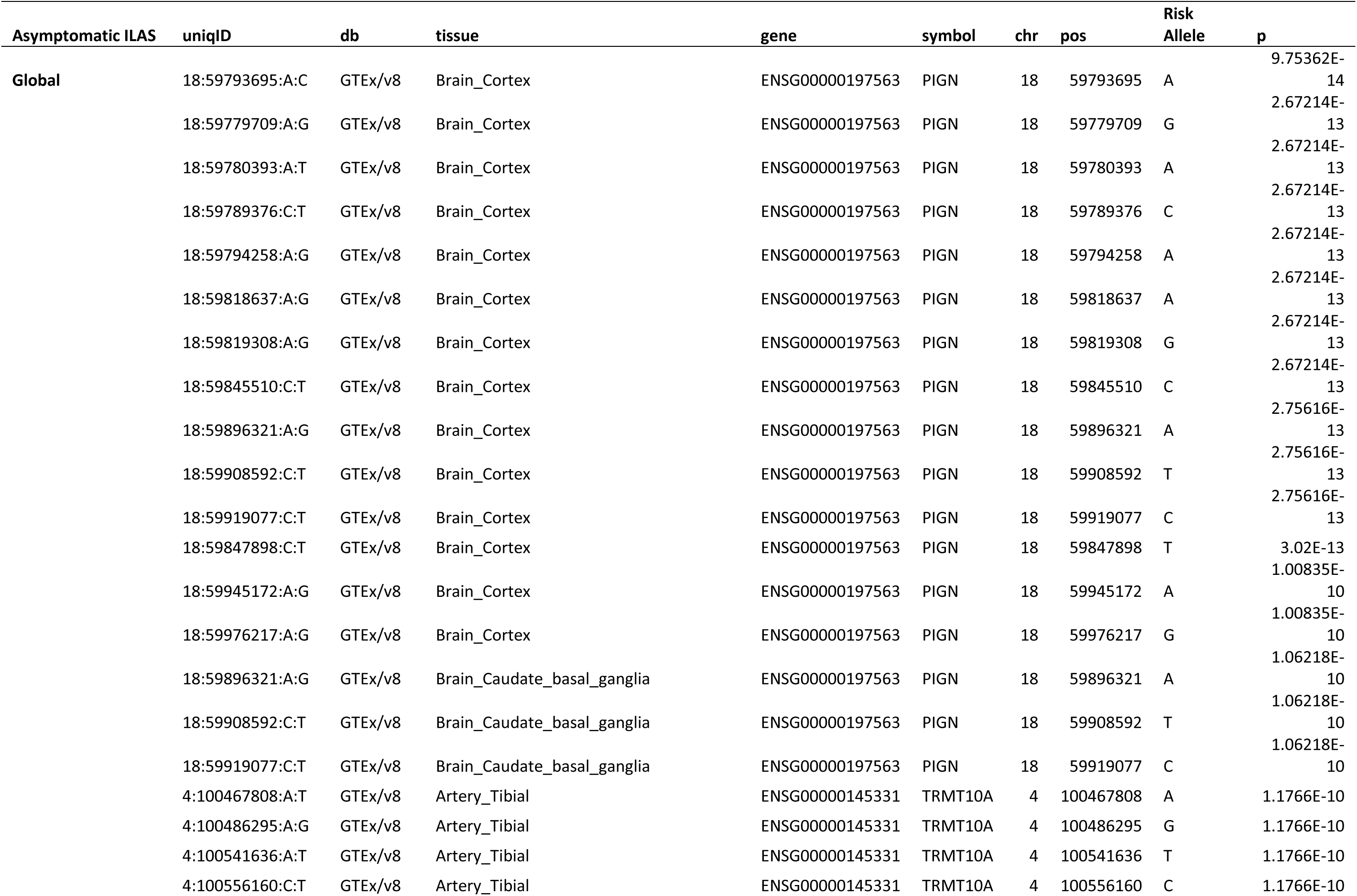
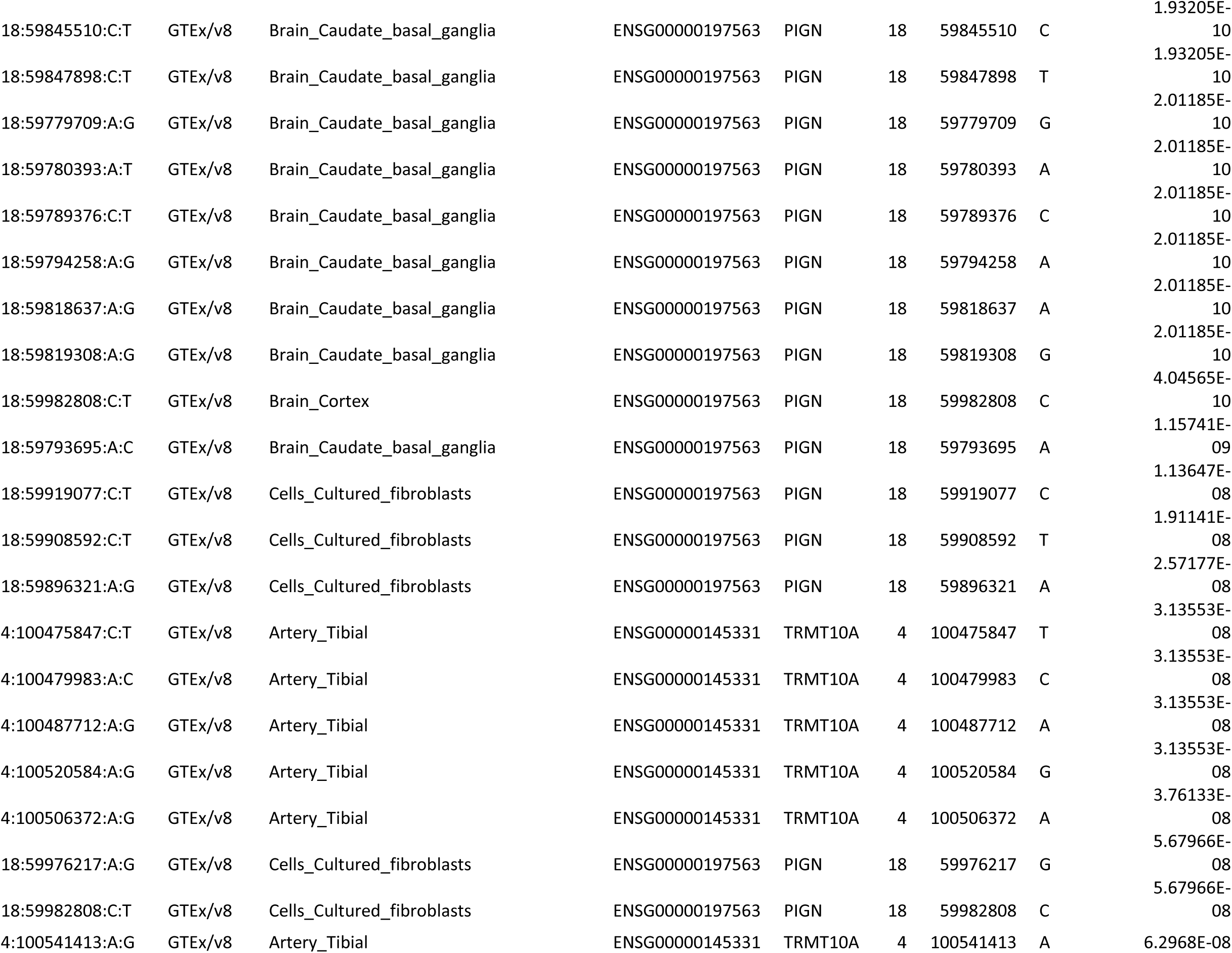
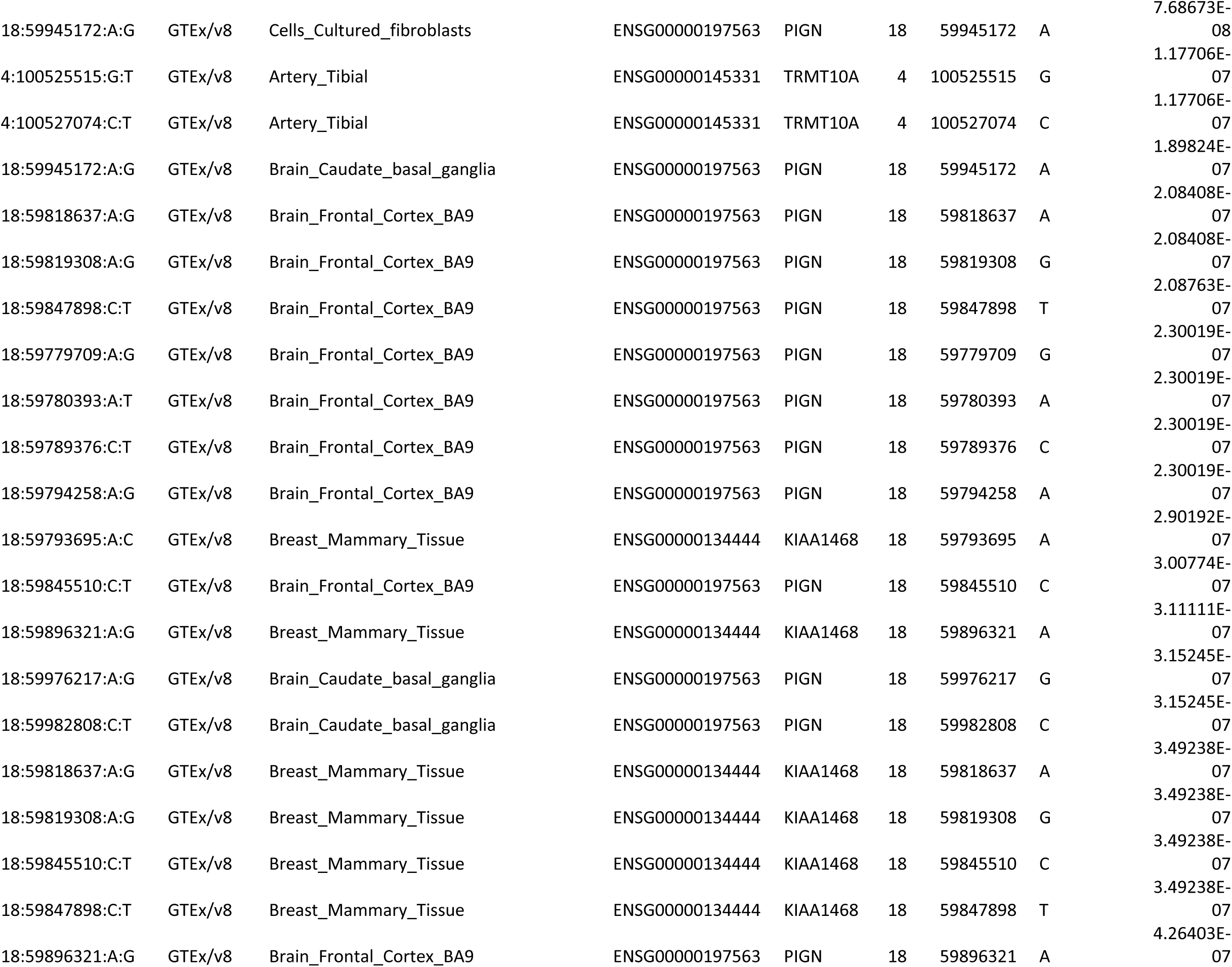
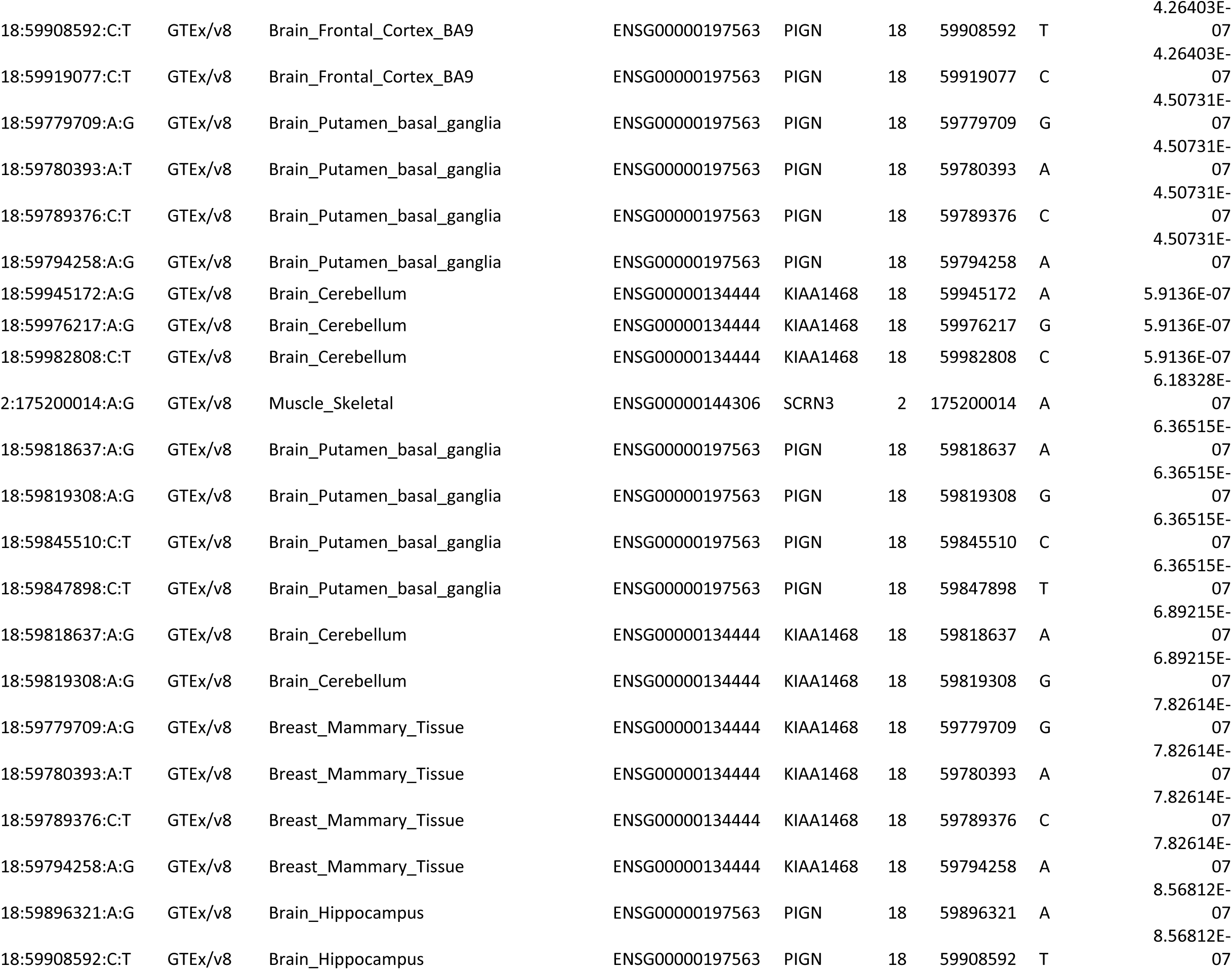
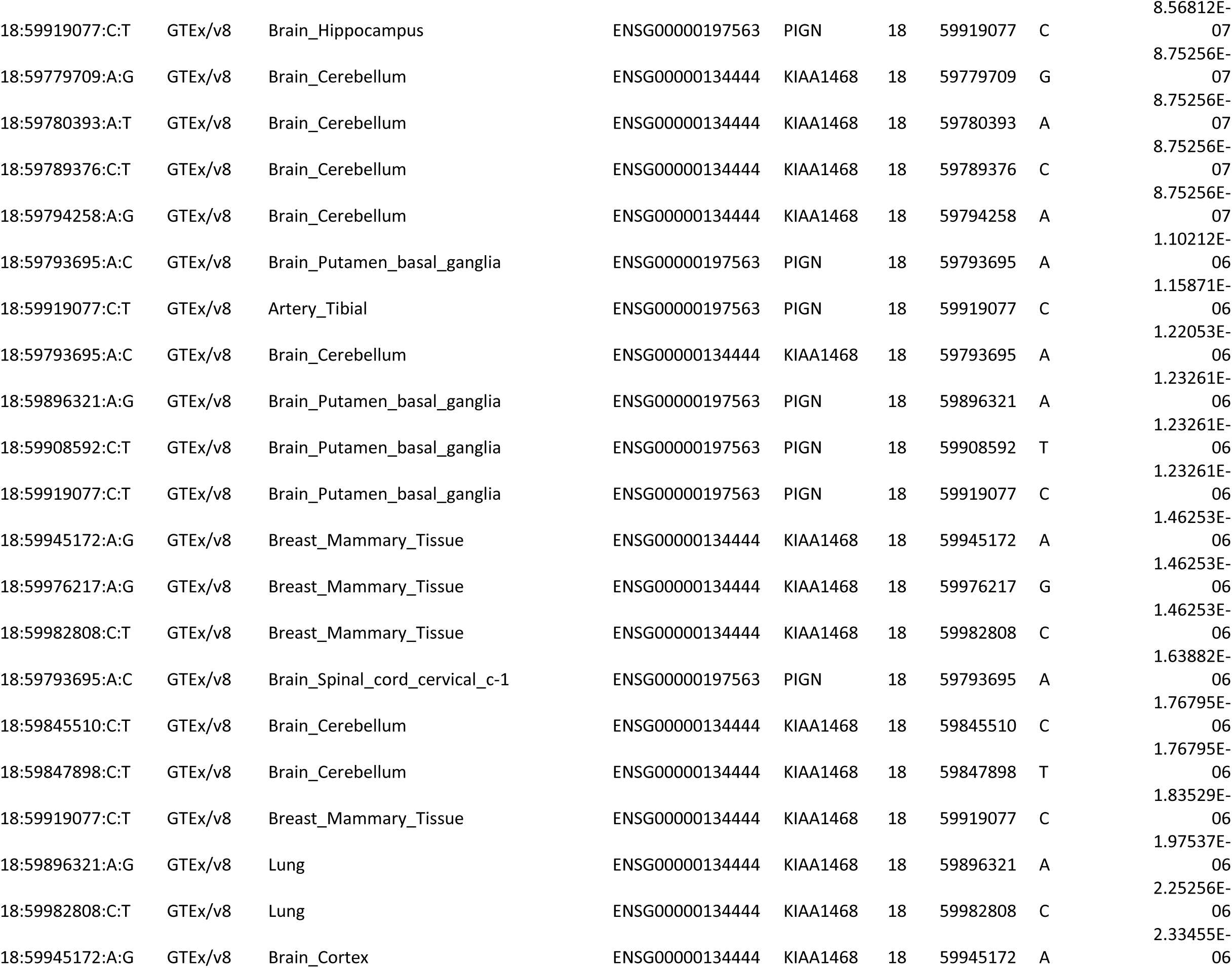
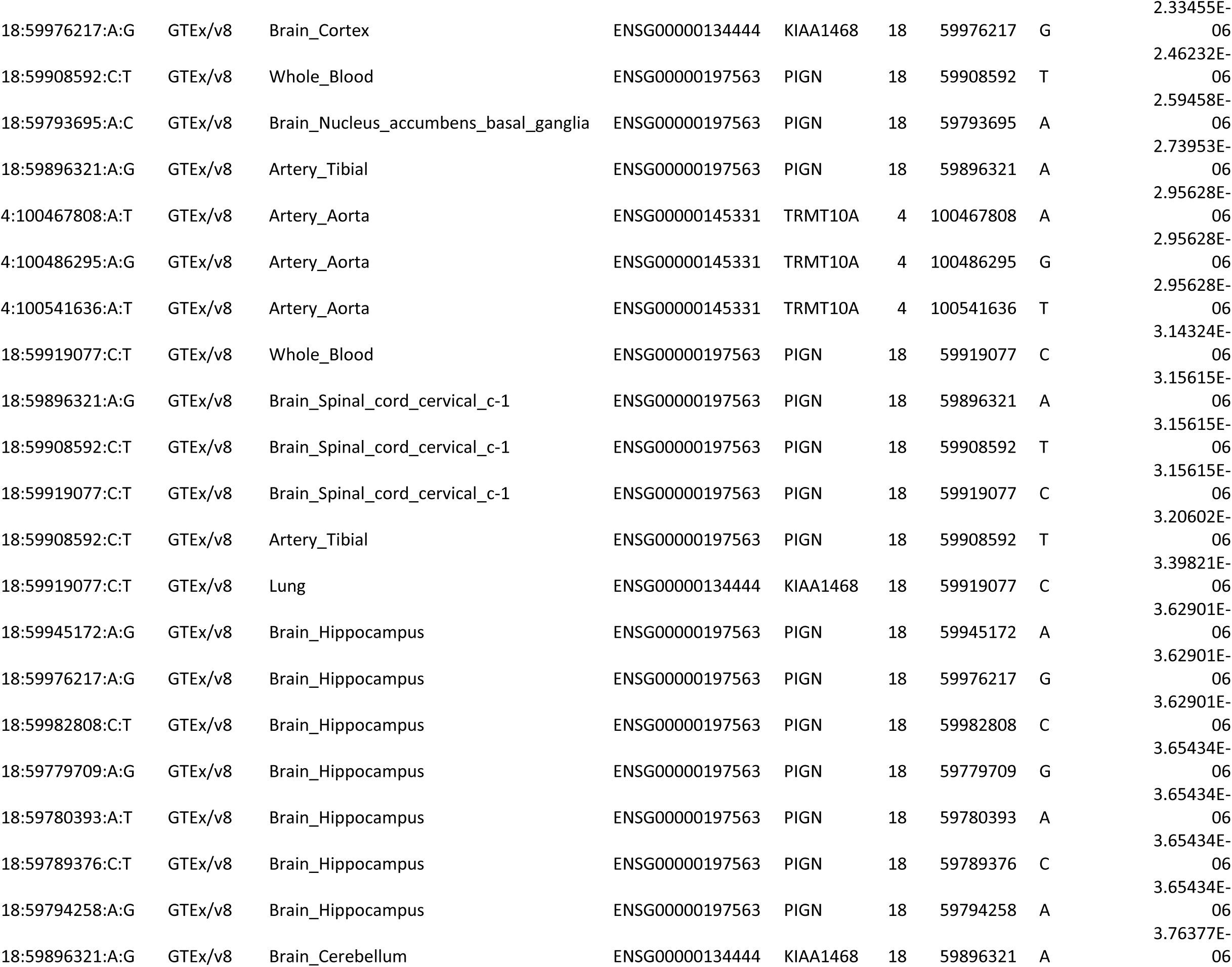
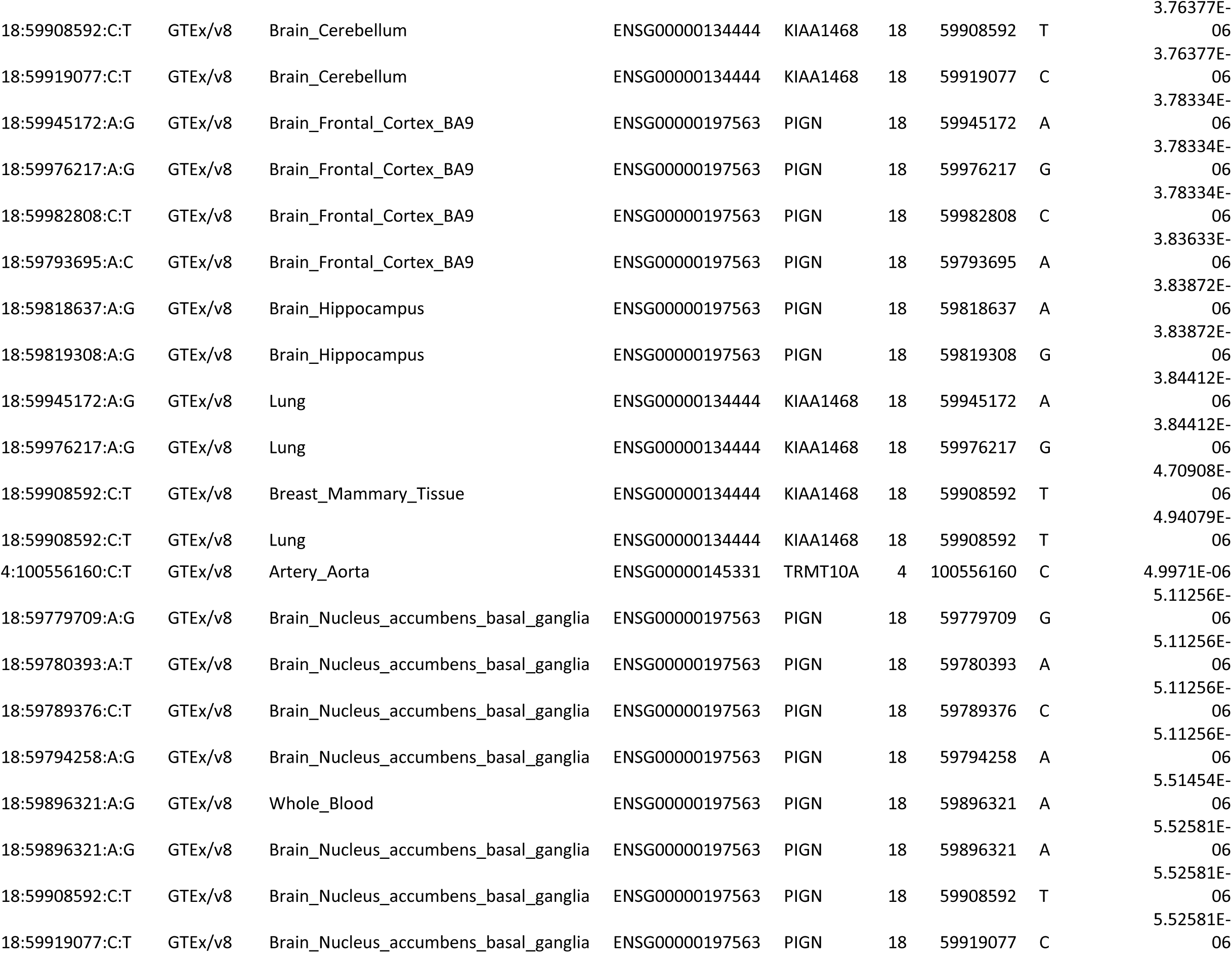
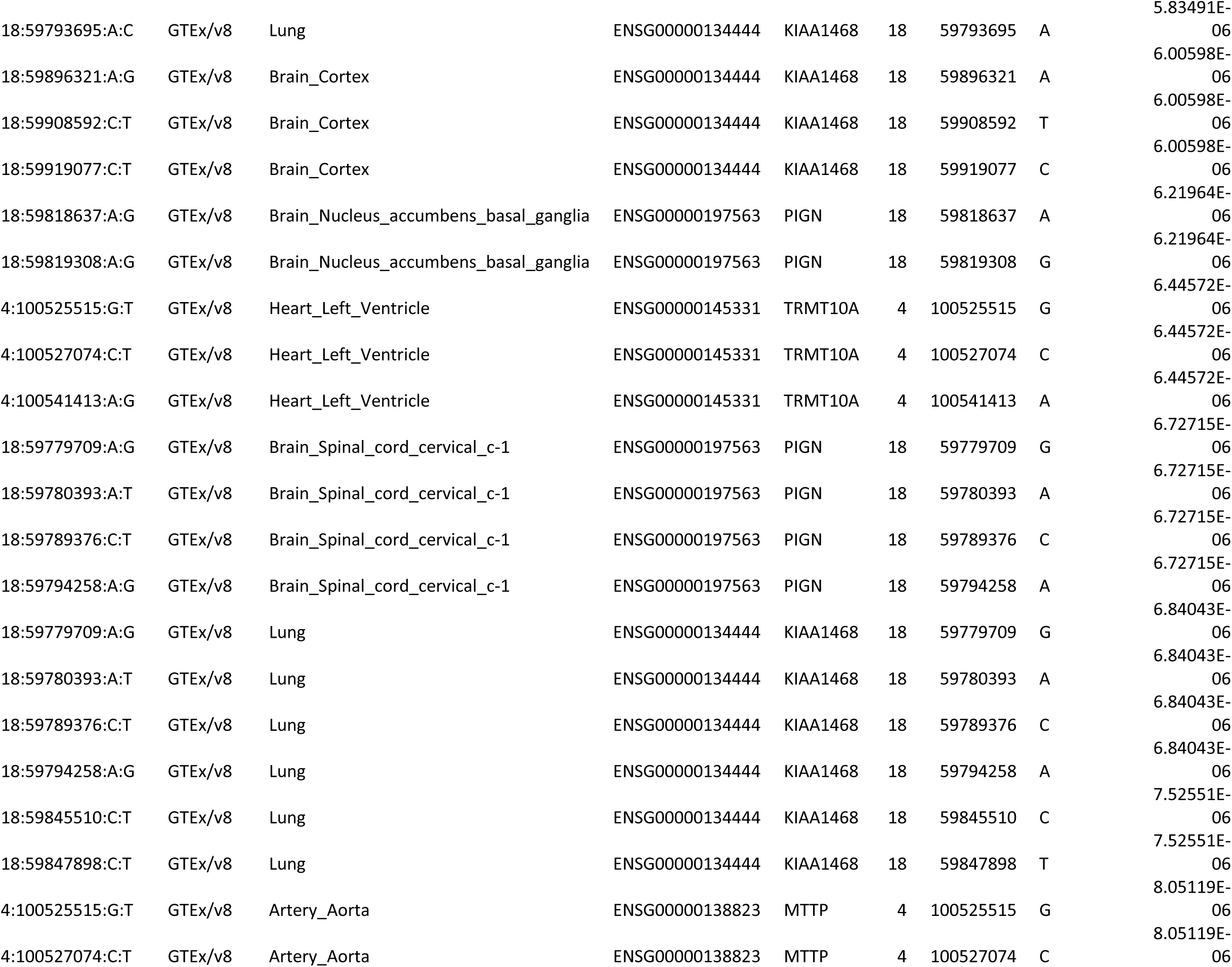
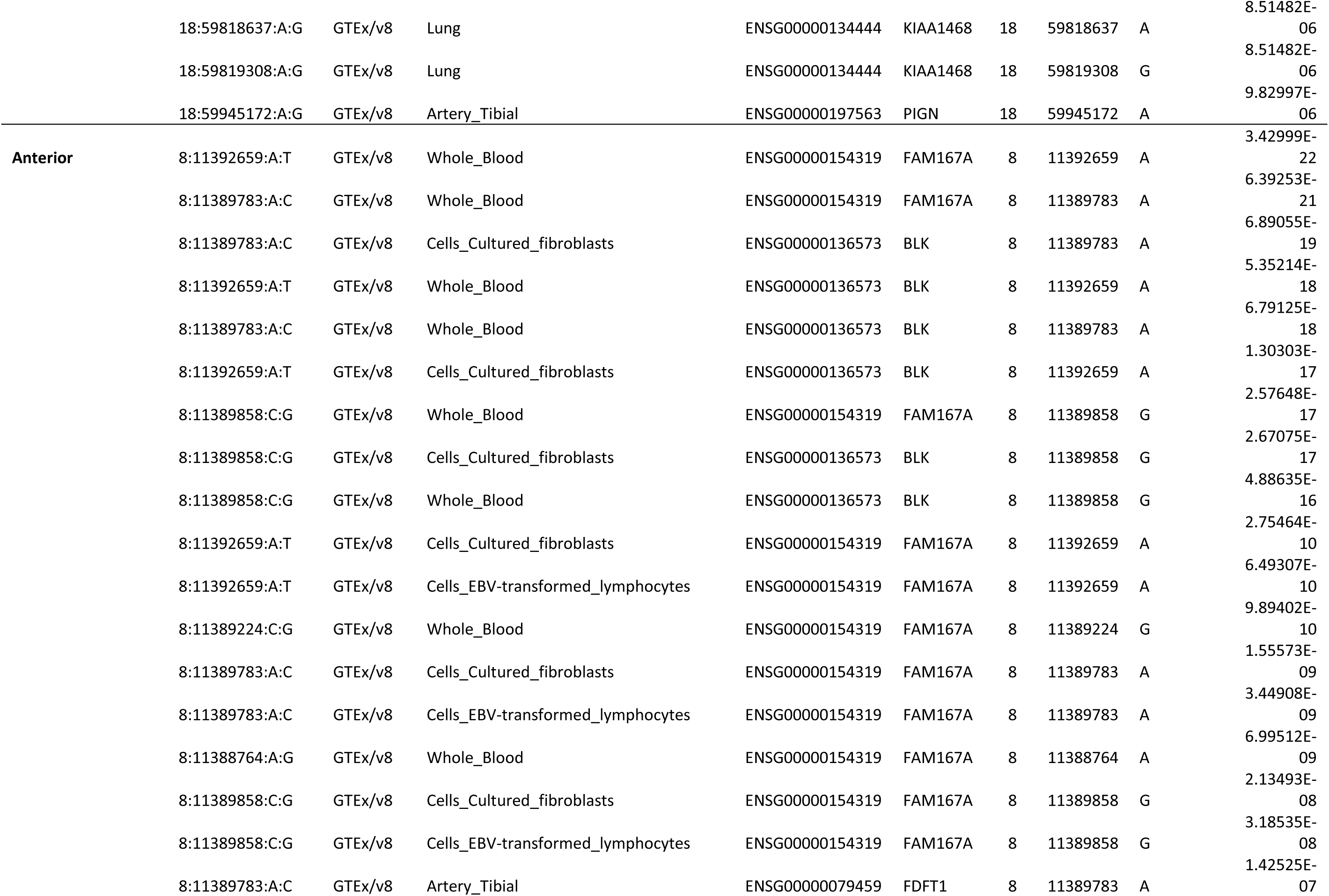
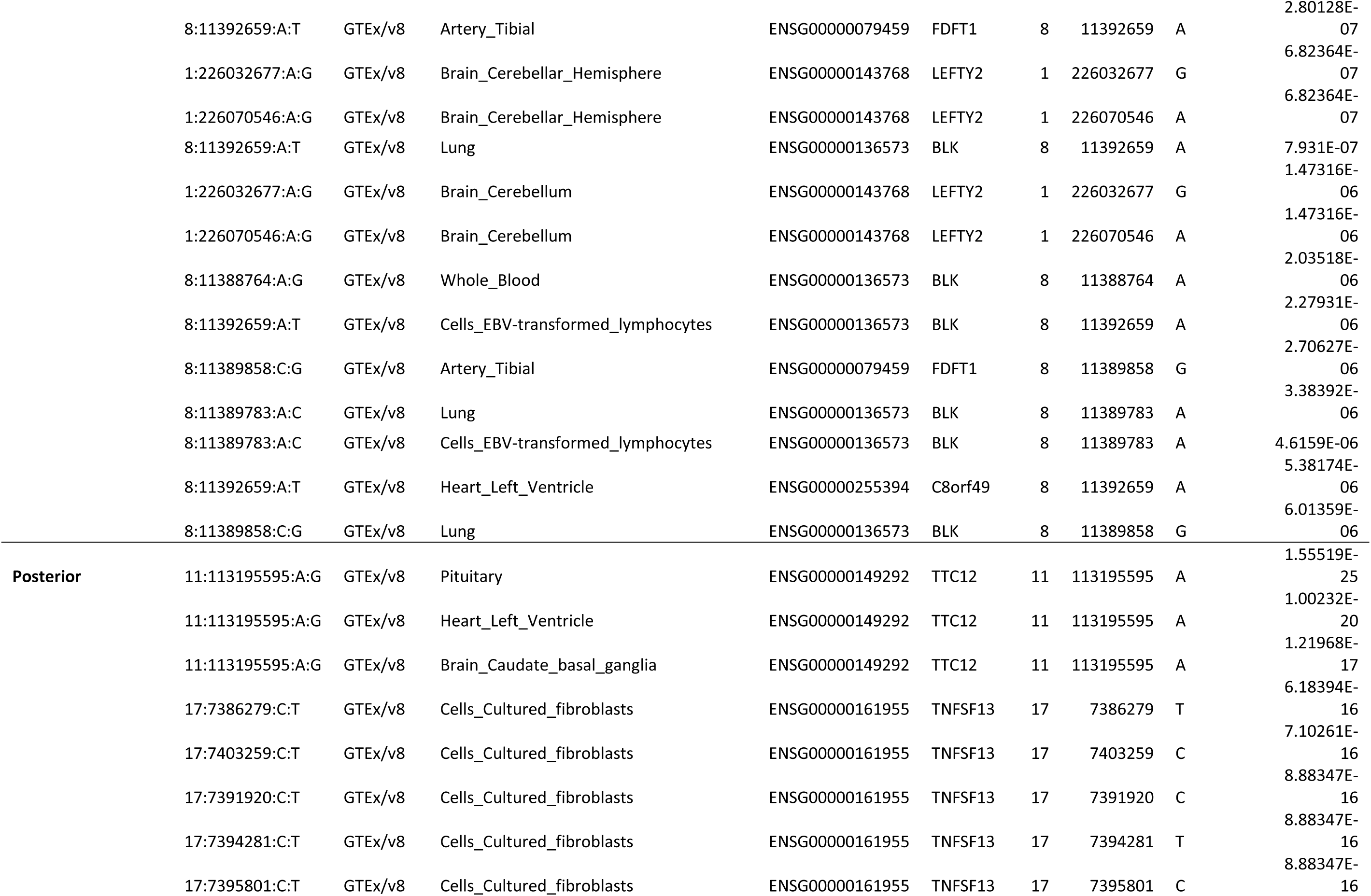
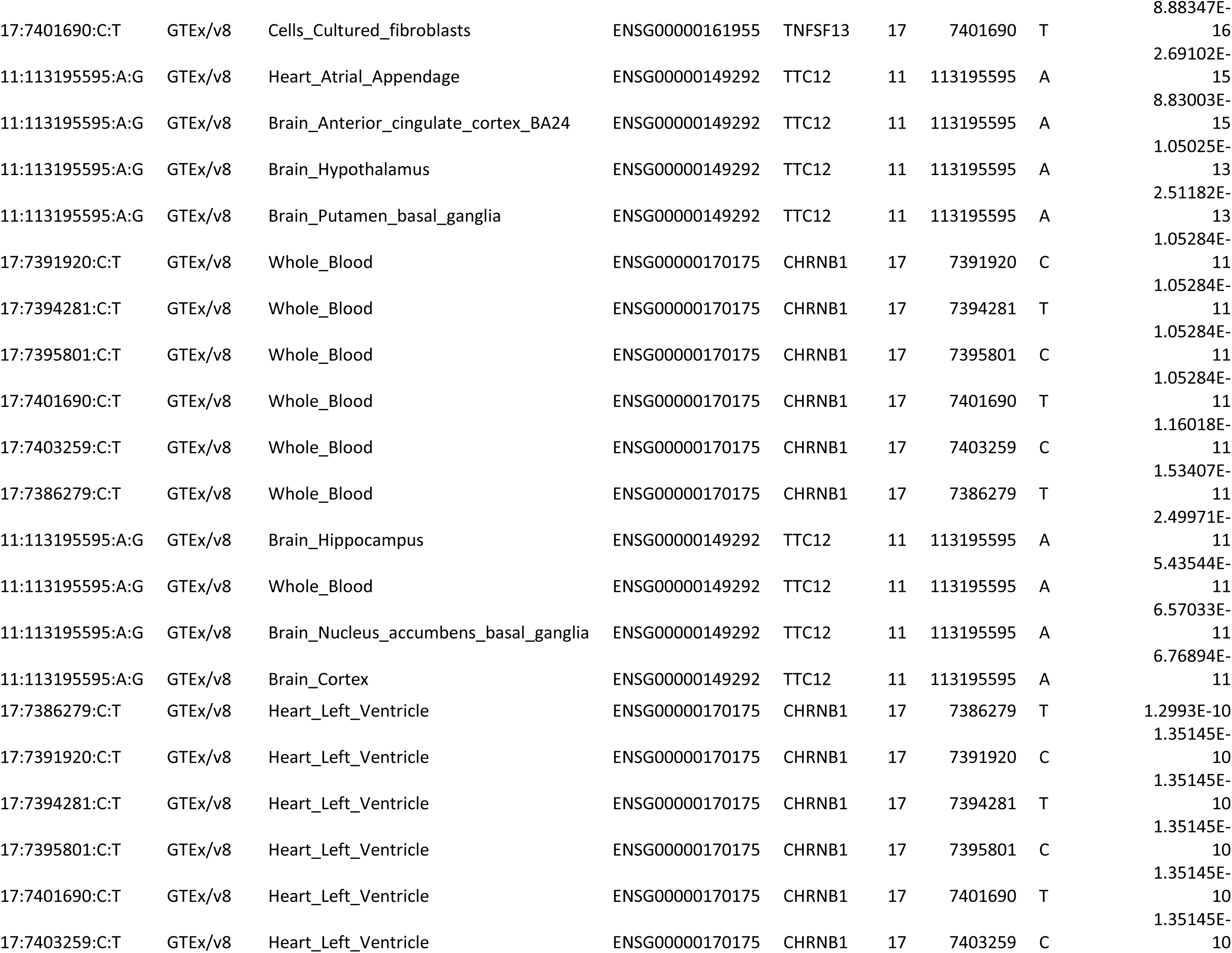
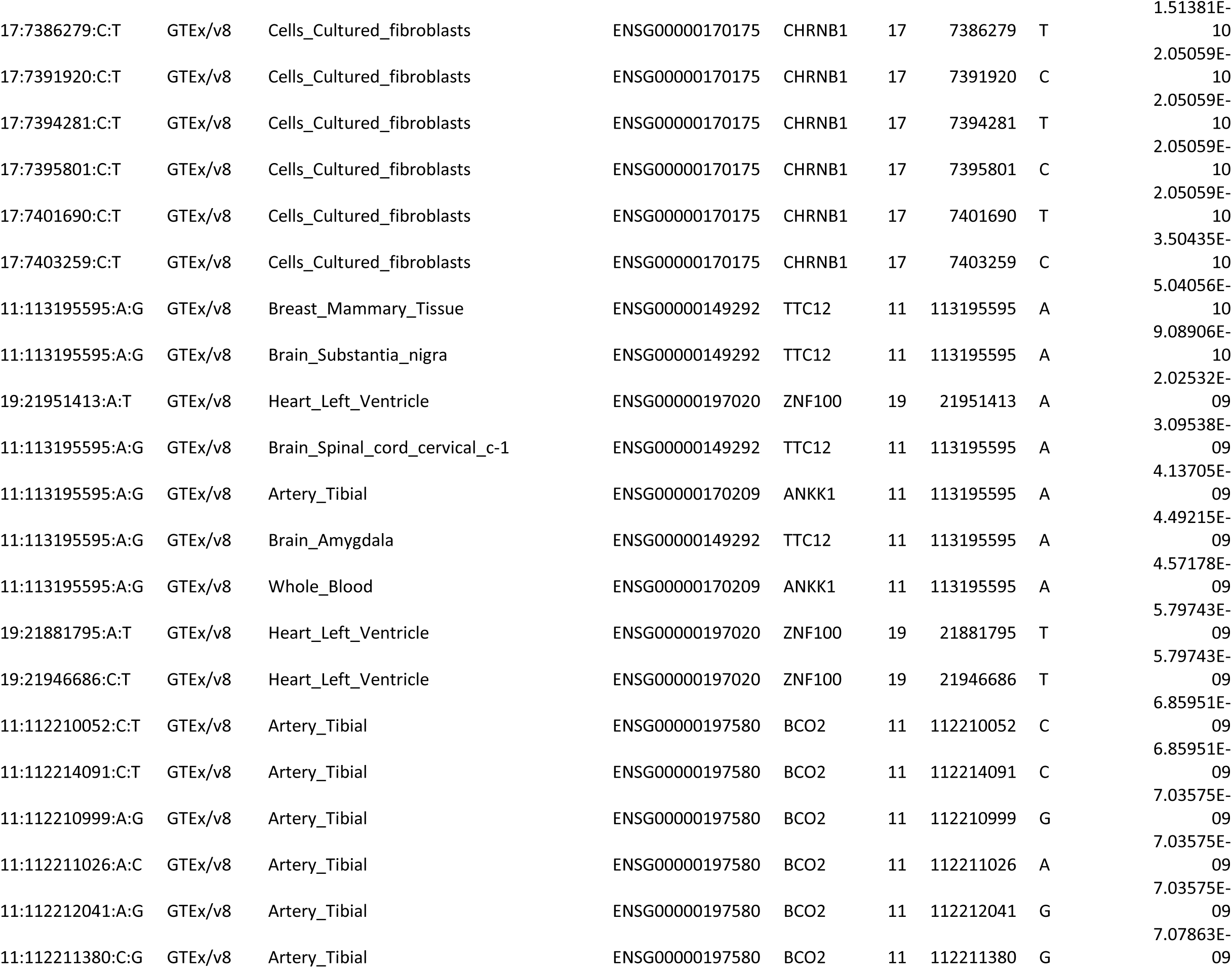
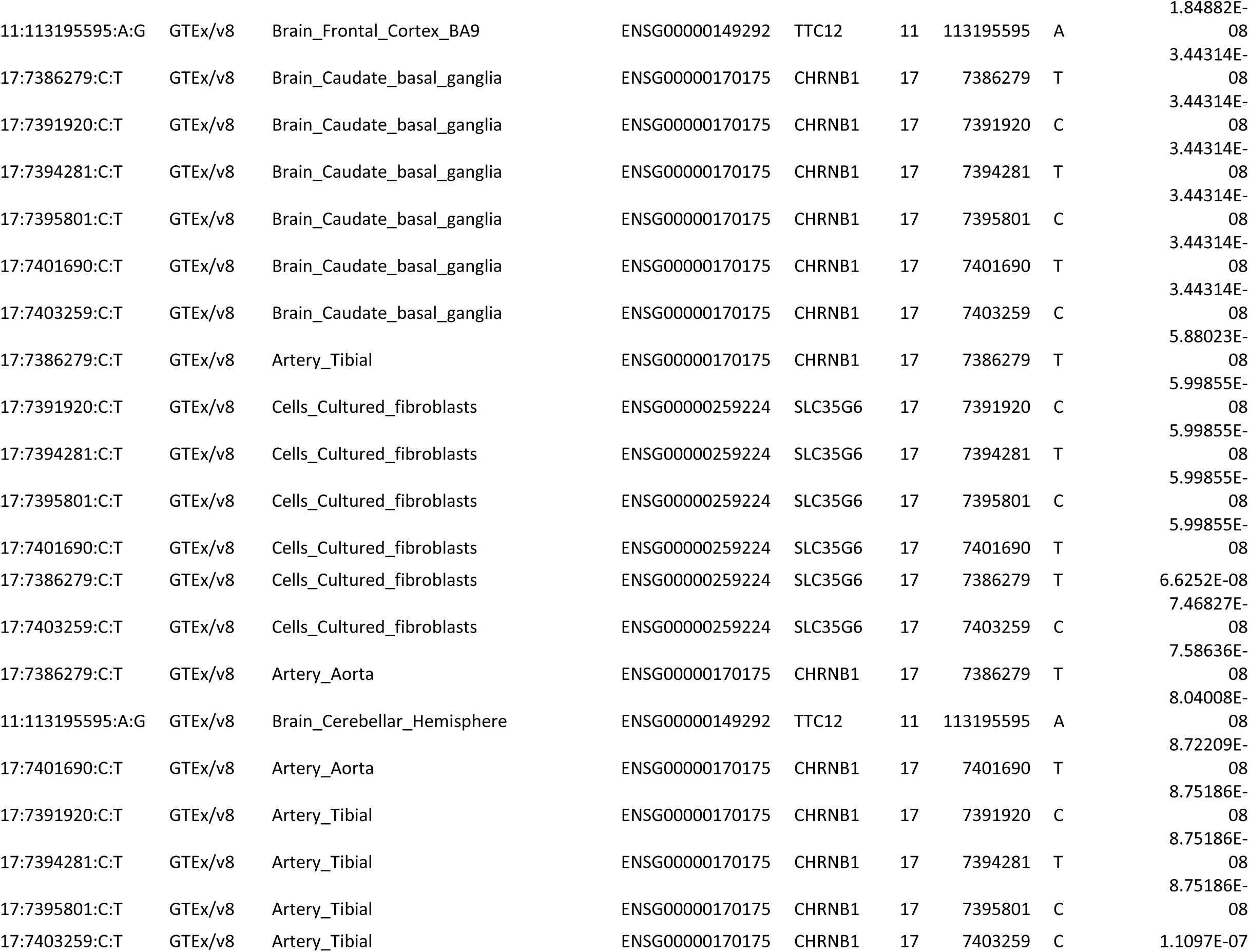
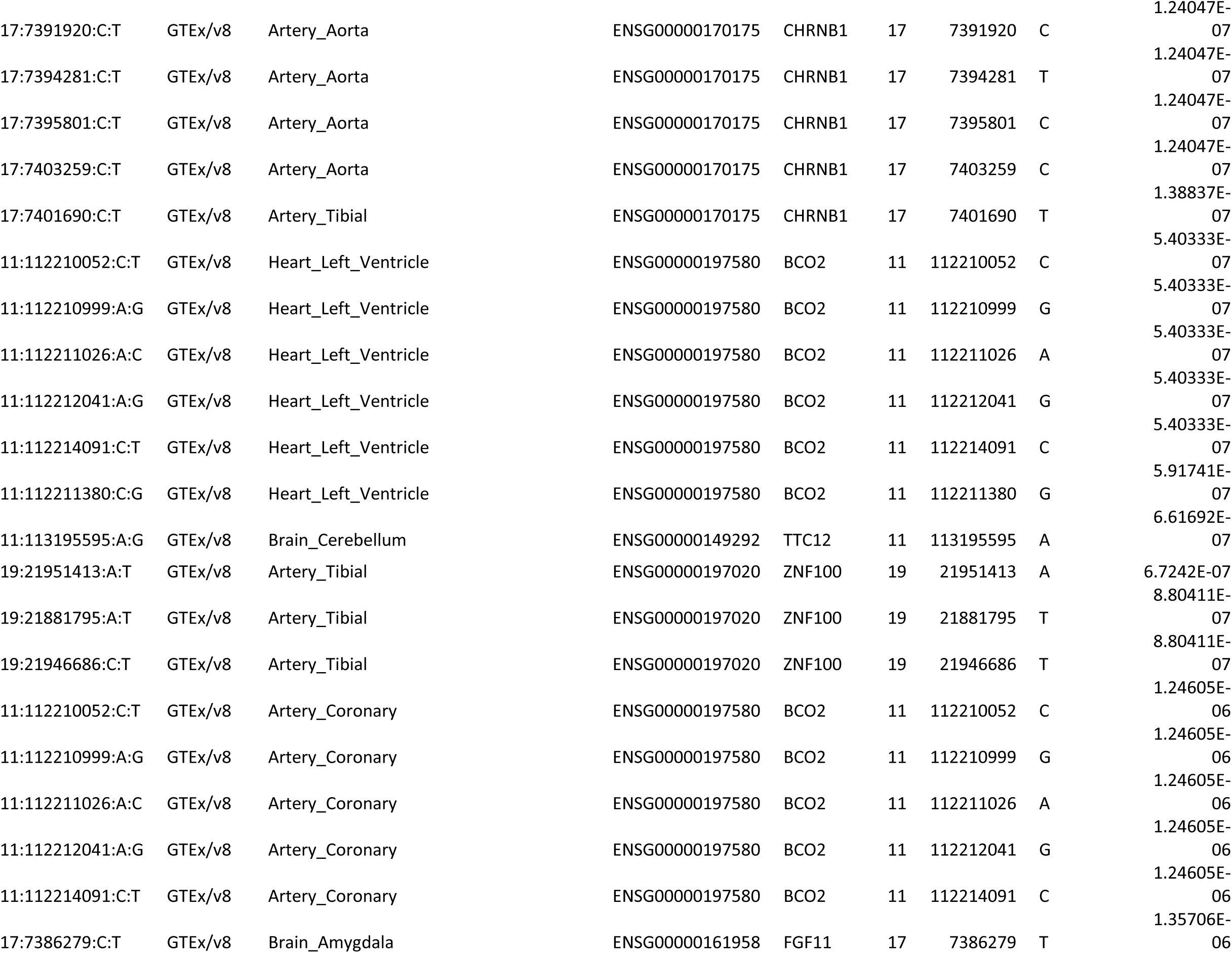
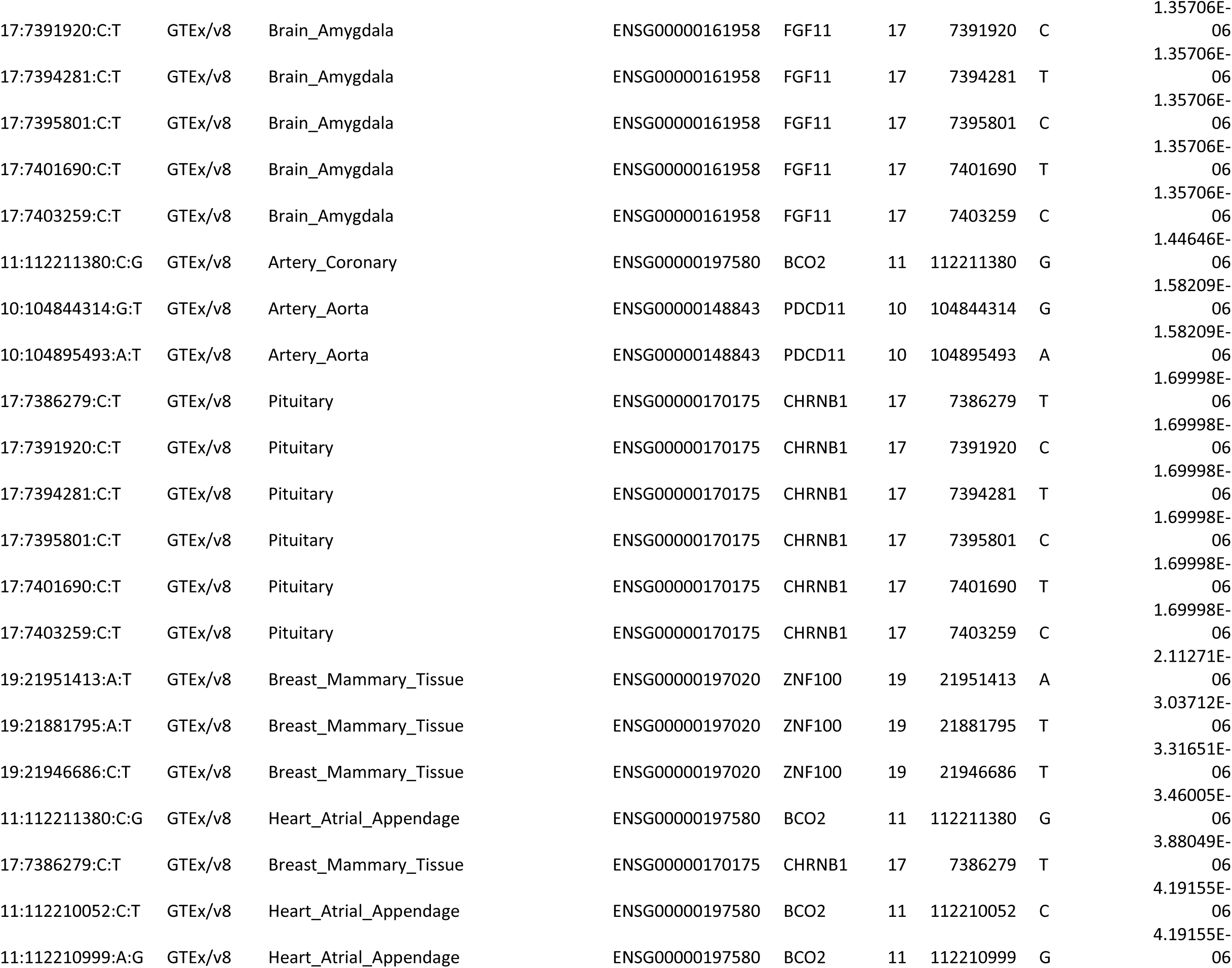
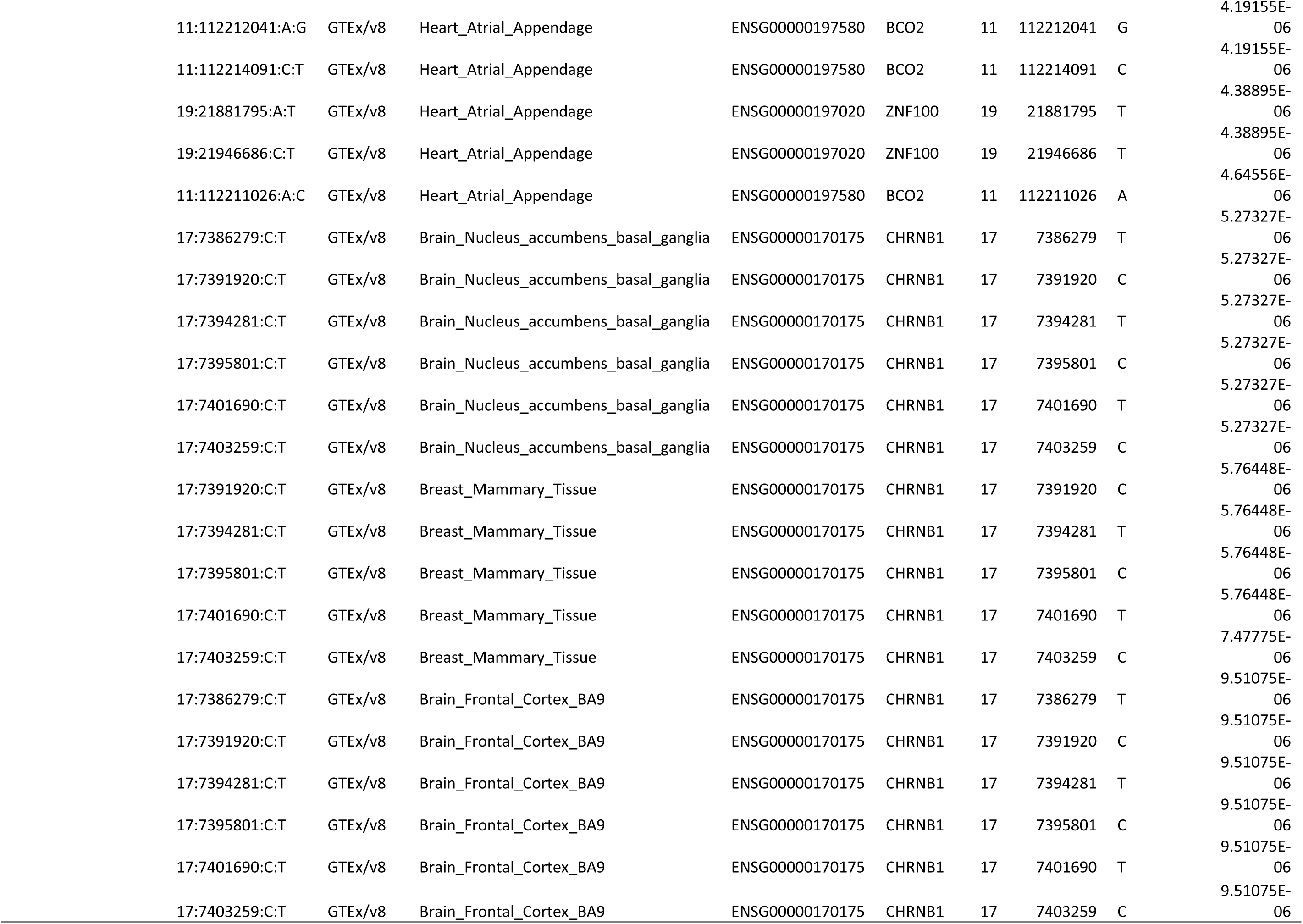
Co-localization of ILAS with Tissue-Specific Gene Expression.

The circos plot showed multiple chromatin interactions between the genomic risk locus on chromosome 1 and genes *TMEM206*, *NEK2*, and *LPGAT1* for global ILAS (Figure 9A); chromosome 1 and genes *SDE2*, *LEGTY2*, *SRP9*, *ENAH*, *LBR*, *TMEM63A*, *TMEM206*, *NEK2*, and *LPGAT1* for anterior ILAS (Figure 9B). For the posterior ILAS, the loci associated with lead SNPs had chromatin interaction with multiple genes (Figure 9C).

### Mendelian Randomization

To establish a causal pathway from ILAS to stroke, coronary artery disease, and atrial fibrillation, we performed a Mendelian Randomization (MR) analysis. We did not observe any association of ILAS with stroke, cardiovascular disease, or atrial fibrillation (Table 8, Figure 10).

**Table 8.** Mendelian Randomization Analysis.

| Outcomes | Method | Beta | se | P |
| --- | --- | --- | --- | --- |
| Ischemic stroke | Simple median | 0.025 | 0.021 | 0.237 |
|  | Weighted median | 0.025 | 0.021 | 0.23 |
|  | Inverse variance weighted (IVW) | 0.017 | 0.016 | 0.292 |
|  | MR Egger | 0.044 | 0.041 | 0.288 |
|  | MR Egger intercept | -0.021 | 0.031 | 0.484 |
| Small vessel stroke | Simple median | 0.001 | 0.028 | 0.983 |
|  | Weighted median | 0.001 | 0.028 | 0.988 |
|  | Inverse variance weighted (IVW) | 0.002 | 0.022 | 0.918 |
|  | MR Egger | 0.012 | 0.078 | 0.882 |
|  | MR Egger intercept | -0.006 | 0.048 | 0.901 |
| Coronary Artery Disease | Simple median | 0.01 | 0.042 | 0.811 |
|  | Weighted median | 0.01 | 0.042 | 0.811 |
|  | Inverse variance weighted (IVW) | 0.017 | 0.034 | 0.621 |
|  | MR Egger | -0.042 | 0.115 | 0.717 |
|  | MR Egger intercept | 0.041 | 0.078 | 0.594 |
| Atrial Fibrillation | Simple median | 0.008 | 0.03 | 0.8 |
|  | Weighted median | 0.007 | 0.03 | 0.815 |
|  | Inverse variance weighted (IVW) | 0.006 | 0.024 | 0.816 |
|  | MR Egger | -0.008 | 0.098 | 0.938 |
|  | MR Egger intercept | 0.008 | 0.058 | 0.89 |

**Figure 10.**
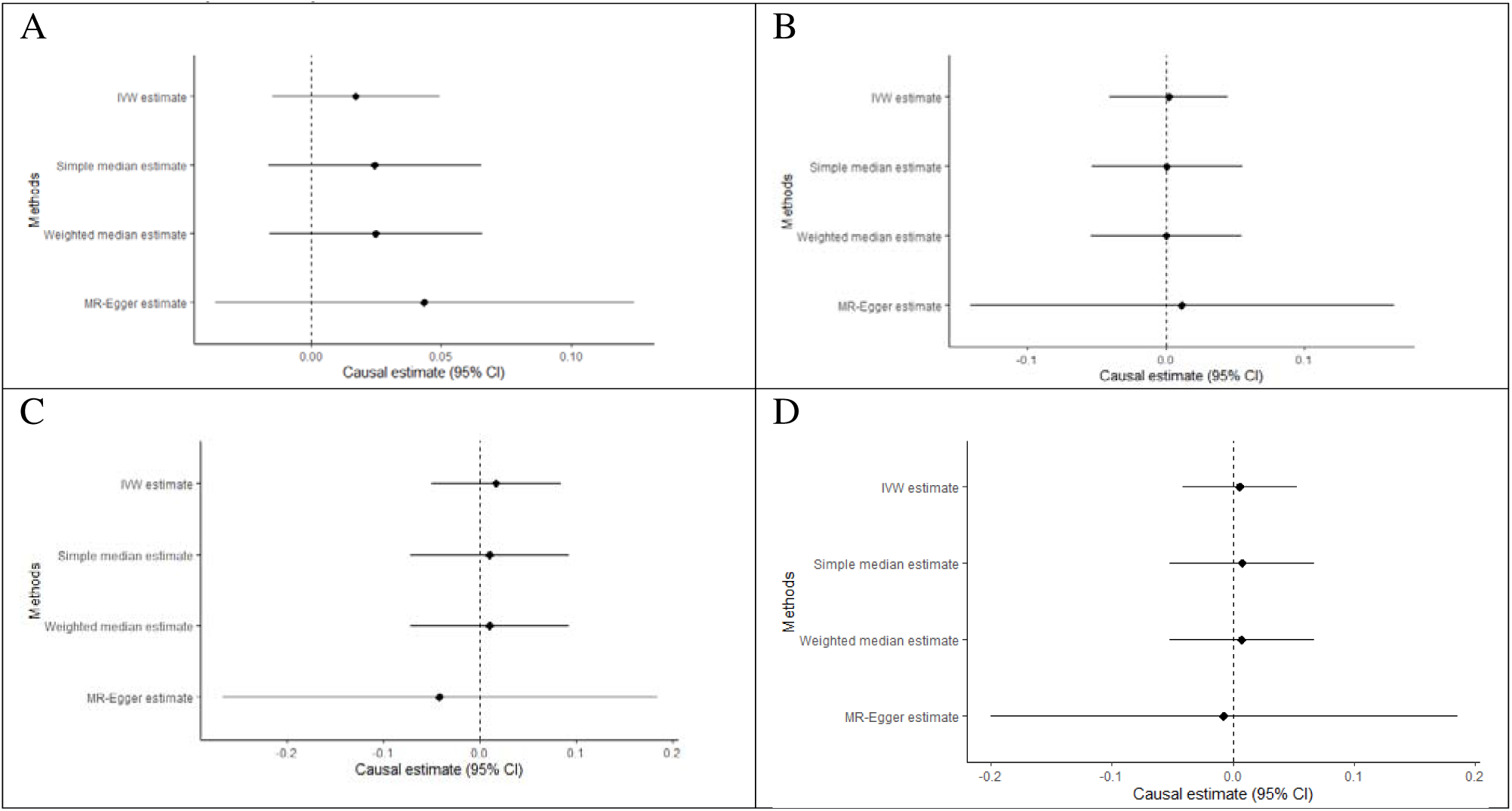
Forest plots of Two-sample MR analysis. (A) Ischemic stroke. (B) Small vessel stroke. (C) Coronary Artery Disease. (D) Atrial Fibrillation.

**Figure 11.**
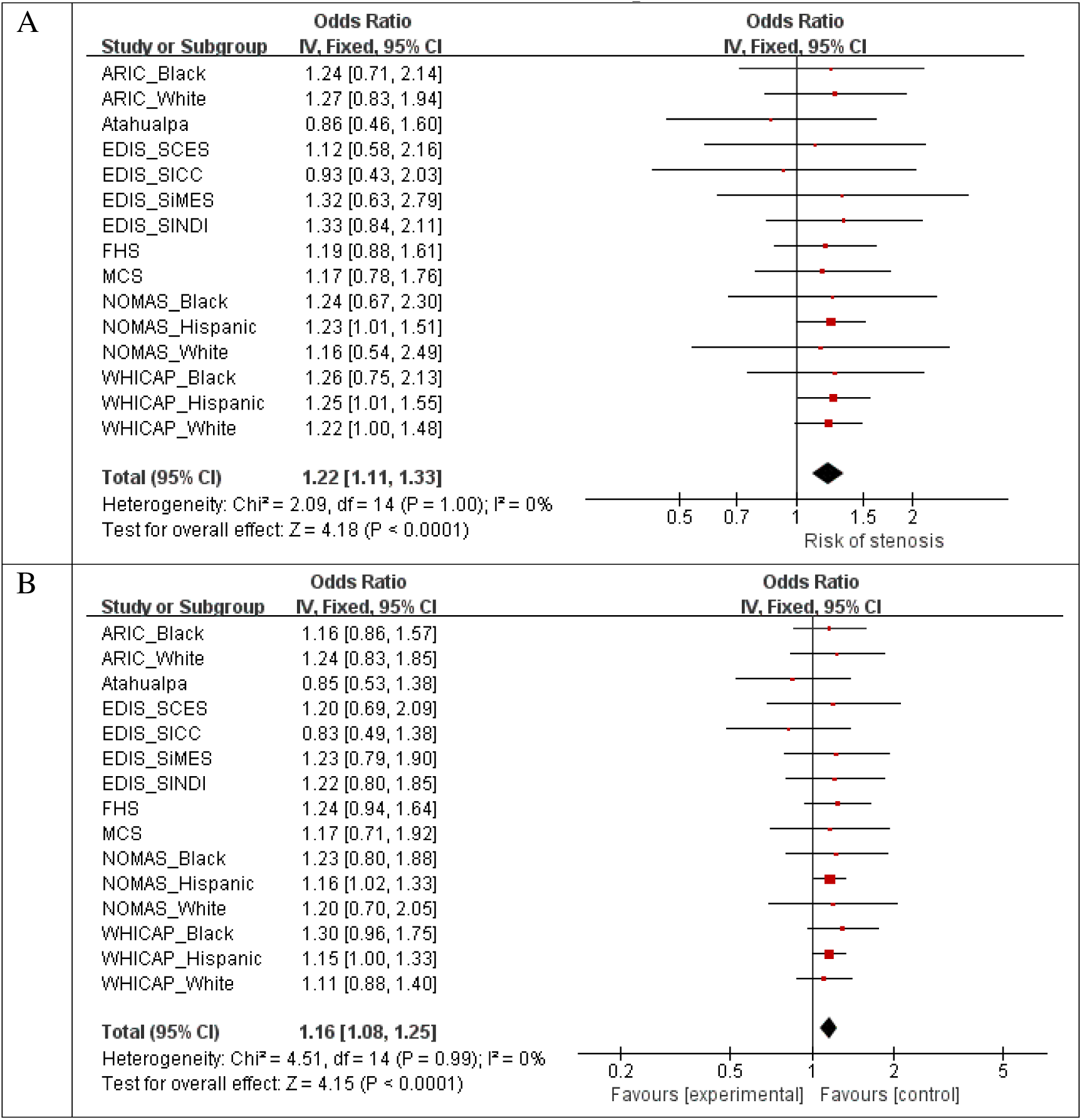
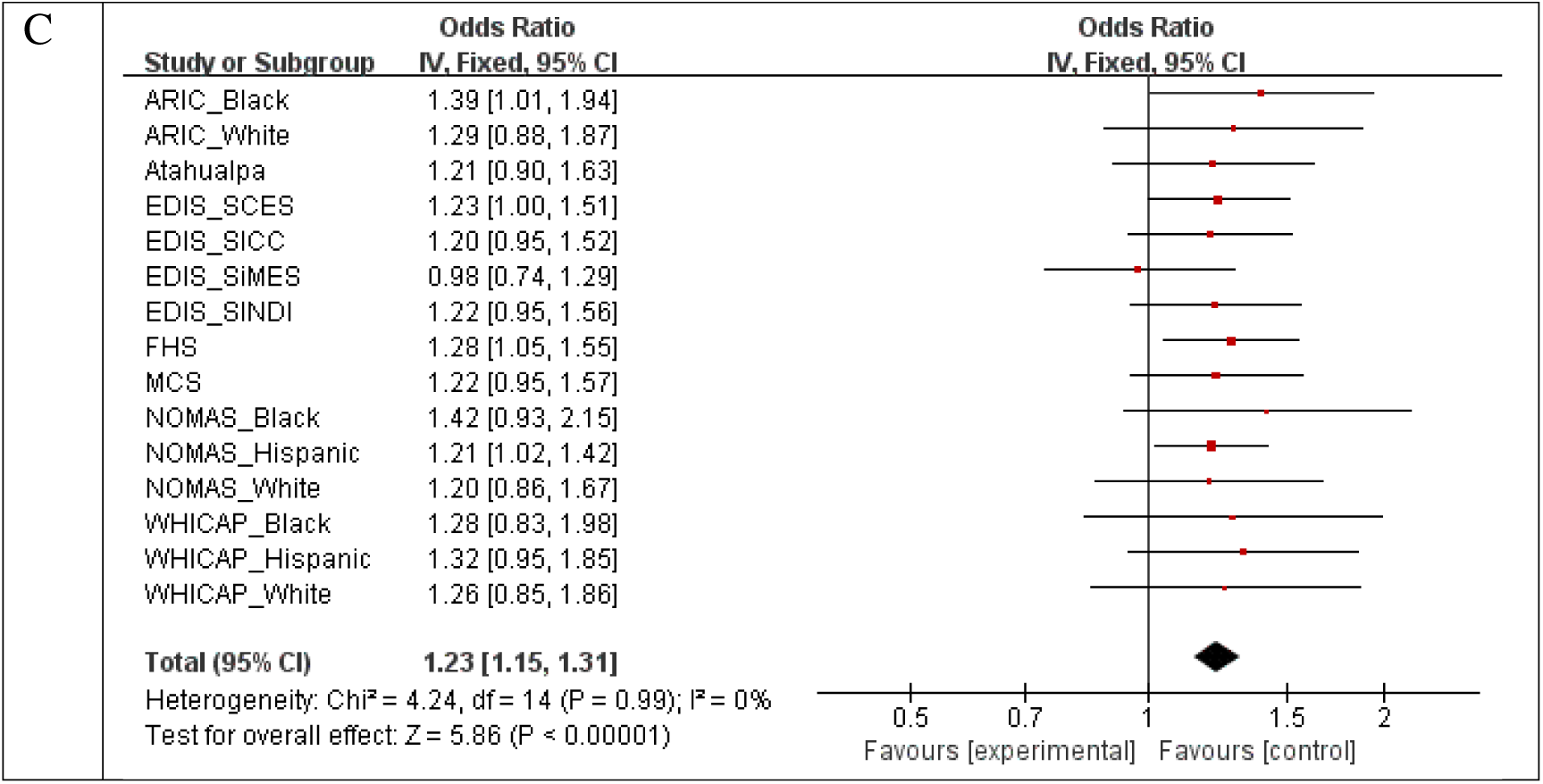
Forest plots for the meta-analysis of top variants. (A) rs75615271 in global ILAS. (B) rs75615271 in anterior ILAS. (C) rs62238282 in posterior ILAS.

**Table 9.**
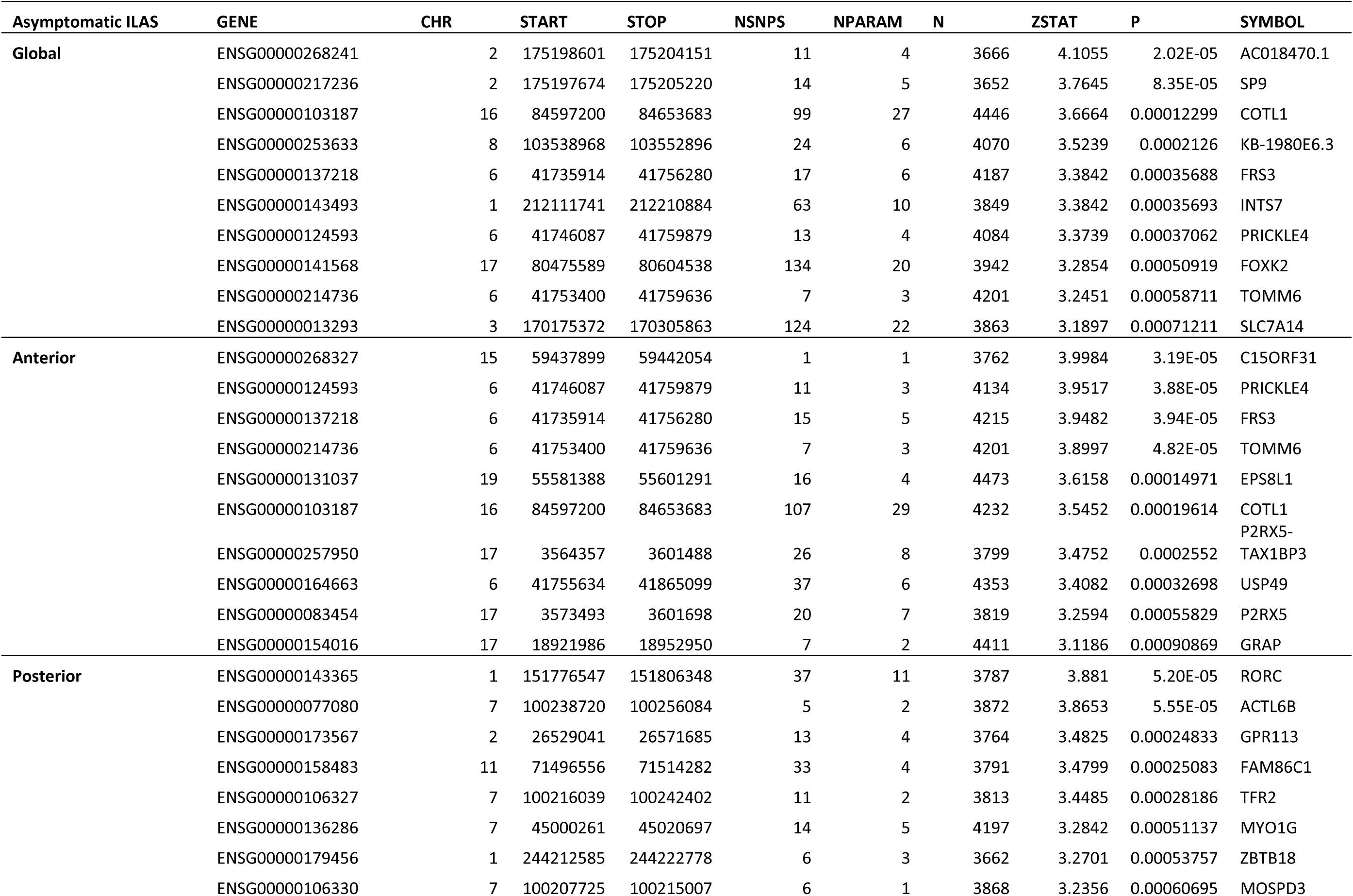
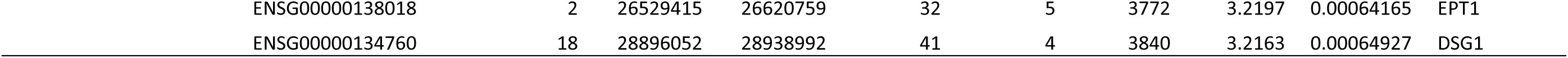
MAGMA gene-based association.

## Discussion

This is the first study to investigate the genetic determinants of asymptomatic ILAS in multi-populations. We identified associations of novel genetic loci with asymptomatic ILAS genetic architecture. Beyond mapping to the nearest genes, we also showed the biological impact of our findings using in silico functional analyses. Our results demonstrated that genetic loci are coupled with gene expression information, which imply biologically relevant pathways.

In this multi-population analysis, we observed a novel variant rs75615271 mapped to *RP11-552D8.1* that has a significant association with global ILAS. *RP11-552D8.1* is PGAT1 antisense RNA 1 (LPGAT1-AS1), located in the Chr1p31, which plays a crucial role in regulating gene expression at multiple levels. Lysophosphatidylglycerol acyltransferase 1 (*LPGAT1*) encodes protein that catalyzes the reacylation of lysophosphatidylglycerol into phosphatidylglycerol. It is a key precursor for the cardiolipin synthesis, which is involved in lipid biosynthesis ^41^. *LPGAT1* has been reported to regulate the biosynthesis of triacylglycerol, which is important for maintaining phospholipid homeostasis and modulating the structural integrity of mitochondrial membranes^42^. Additionally, *LPGAT1* plays a role in lipid metabolism, and its impact on body mass index and body fat have been confirmed ^43^. These effects of *LPGAT1* on the organism occur when it is in its regular expression profile, but elevated expression levels may contribute to the development of certain diseases. Previous studies showed that *LPGAT1* gene expression is upregulated in tumor tissue compared to normal tissue ^44–46^. In our study, the variant *RP11-552D8.1* rs75615271 was associated with presence of asymptomatic ILAS.

A gene set associated with asymptomatic ILAS was identified, including *LPGAT1*, never in mitosis related kinase 2 (*NEK2*), integrator complex subunit 7 (*INTS7*), denticleless E3 ubiquitin protein ligase homolog (*DTL*), proton activated chloride channel 1 (*PACC1*, also known as *TMEM206*). *NEK2* encodes a serine/threonine-protein kinase that is essential for mitotic regulation. This protein localizes to the centrosome, and undetectable during G1 phase, but accumulates progressively throughout the S phase, reaching peak levels in late G2 phase^47^. *NEK2* is associated with poor prognosis of clear cell renal cell carcinoma and promotes tumor cell growth and metastasis ^48^. High expression of *NEK2* was associated with vascular invasion and tumor grade in multiple patient cohorts of pancreatic cancer^49^. *NEK2* is abnormally overexpressed in a wide range of human cancers and is implicated in various aspects of malignant transformation, including tumorigenesis, drug resistance and tumor progression^50^. *INTS7* encodes a subunit of the integrator complex that is associated with the C-terminal domain of RNA polymerase II and mediates 3’-end processing of the small nuclear RNAs U1 and U2. The expression level of *INTS7* may correlate with tumor microenvireoment, immunotherapy responsiveness^51^. Several studies showed that *INTS7* is upregulated in several solid tumors, such as cholangiocarcinoma, hepatocarcinoma, cervical squamous cell carcinoma, endocervical adenocarcinoma, and breast cancer ^52^. In addition, *INTS7* has been linked to bipolar disorder ^53^. *DTL*, also known as *CDT2* gene, contributes to ubiquitin-protein transferase activity that is involved in several key processes, including protein ubiquitination, regulation of G2/M transition of mitotic cell cycle, and translesion synthesis^54^. *CDT2* contains multiple WD40-repeat domains that play an essential role in regulating the *CDT1* degradation after DNA damage ^55^. Previous studies have shown that the CRL4-CDT2 complex, together with Rad6/18, monoubiquitinated PCNA promote the translation DNA synthesis in undamaged cells ^56^. In addition, the CRL4-CDT2 complex can degrade DNA replication-related proteins in a proteasome-dependent manner during DNA replication, implying a crucial role of *CDT2* in the regulation of DNA replication ^57^. Further research revealed that *CDT2* is augmented in head and neck squamous cell carcinoma (HNSCC) and is necessary for those tumor cells to proliferate. Its main role is to inhibit abnormal DNA replication. Inactivation of CRL4-CDT2 increases the radiosensitivity of HNSCC cells^58^. Moreover, USP46 protein could mediate the stability of *CDT2* and promote the growth of HPV-positive tumors, suggesting the potential role of *DTL* in tumor progression ^59^. *PACC1*, also known as *TMEM206*, is an integral component of plasma membrane, which is involved in pH-gated chloride channel activity and chloride transport^60^. Previous studies revealed that *TMEM206* is linked to cell volume changes under acidic pH and had the functions of the proton-activated Cl^−^ channel^61–64^. A recent study indicated the key role of *TMEM206* in macropinosome resolution ^65^. Macropinocytosis is of central importance for cancer cells, as they employ it to take up nutrients and proliferate in hypoxic, acidic and nutrient poor environments^66^.

Purinergic receptor P2X 5 (*P2RX5*) was identified as a Hispanic-specific gene related to anterior ILAS. P2X5 is a member of the P2X family of ATP-gated nonselective cation channels, which exist as trimeric assemblies^67^. *P2RX5* encodes the P2X5 purinergic receptor, a ligand-gated ion channel activated by ATP, and plays a role in endothelial cell differentiation and autocrine regulation ^68, 69^, as well as having functional roles in adult mouse astrocytes ^70^. A study showed surface and intracellular *P2RX5* expression was upregulated in activated antigen-specific CD4+ T cell clones, which indicated a functional role of the human *P2RX5* splice variant in T cell activation and immunoregulation ^71^. In humans, *P2RX5* exists as a natural deletion mutant lacking amino acids 328–349 of exon 10, meaning that only a proportion of the human population express fully functional P2X5 receptors, and amino acid substitutions within this gene can markedly impact the receptor’s responsiveness to its ATP ligand ^72^. A study using DNA methylation and genetics indicated *P2RX5* have related roles in the cerebrovascular system ^73^ .

Transmembrane Serine Protease 7 (*TMPRSS7*) was identified in an Asian-specific gene related to posterior ILAS. *TMPRSS7* encodes the protein that belongs to the type II transmembrane serine protease family, the 17 human members. They play physiological and pathological roles in digestion, cardiac function, and blood pressure regulation ^74–76^. They have also been implicated in tumor growth, invasion and metastasis, and the genetic variant rs1844925 of *TMPRSS7* has been associated with the risk for and prognosis of breast cancer ^77^. Another study reported that rs147783135 of *TMPRSS7* was related to ischemic stroke, with the minor T allele being protective against this condition ^78^. Given the potential roles of *TMPRSS7* in tumor growth and blood pressure regulation, the association of this gene with ischemic stroke may reflect an effect on atherosclerosis or blood pressure^74–76^.

Our study has some limitations that need to be recognized. First, due to the relatively modest sample sizes of each population, the statistical power to detect population-specific associations or functional associations was limited. Consequently, imbalance of cases and controls across different datasets may limit the applicability of the study’s finding to population-specific groups. Lastly, different datasets were genotyped using different GWAS platforms. It is not clear how this might have affected the imputation quality.

## Conclusions

In summary, we identified one significant variant associated with asymptomatic ILAS in a multi-population. Our study provides insights into a potential biological mechanism for the association between these loci and asymptomatic ILAS. Identifying genes associated with these loci and understanding their function may help us to elucidate the mechanism through which asymptomatic ILAS may influence cerebrovascular health.

## Declarations

## Data Availability

All data produced in the present study are available upon reasonable request to the authors.

