## Supplemental Tables and Figures for "Multi-population Genome-Wide Association Study Identifies Multiple Novel Loci associated with Asymptomatic Intracranial Large Artery Stenosis": Supp_Figures_04092025.docx

Figure 1 Manhattan Plot of GWAS summary statistics for Global ILAS. (A) Asian. (B) African American. (C) White. (D) Hispanic.

| A | 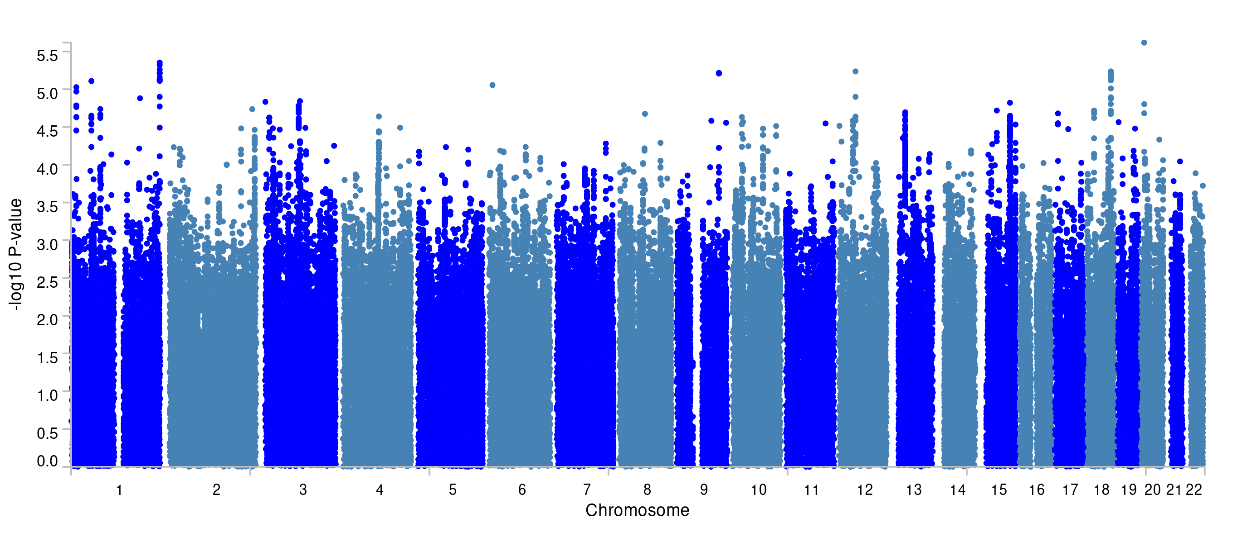 |
| --- | --- |
| B | 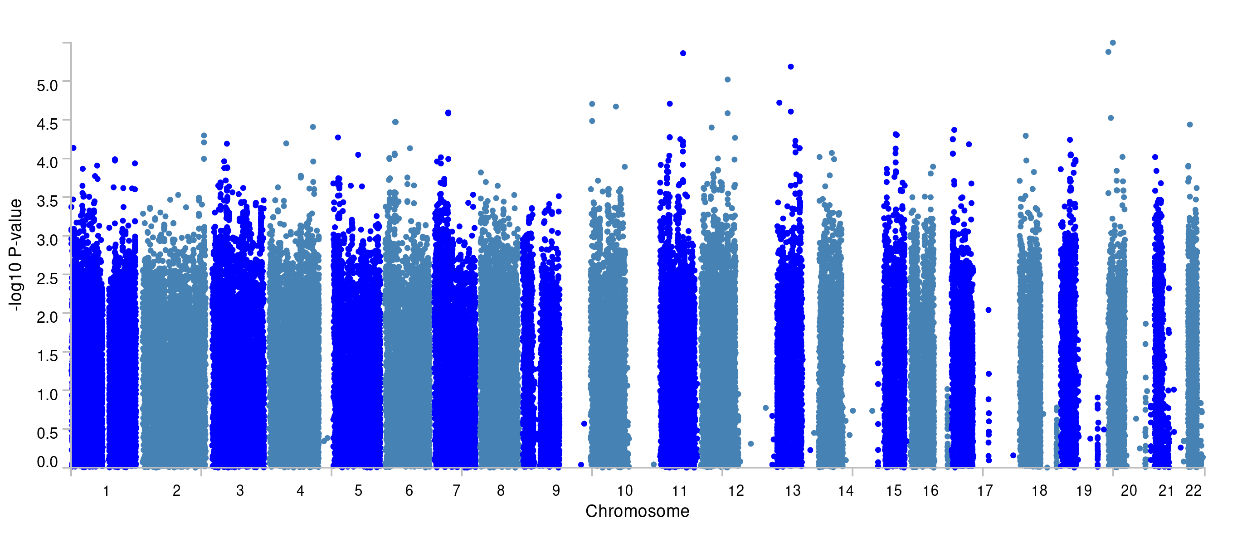 |
| C | 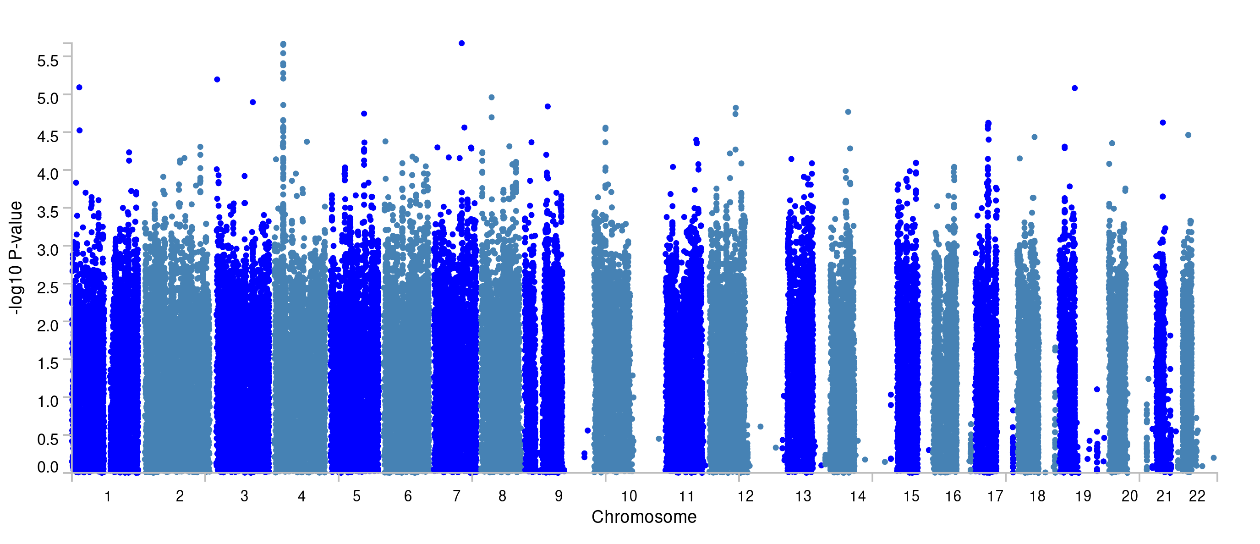 |
| D | 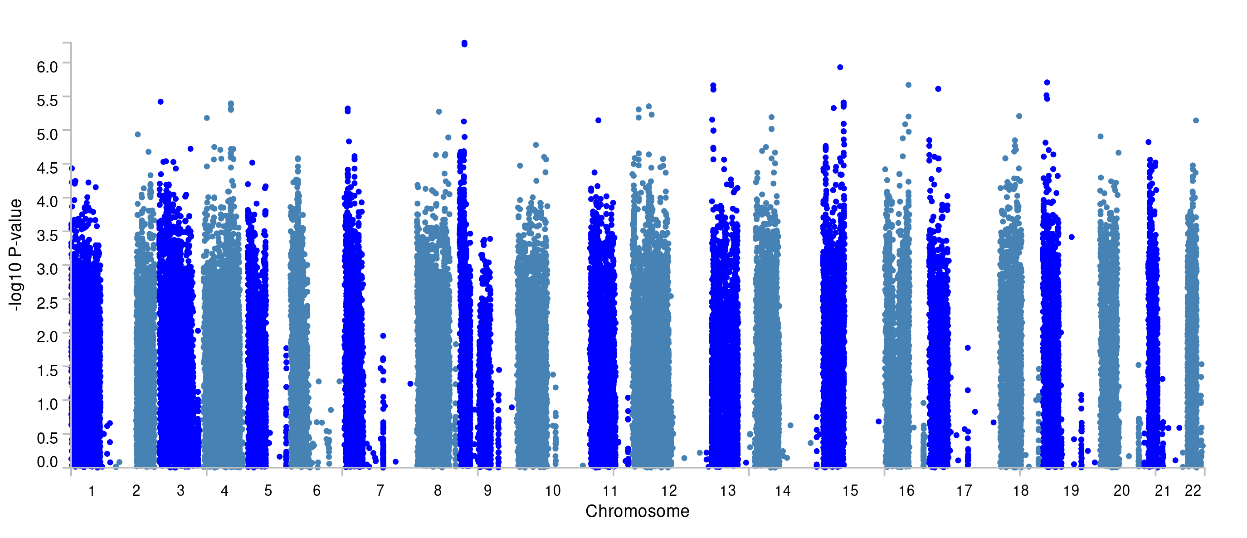 |

Figure 2 Manhattan Plot of GWAS summary statistics for Anterior ILAS. (A) Asian. (B) African American. (C) White. (D) Hispanic.

| A | 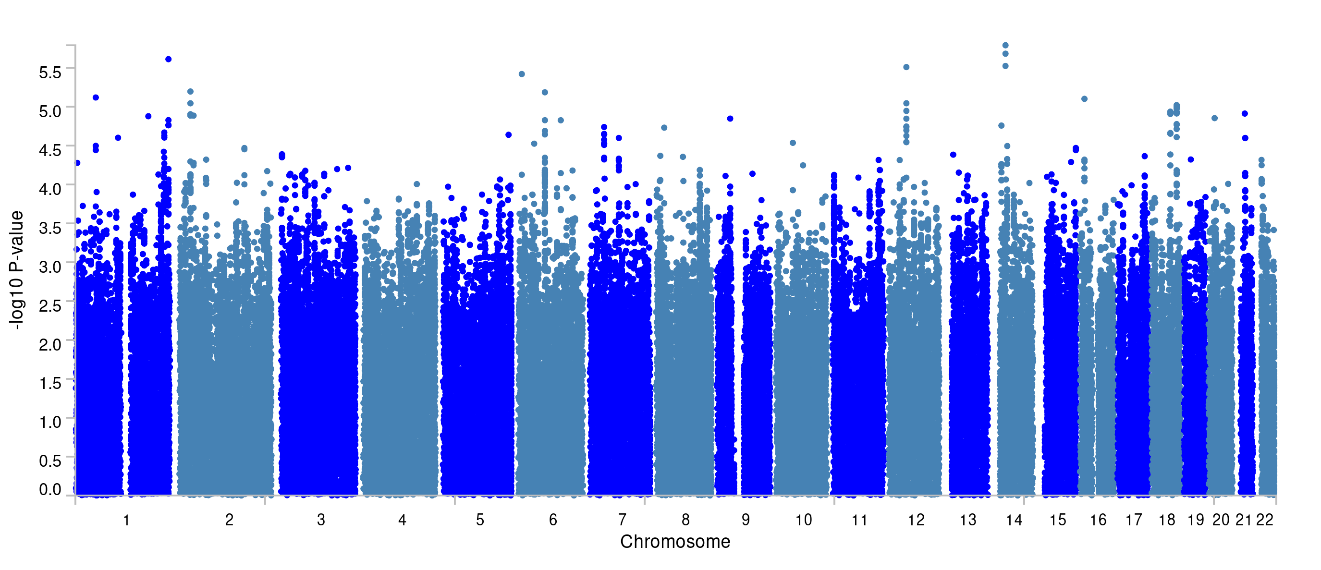 |
| --- | --- |
| B | 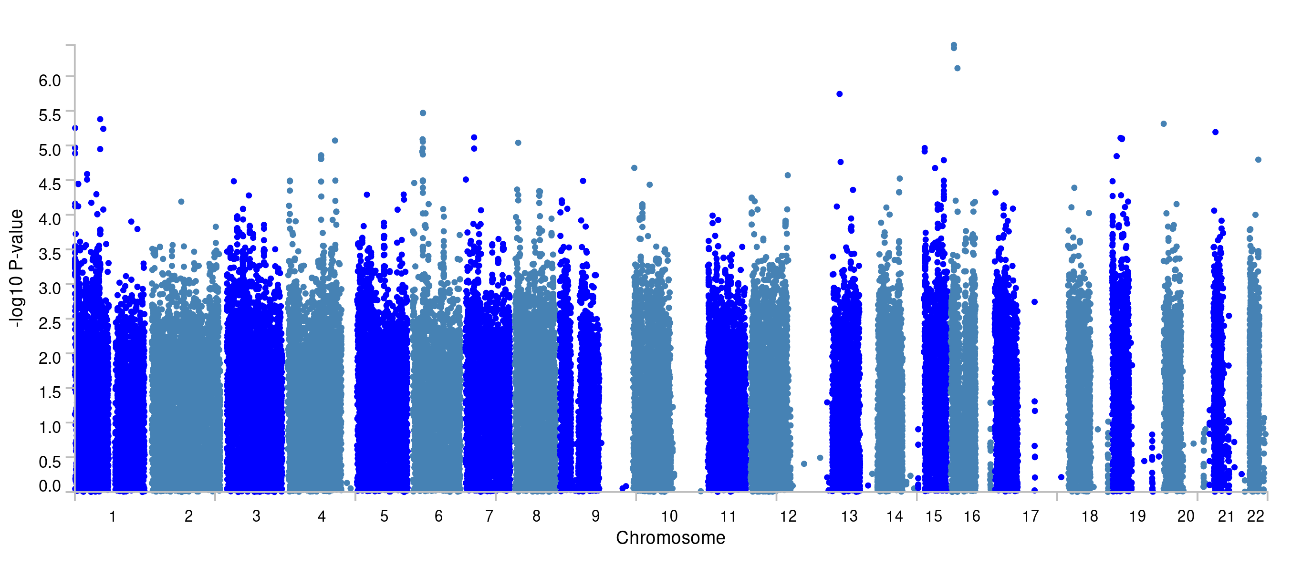 |
| C | 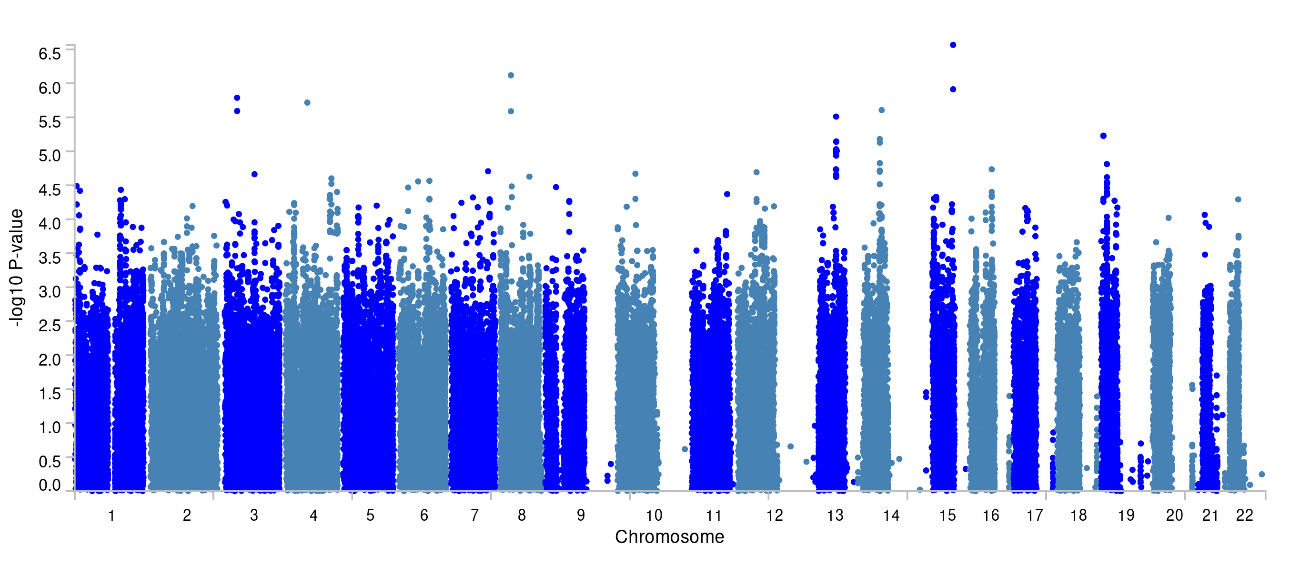 |
| D | 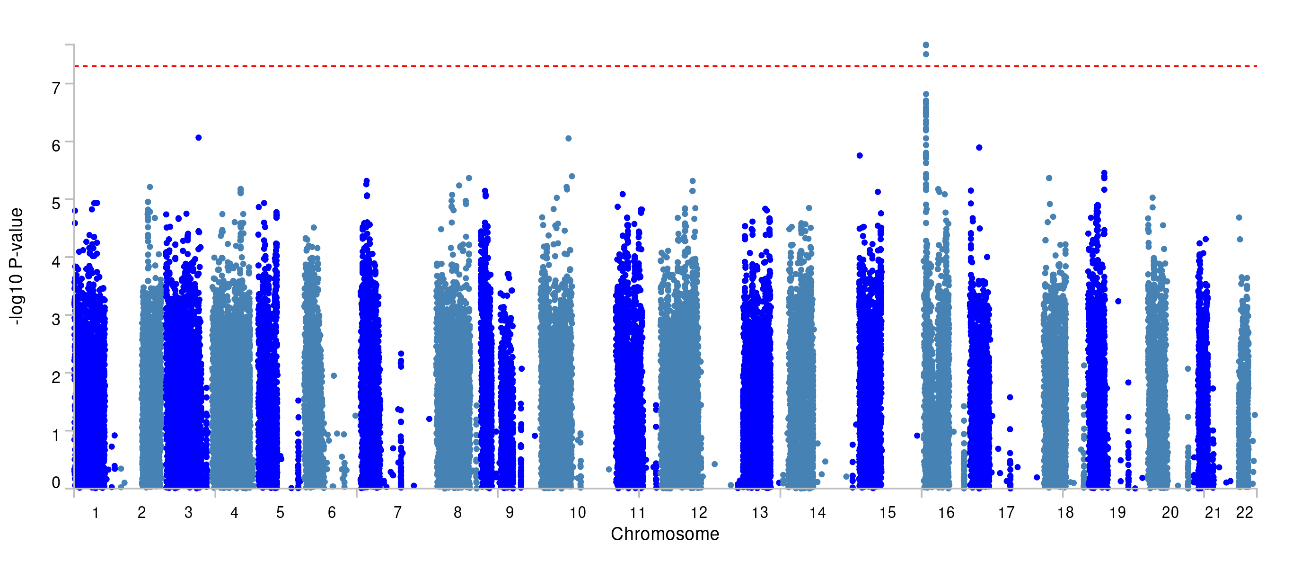 |

Figure 3 Manhattan Plot of GWAS summary statistics for Posterior ILAS. (A) Asian. (B) African American. (C) White. (D) Hispanic.

| A | 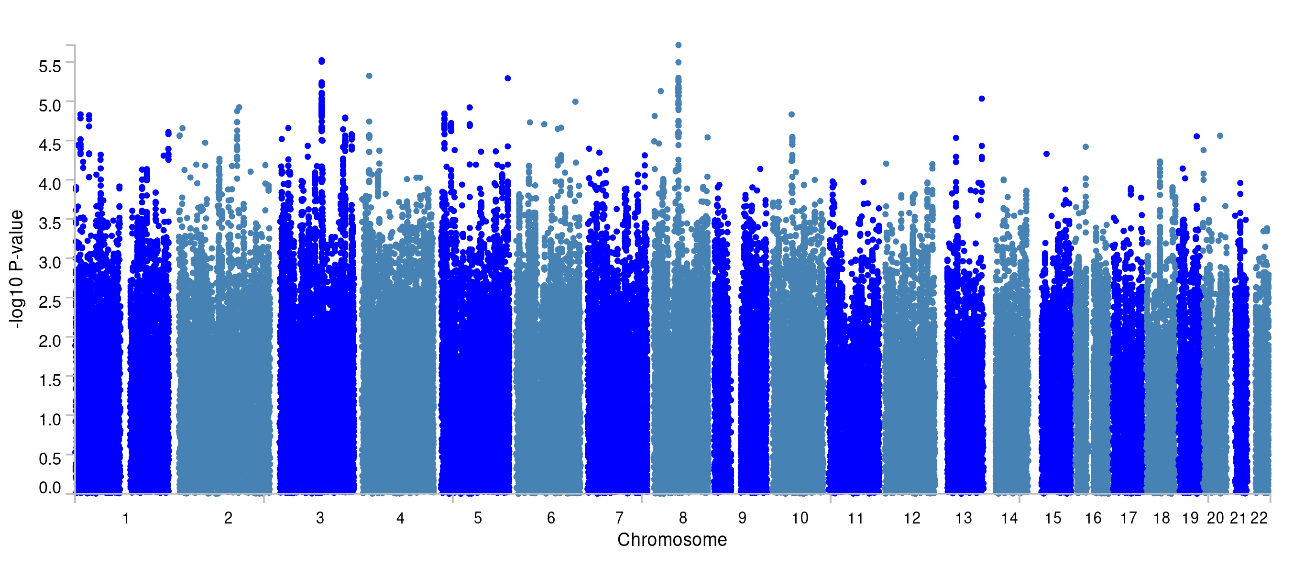 |
| --- | --- |
| B | 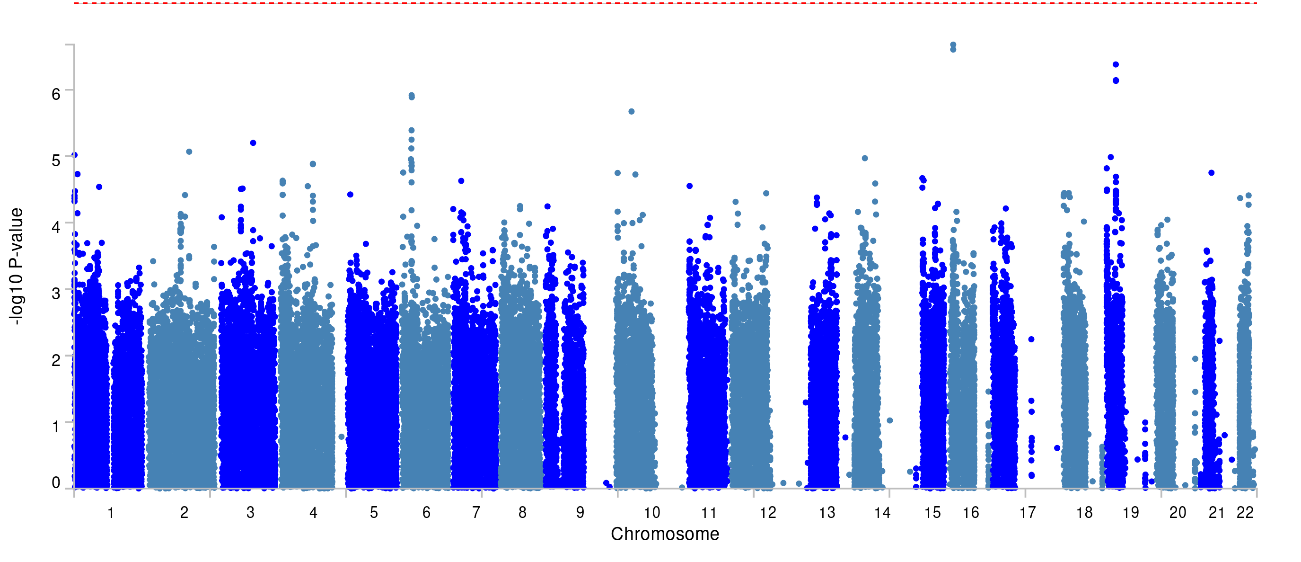 |
| C | 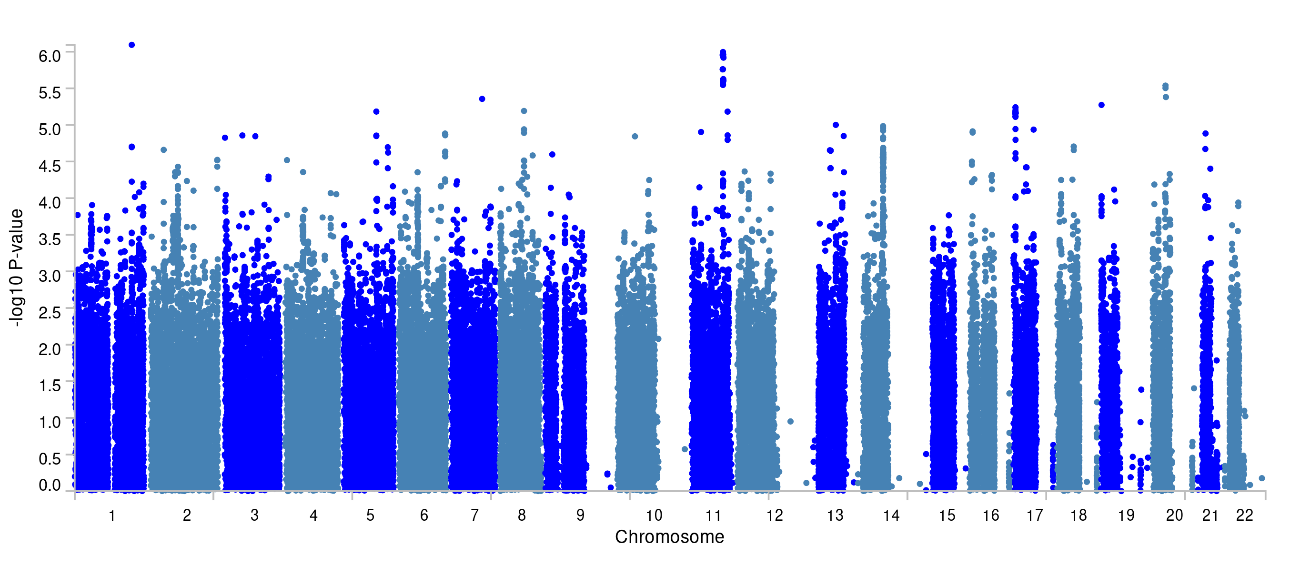 |
| D | 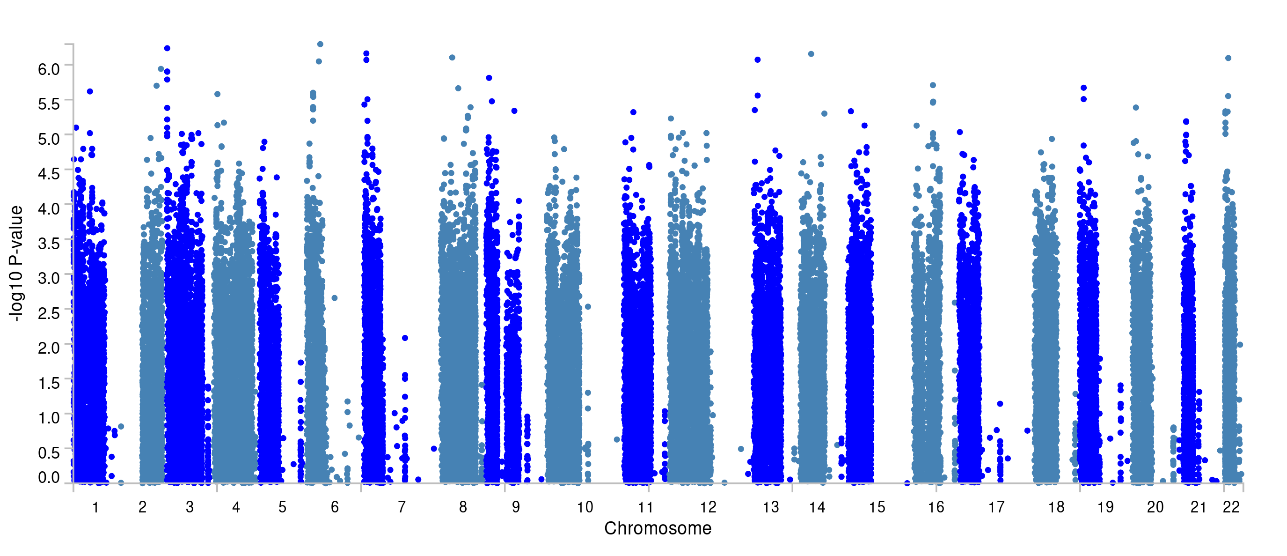 |

Figure 4 QQ plot of GWAS summary statistics for Global ILAS. (A) Asian. (B) African American. (C) White. (D) Hispanic.

| 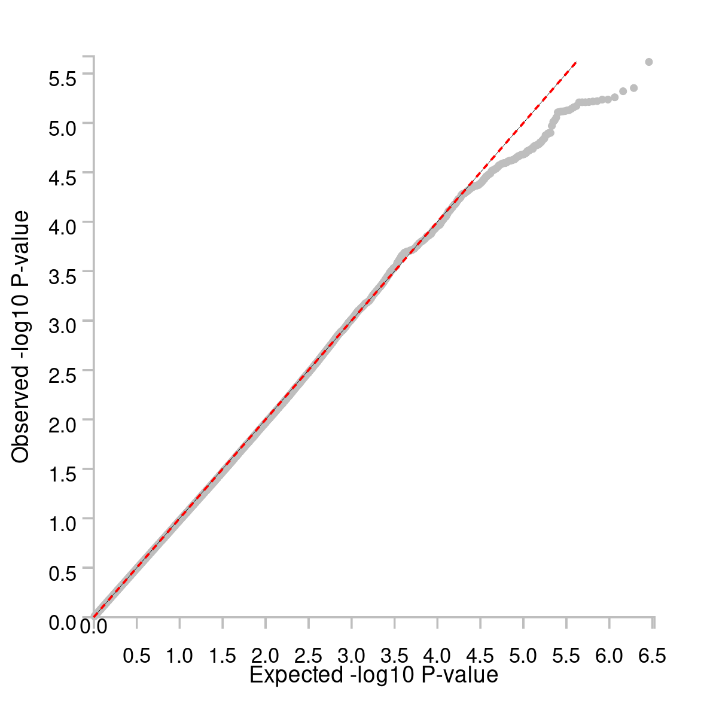 | 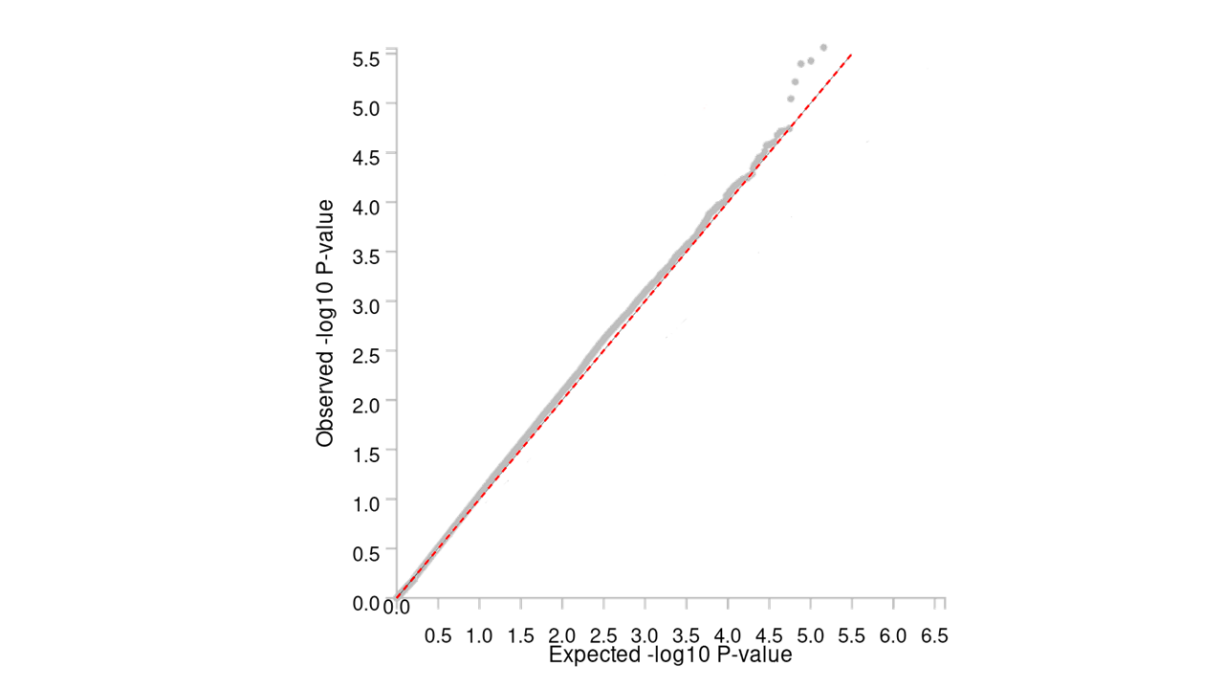 |
| --- | --- |
| A | B |
| 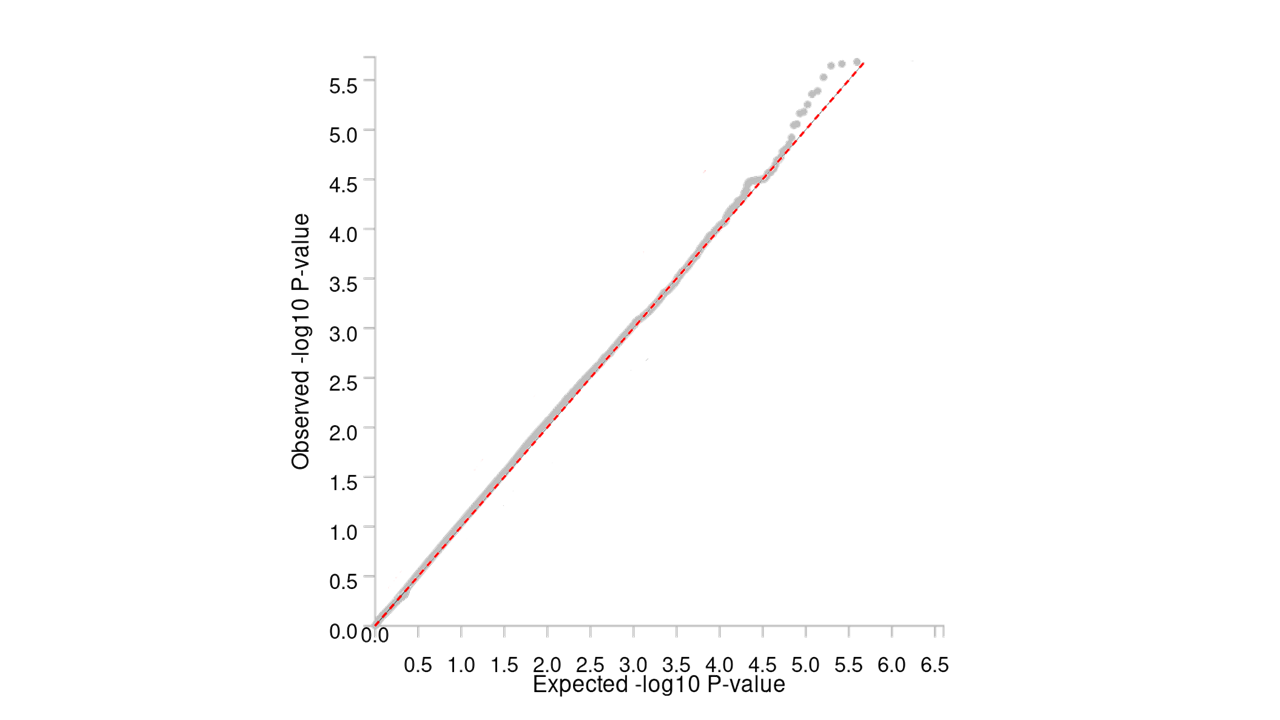 | 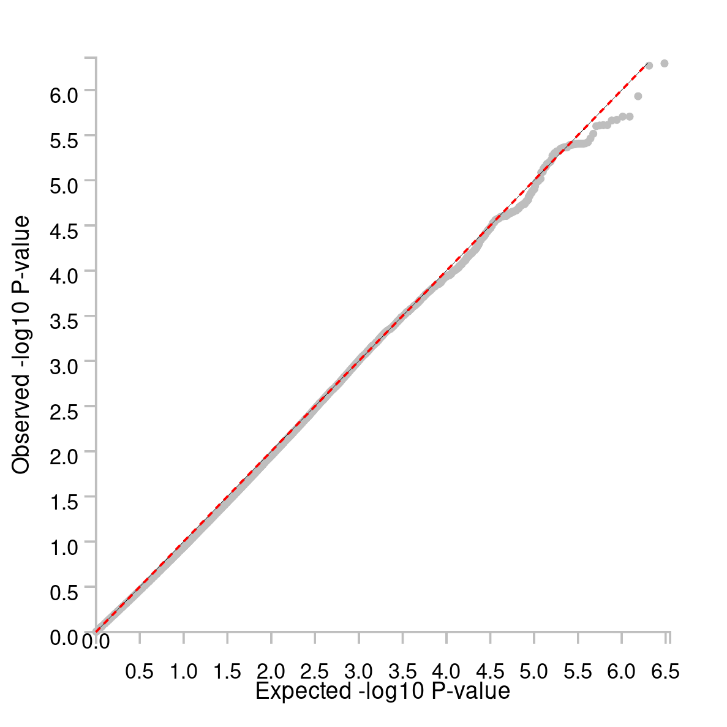 |
| C | D |

Figure 5 QQ plot of GWAS summary statistics for Anterior ILAS. (A) Asian. (B) African American. (C) White. (D) Hispanic.

| 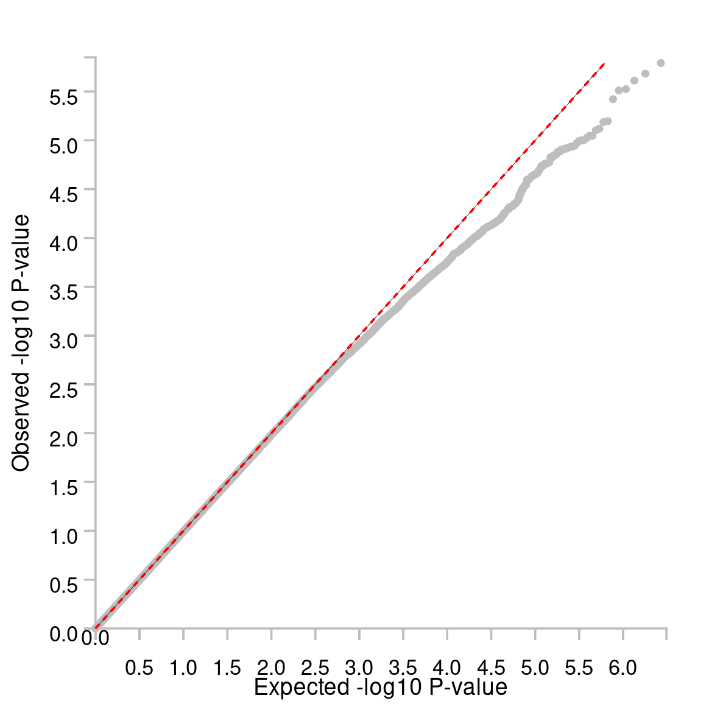 | 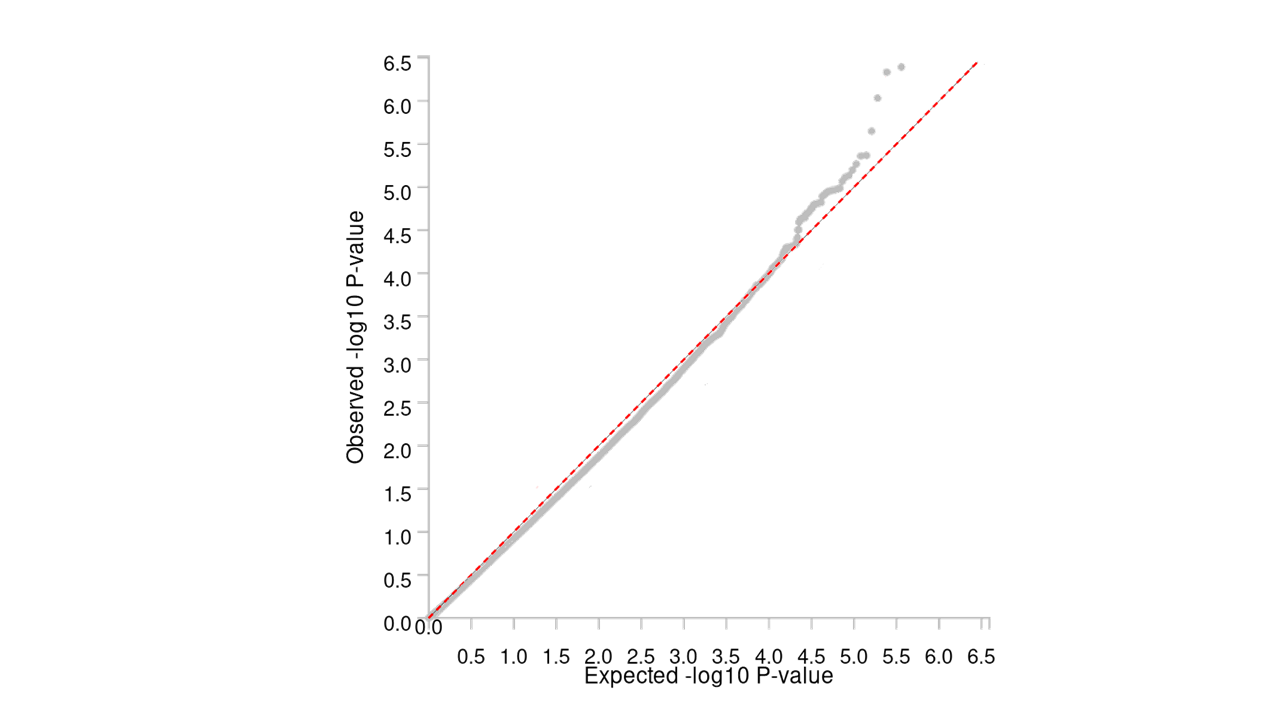 |
| --- | --- |
| A | B |
| 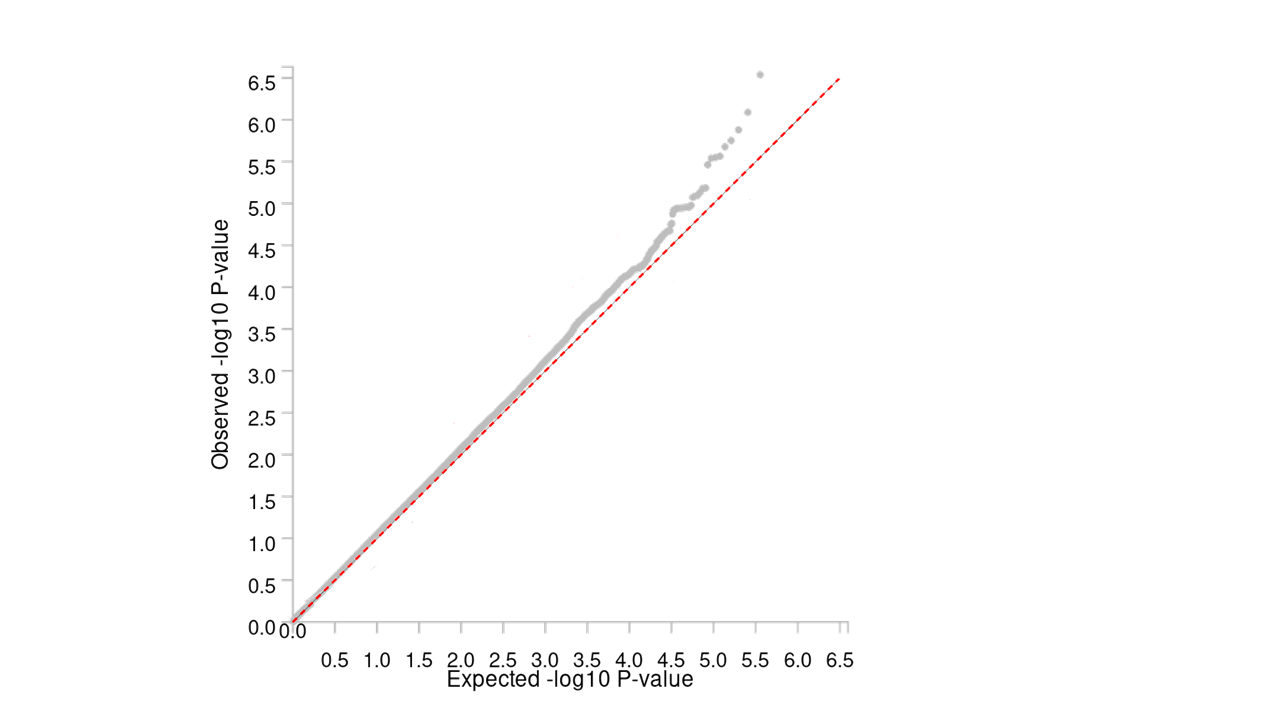 | 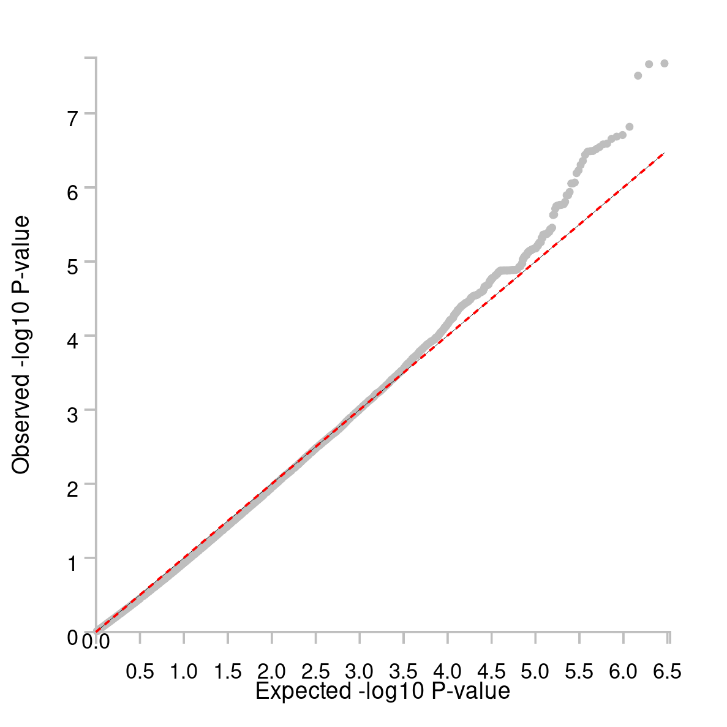 |
| C | D |

Figure 6 QQ plot of GWAS summary statistics for Posterior ILAS. (A) Asian. (B) African American. (C) White. (D) Hispanic.

| 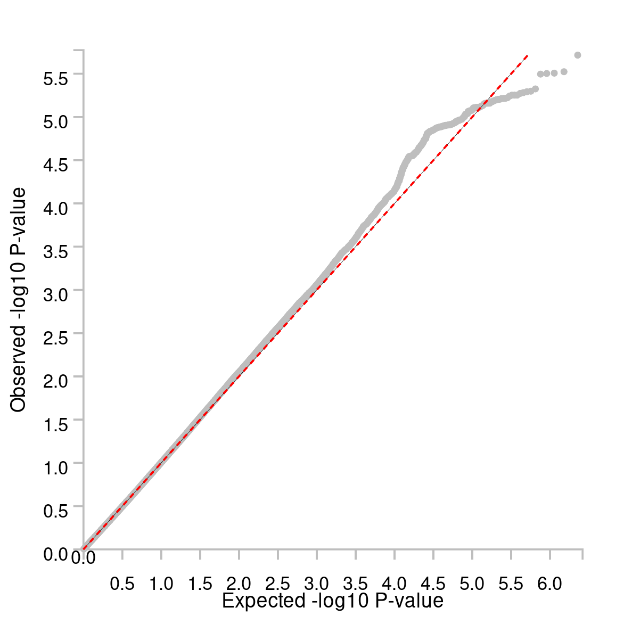 | 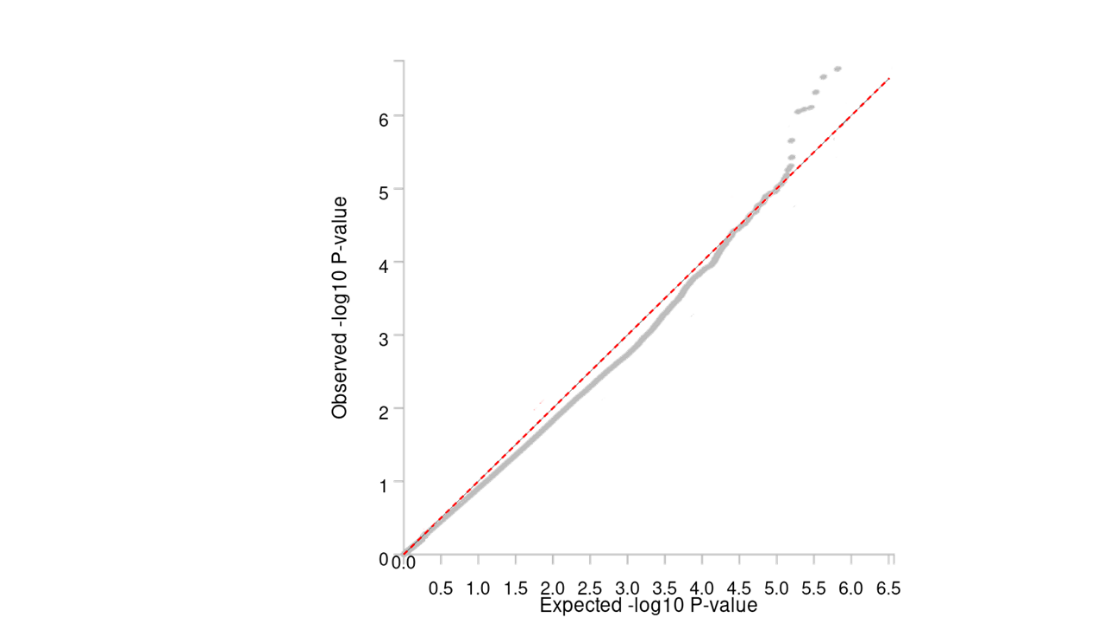 |
| --- | --- |
| A | B |
| 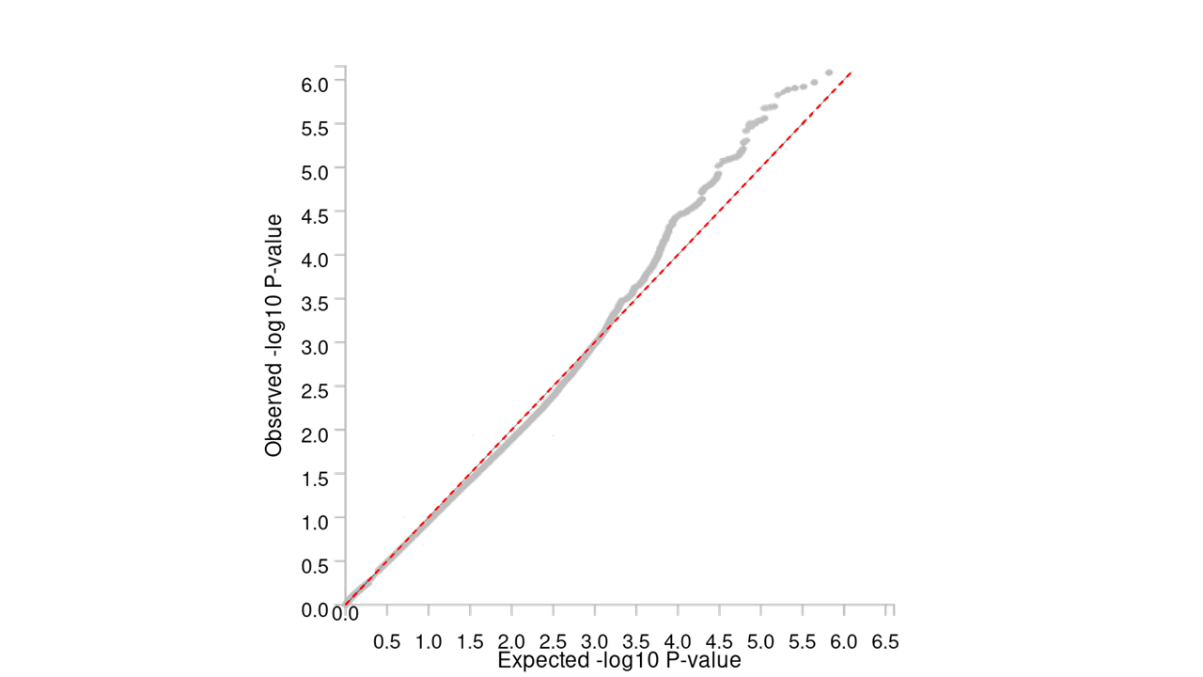 | 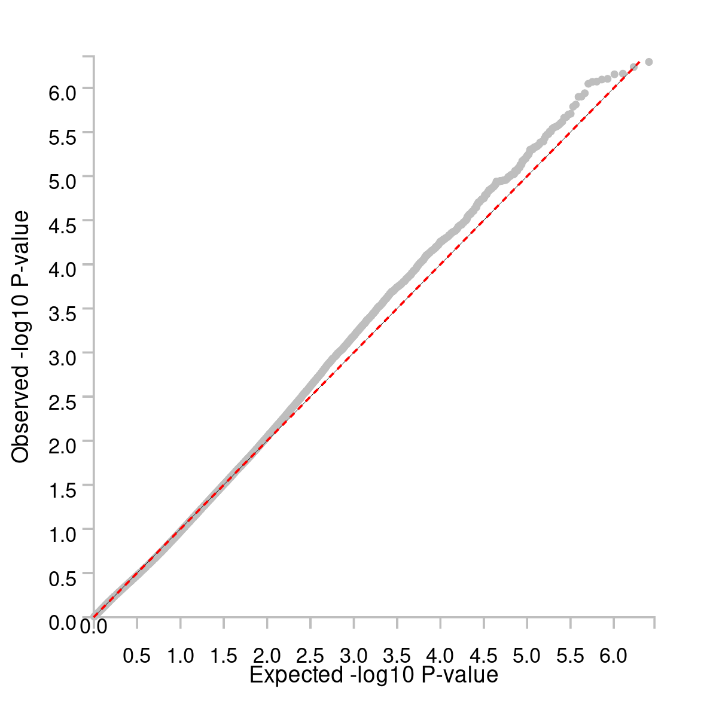 |
| C | D |

Figure 7 Mahattan Plot of gene-based test for Global ILAS. (A) Asian. (B) African American. (C) White. (D) Hispanic.

| A | 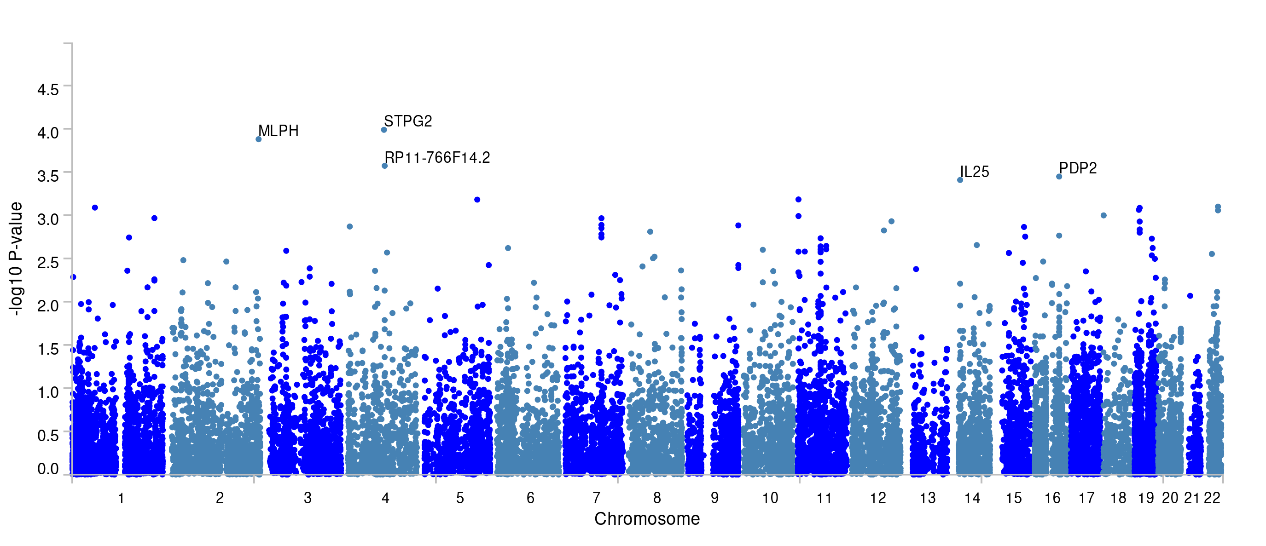 |
| --- | --- |
| B | 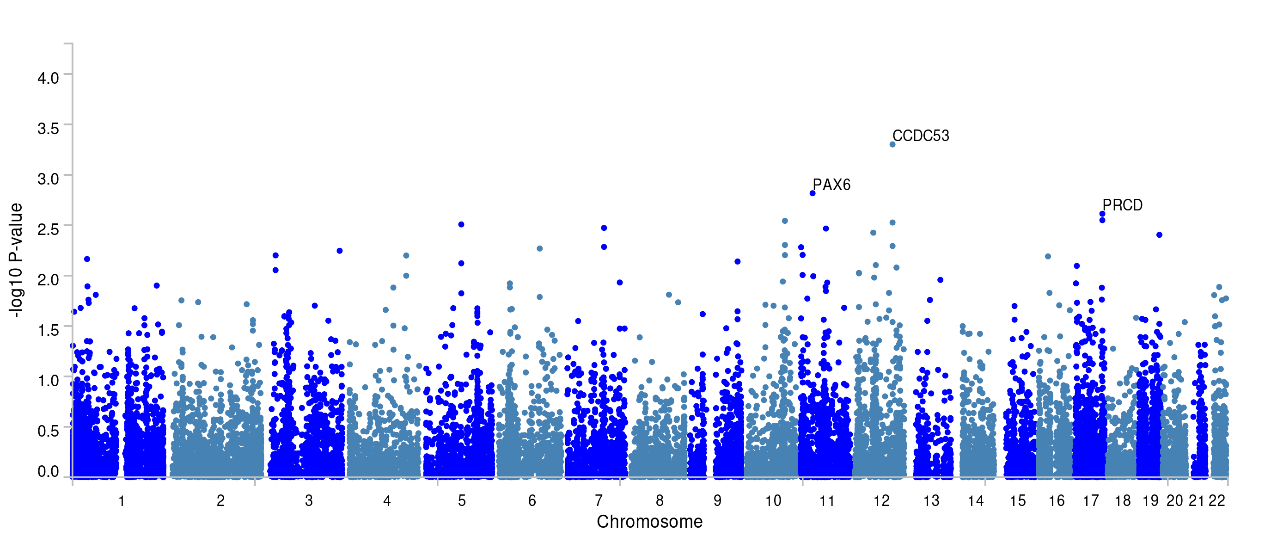 |
| C | 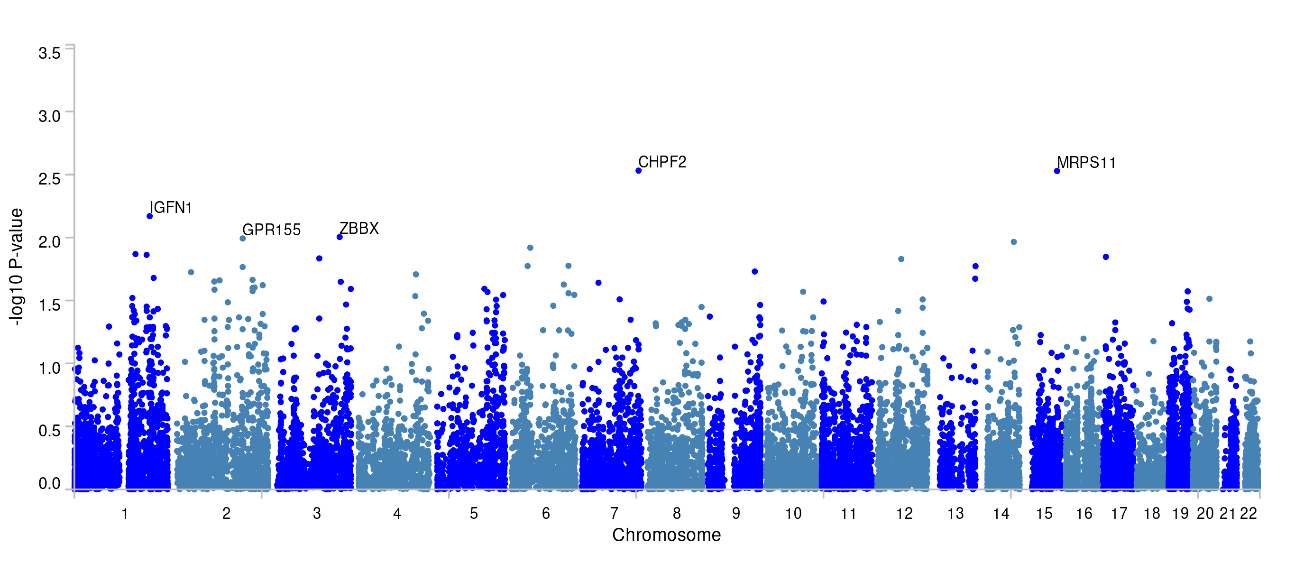 |
| D | 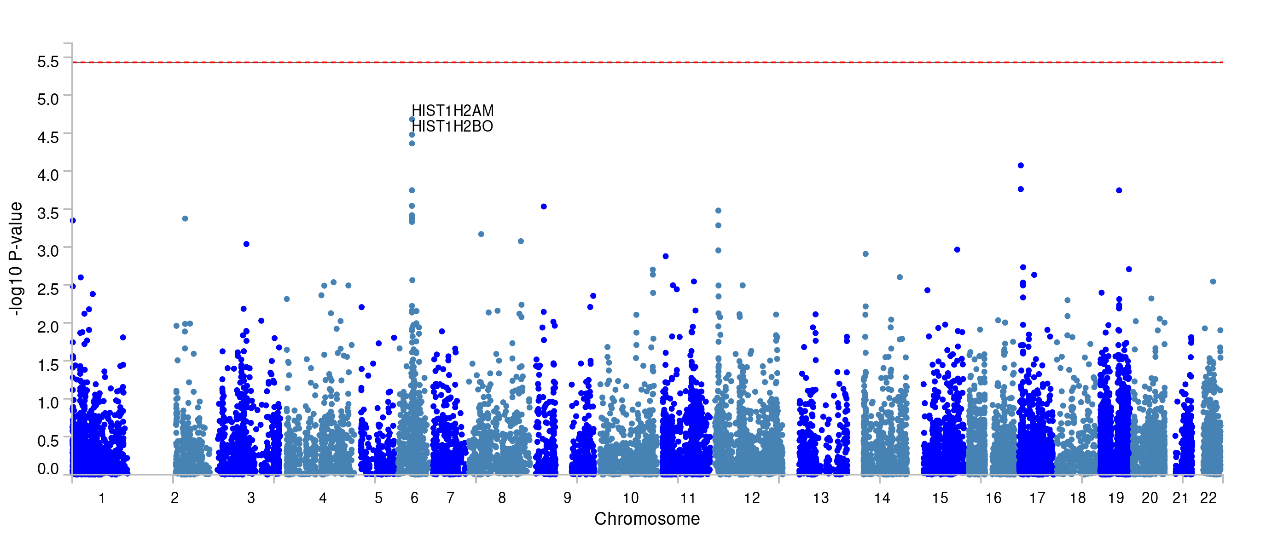 |

Figure 8 Mahattan Plot of gene-based test for Anterior ILAS. (A) Asian. (B) African American. (C) White. (D) Hispanic.

| A | 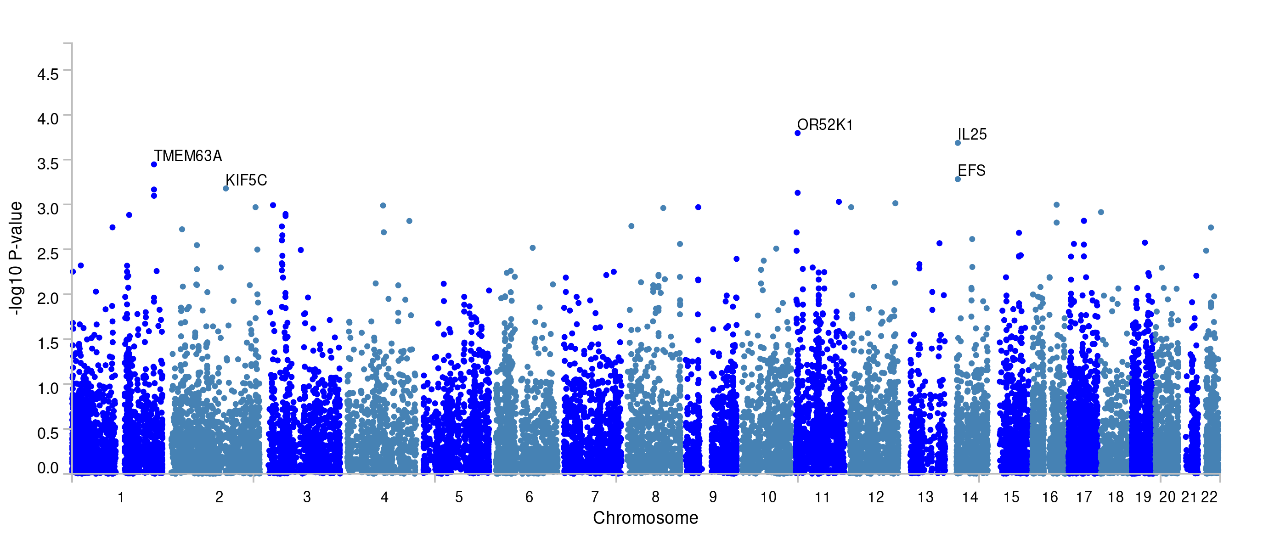 |
| --- | --- |
| B | 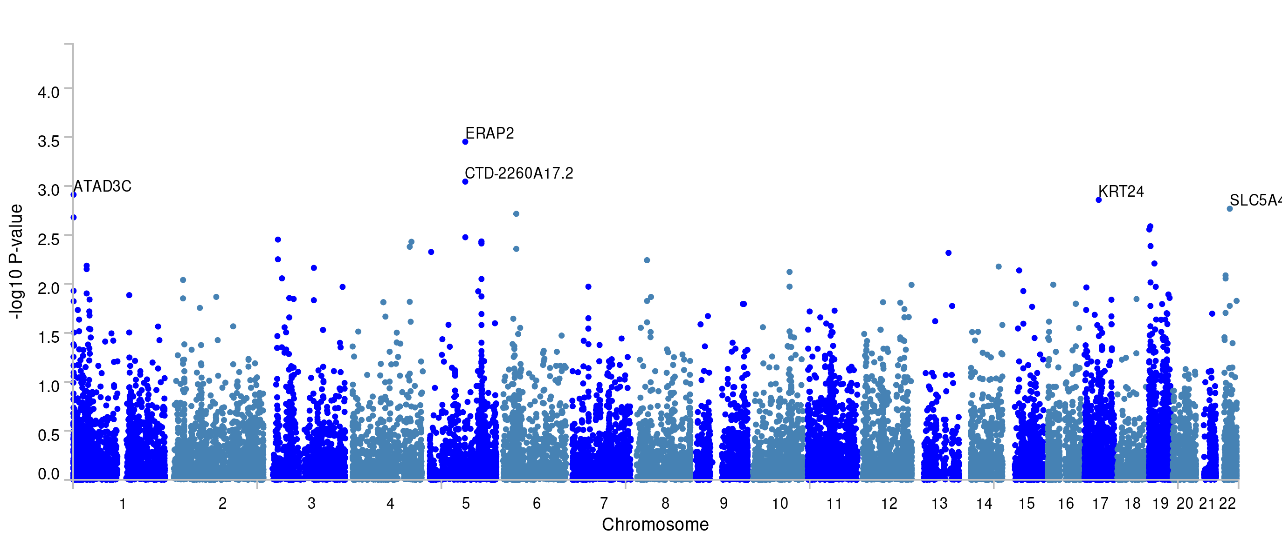 |
| C | 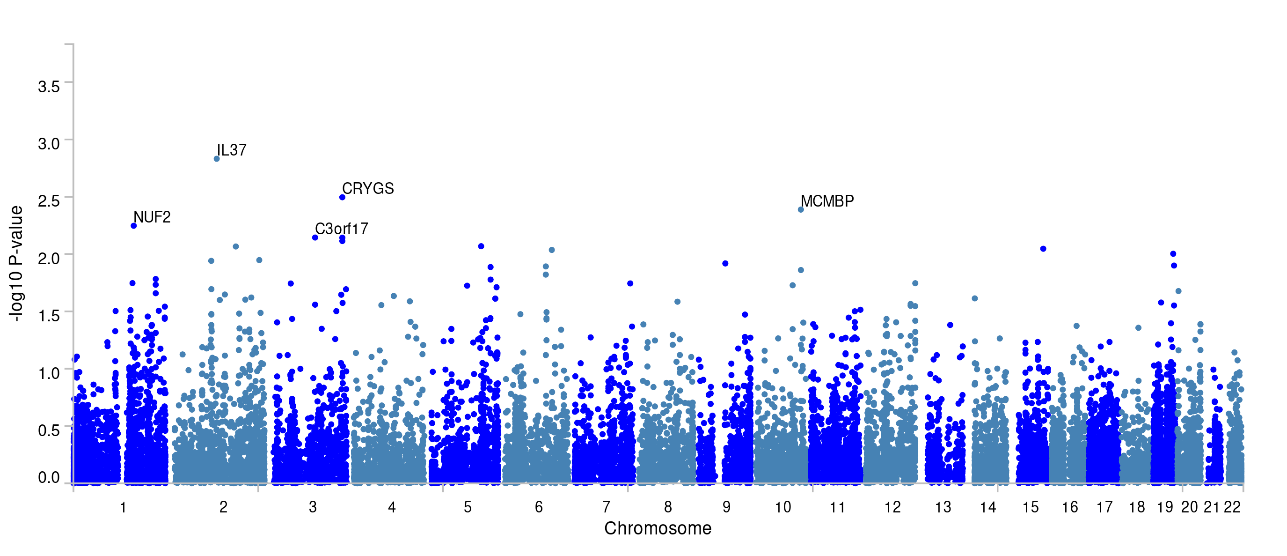 |
| D | 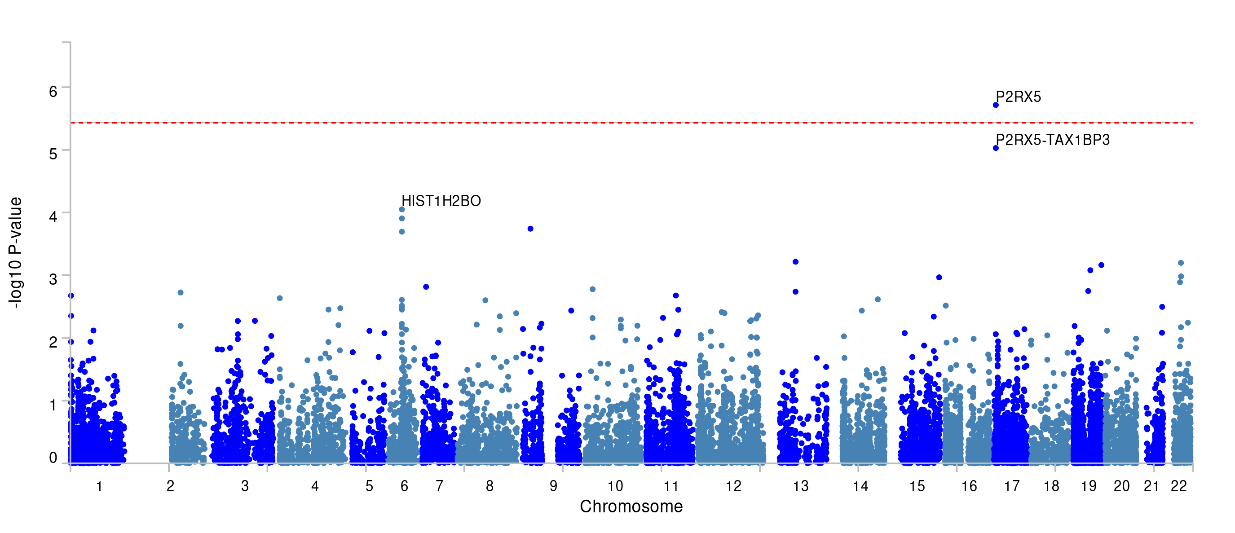 |

Figure 9 Mahattan Plot of gene-based test for Posterior ILAS. (A) Asian. (B) African American. (C) White. (D) Hispanic.

| A | 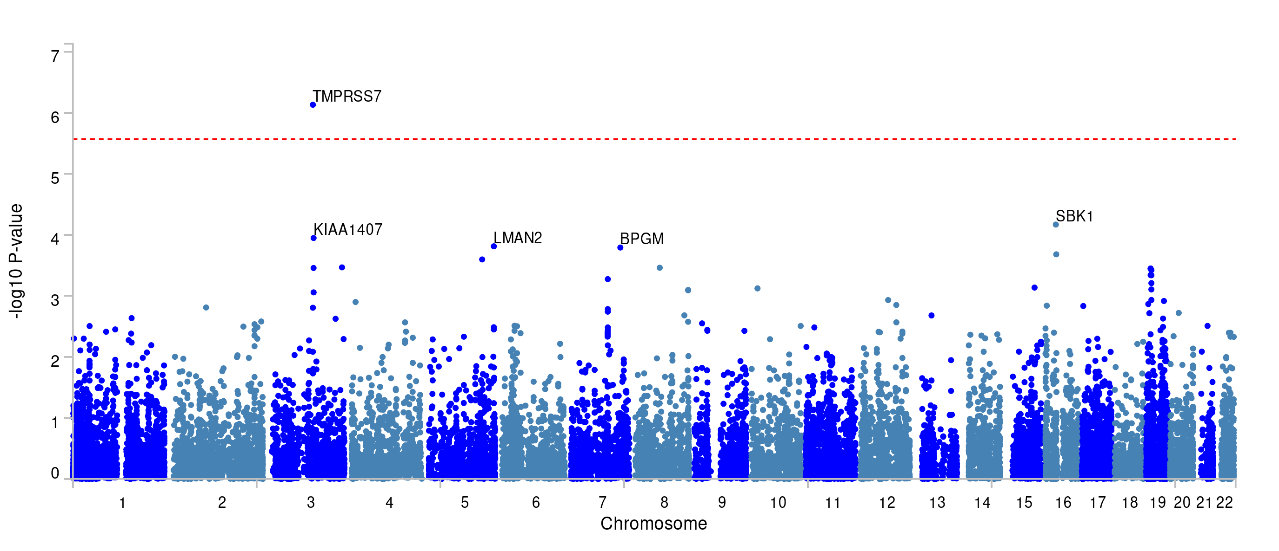 |
| --- | --- |
| B | 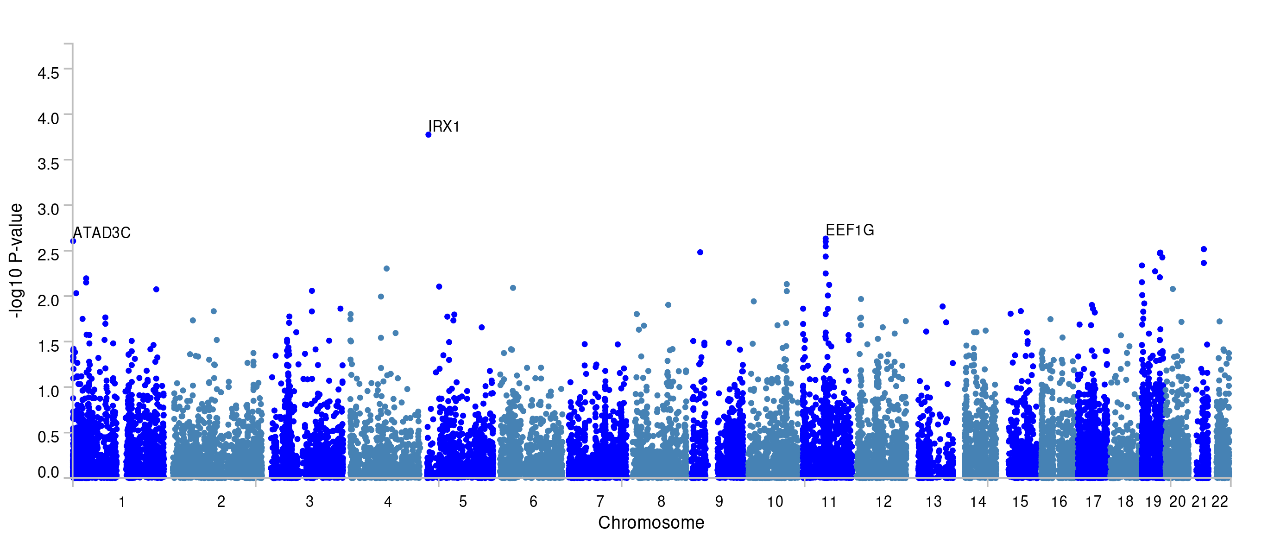 |
| C | 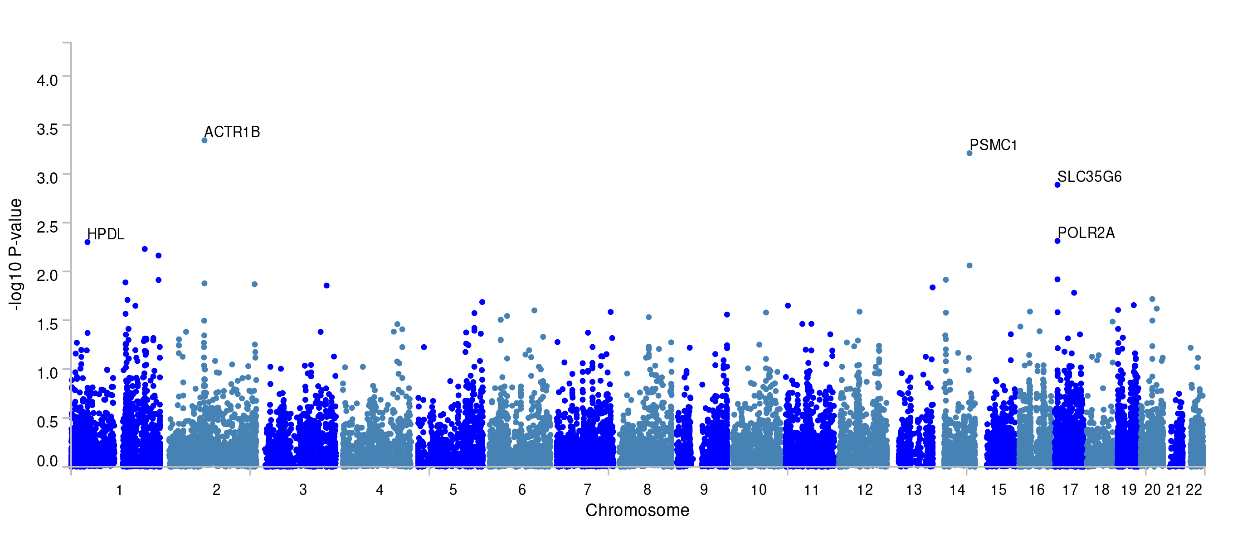 |
| D | 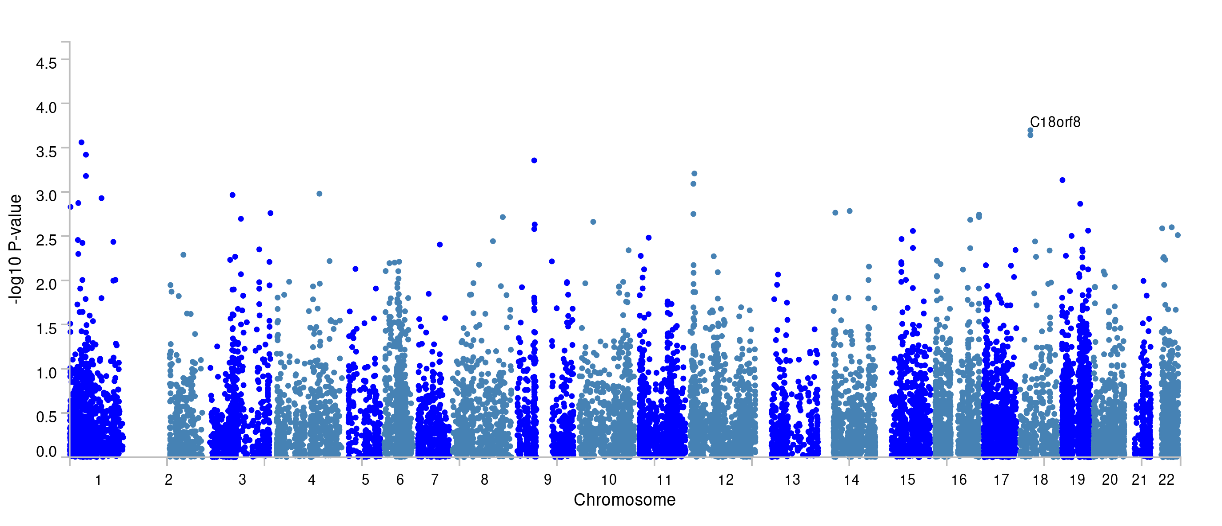 |

Figure 10 Regional plot. (A) rs1528343 (18:8464372) for Hispanic Anterior ILAS.

(B) rs73856305 (3: 111781467) for Asian Posterior ILAS.

| A | 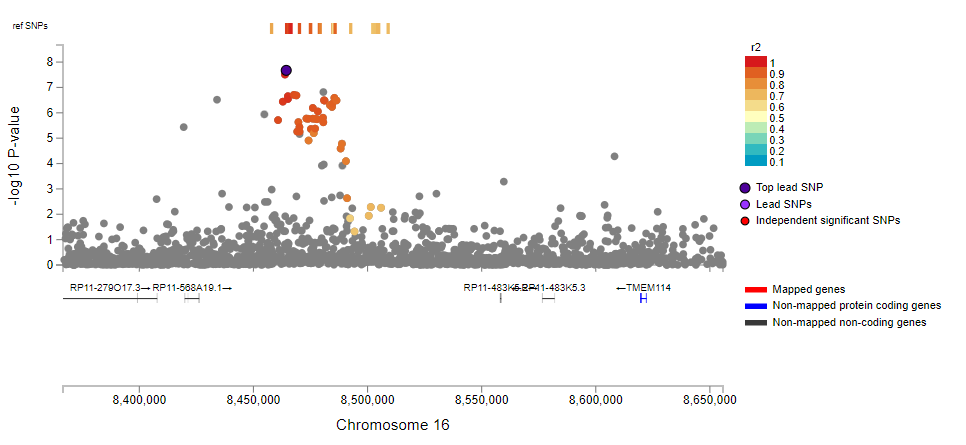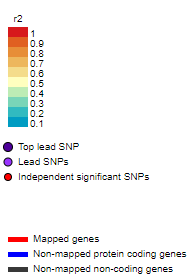 |
| --- | --- |
| B | 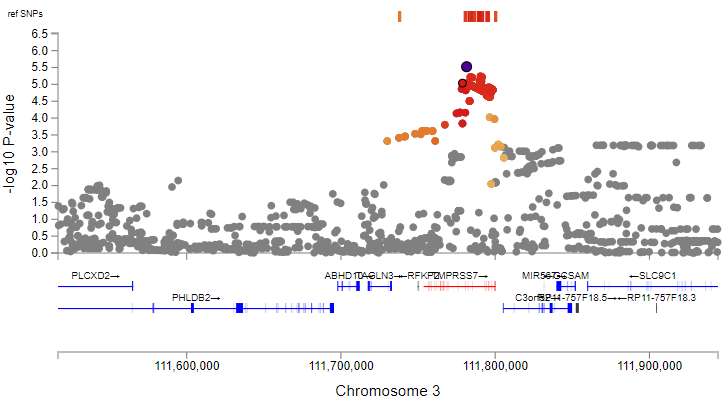 |
